# Harmonization of microbiome data in the Environmental influences on Child Health Outcomes (ECHO) Cohort to analyze environmental influences on the child microbiome

**DOI:** 10.64898/2026.11.06.26367438

**Authors:** Justine W. Debelius, Shaoming Xiao, Noel T. Mueller, Sarah S. Comstock, Kristen L. McArthur, Zone Li, Ekaterina Smirnova, Vanja Klepac-Ceraj, Christian Rosas-Salazar, Cara Wychgram, Meredith Palmore, Grier P. Page, Judy L. Aschner, Patricia A. Brennan, Carlos A. Camargo, Scott Collingwood, Suman R. Das, Janice A. Espinola, James E. Gern, Tina V. Hartert, Daniel J. Jackson, Margaret R. Karagas, Leslie D. Leve, Thomas G. O’Connor, Ryohei Shibata, Meghan H. Shilts, Zhaozhong Zhu, Amii M. Kress, the ECHO Cohort Consortium

**Affiliations:** Department of Psychology Department of Epidemiology, Johns Hopkins Bloomberg School of Public Health, Baltimore, MD, USA; Department of Epidemiology, Colorado School of Public Health, Aurora, CO, USA; Department of Pediatrics, University of Colorado School of Medicine, Aurora, CO, USA; Department of Food Science & Human Nutrition, Michigan State University, East Lansing, MI, USA; Department of Biostatistics, Virgina Commonwealth University, Richmond, VA, USA; Department of Biological Sciences, Biochemistry Program, Wellesley College, Wellesley, MA, USA; Department of Pediatrics, Vanderbilt University Medical Center, Nashville, TN, USA; Genomics and Translational Research Center, RTI International, Atlanta, GA, USA; Department of Pediatrics, Hackensack Meridian School of Medicine, Center for Discovery and Innovation, Nutley, NJ, USA; Department of Psychology, Emory University School of Medicine, Atlanta, GA, USA; Department of Emergency Medicine, Massachusetts General Hospital, Boston, MA, USA; Department of Pediatrics, University of Utah School of Medicine, Salt Lake City, UT, USA; Department of Medicine, Vanderbilt University Medical Center, Nashville, TN, USA; Department of Pediatrics, University of Wisconsin-Madison School of Medicine, Madison, WI, USA; Department of Medicine, University of Wisconsin-Madison School of Medicine, Madison, WI, USA; Department of Epidemiology, Dartmouth Geisel School of Medicine, Hanover, NH, USA; Prevention Science Institute, Oregon State University, Eugene, OR, USA; Department of Psychiatry, University of Rochester Medical Center, Rochester, NY, USA; Department of Neuroscience, University of Rochester Medical Center, Rochester, NY, USA; Department of Obstetrics and Gynecology, University of Rochester Medical Center, Rochester, NY, USA

**Author notes:** **Corresponding Authors:** Justine Debelius, Department of Epidemiology, Johns Hopkins Bloomberg School of Public Health, 615 Wolfe St, Baltimore, MD, USA 21205. See Supplemental File 2 for a list of collaborators (the ECHO Cohort Consortium).

**Keywords:** environmental exposure, microbiota, infant, child, cohort studies

## Abstract

The human microbiome is critical for health across the life course and may mediate the relationship between early-life exposures and health outcomes. The Environmental influences on Child Health Outcomes (ECHO) Cohort provides a unique opportunity to investigate this relationship in children. Individual cohort sites within the ECHO Cohort collected and sequenced microbiome data prior to the consortium formation. Sequencing data were annotated using a common protocol. We summarized the relationship between participant characteristics and features of the microbiome to understand how ECHO can advance the study of the connection between environmental exposures, the microbiome, and health outcomes. A total of 15,509 high-quality extant microbiome samples were collected from 1,086 pregnancies and 5,701 children from 21 ECHO cohort sites. The observed microbial communities were shaped by body site, participant type (pregnant person, child), age, and sample processing. Children lived in 40 states across the United States; high-quality geolocation data were available for more than 60% of children with gut samples. Assay measures, including PFAS, and were available for more than 1,000 children with gut samples. Extant ECHO microbiome data provide an opportunity to characterize the effect of environmental exposures on the developing human microbiome and the role of these exposures in shaping long-term health.

## INTRODUCTION

The human microbiome—the collection of bacteria that live in and on the human body— is critical for health across the lifespan. Differences in the microbiome have been linked to key early-life outcomes, including pre-term birth (1, 2); asthma and atopic disease (3–5); neurodevelopmental outcomes, including autism spectrum disorder (ASD) (6–8); and energy balance and obesity (9, 10). Microbiome alterations associated with environmental exposures have been observed in children and adults, including associations with toxicants (11–15), greenspace (16–18), and community-level social determinants of health (19, 20). Additionally, microbial communities may be effect modifiers or mediators of exposure–disease associations; however, evaluating these relationships requires robust data about the effect of environmental exposures on the microbiome and the influence of the early life microbiome on disease risk (16, 21).

The microbiome is dynamic, with substantial development occurring in the first two years of life. In healthy pregnancies, an infant’s first exposure to microbes occurs at birth; the delivery mode seeds the subsequent microbial communities (22, 23), which is followed by a rapid community assembly and ecological succession (24–26). Microbiome acquisition is shaped by a variety of factors, including breastfeeding, antibiotic use, genetics, family structure, environmental exposures, and cultural practices (16, 27–40). The introduction of solid foods shifts the microbiome toward a more adult community (24–26). While it is possible to change the adult microbiome, there is less plasticity after the microbiome has been established (24–26, 41). Therefore, early life is a critical window for shaping the microbiome with implications for health across the life course.

Large, diverse, multi-site cohort studies are needed to understand the factors that affect the assembly of microbial communities in young children. The Environmental influences on Child Health Outcomes (ECHO) Cycle 1 Cohort is a consortium of 69 geographically diverse pregnancy and pediatric cohort sites in the United States (U.S.) (42). Extensive efforts have been undertaken to harmonize data collected under disparate cohort site protocols to ensure that these data may be analyzed together. Within the ECHO Cohort, there are extant microbiome data, banked specimens for microbiome analysis, and ongoing efforts to collect new specimens for microbiome analysis. Here, we describe the harmonization process for extant microbiome data, present a descriptive analysis of harmonized data from extant microbiome samples within the ECHO Cohort, and discuss opportunities to use these data, along with ECHO’s rich phenotypic and environmental exposure data.

## METHODS

Twenty-two individual cohort sites submitted their previously sequenced microbiome data and associated sample metadata to the ECHO Data Analysis Center (DAC). Gut and vaginal samples were collected from mothers during pregnancy; gut, oral, and upper airway samples were collected from children. Oral samples were only submitted by one cohort site (43–45) and were excluded from subsequent analyses as they could not be compared across multiple cohort sites. Thirteen cohort sites recruited healthy participants and children (46–59); four cohort sites focused on children at increased risk for asthma or other allergic diseases (60–64); one cohort site enrolled children admitted to the neonatal intensive care unit (NICU) at birth (65); and three cohort sites focused on adopted children and their biological and adoptive siblings (66, 67). Details on cohort site source populations, study designs, and inclusion and exclusion criteria have been previously described (42). Table S1 summarizes the cohort site–specific inclusion and exclusion criteria, microbiome-specific inclusion and exclusion criteria, and the original microbiome study design. Cohort site–specific sample collection, storage, extraction amplification, and sequencing are described in the Supplemental Methods and Table S2.

The study protocol was approved by the single ECHO institutional review board, WCG IRB (WCG IRB #20181210). Written informed consent or parent’s/guardian’s permission was obtained along with child assent as appropriate, for the ECHO Cohort Data and Biospecimen Collection Protocol participation and for participation in specific cohort sites.

### Bioinformatic Processing

Our goal was to develop a pipeline that maximized high-quality microbiome data available from each ECHO cohort site while providing common microbiome identifiers for direct comparison. The extant microbiome data were amplified with five primer pairs targeting five hypervariable regions; in some cases, the primer pairs were non-overlapping, meaning direct comparison would require a scaffold (e.g., reference sequence or common taxonomic annotation) (68–70).

Cohort sites followed a standardized data submission protocol for uploading both sequencing data and metadata to a common location via a secure file transfer (Figure S1). Metadata included a description of sample collection, extraction, sequencing, and processing, along with an optional form describing participant characteristics at the time of sample collection. Once uploaded, sequencing data underwent bioinformatic processing at the DAC. Samples were considered eligible for analysis if the sample was found across all uploaded files (i.e., had sequencing data and minimum processing associated metadata) and if the sequencing files had transferred correctly based on a hashed comparison.

Bioinformatics were performed using QIIME 2 (v 2021.11) (71). Initial bioinformatic processing was done on a per-cohort site and per-primer pair basis (e.g., we processed samples amplified with different primers but within the same cohort site separately.) Primer trimming was performed using q2-Cutadapt (Cutadapt v. 3.7) (72); reads were joined in using q2-vsearch if quality and length allowed (vsearch v. 2.7.0; Table 3) (73); and quality filtered using the q2-quality-filter plugin with default parameters (71, 74). Reads were denoised with the Deblur algorithm (Table S3) (75). We chose a common trim length of 150 nucleotides (nt) for samples amplified with primers from the Earth Microbiome Project (515F-806R or 515F-926R) to allow the direct use of amplicon sequence variants (ASVs), a single nucleotide-resolved microbiome feature; this approach has been used in previous large studies of harmonized data with these primer pairs (76, 77). Per-cohort processing parameters were retrieved through provenance extract action using code based on the q2-provenance-replay functionality in QIIME 2 (78). Table S3 describes the bioinformatic parameters for each cohort site and primer pair combination.

We selected the Silva reference database, a popular pan-environmental database for samples collected across all body sites (79). The use of the QIIME 2 RESCRIPt plugin to prepare this database can be found in the Supplemental Methods (80). To provide a common, directly comparable scaffold across all samples, we clustered sequences into operational taxonomic units (OTUs) at 97% identity against the Silva reference database using vsearch (Table S3) (73, 79). In this technique, features are assigned a common identifier based on the reference database sequence they match to. Taxonomy is inherited from that reference database, and phylogenetic relationships are inferred from a tree built based on the full-length reference sequences. Construction of this tree is described in the Supplemental Methods.

ASVs provide a more specific way to identify microbes in 16S rRNA sequencing data; however, they are only comparable with the same primer pair and trim length (81). Therefore, we prepared an ASV table for all samples amplified with 515F (Earth Microbiome Project) primers (Tables S2 and S3) (82). Taxonomy for ASVs is inferred based on a classifier; the preparation and application of a region-specific taxonomic classifier using the Silva 138.1 reference data can be found in the Supplemental Methods (79, 82, 83). The ASV-associated phylogenetic tree was constructed using fragment insertion via the SEPP algorithm into the Greengenes 13_8 reference tree (84, 84).

Species-level annotation is important for vaginal samples. To produce a species-level table for vaginal samples, we annotated with Optivag, a publicly available, manually curated database of vaginal taxa (85). The two-step taxonomic classification process is described in the Supplemental Methods. Annotated features were combined into a species-level assignment. Phylogeny cannot be calculated for species-level data from Optivag.

### Diversity Calculations

Alpha (within-sample) and beta (between-sample) diversity were calculated for both the Silva 138.1 OTU table and 515F 150nt ASV table. For each table, rarefied tables were calculated at 1,000, 2,500, 5,000, and 10,000 sequences/sample as the average of ten rarefaction instances. A rarefaction curve suggested diversity was stabilized at these depths (Figures S2, S3). Alpha diversity was measured using observed features, Shannon diversity (86), Simpson dominance (87), and Pielou’s evenness (88) at all rarefaction depths. Beta diversity was measured using Jaccard distance (89), Bray Curtis dissimilarity (90), unweighted UniFrac distance (91, 92), and weighted UniFrac distance (92, 93) at all rarefaction depths. The UniFrac metrics were omitted for the species-level vaginal data as no phylogenetic tree was available. All diversity calculations were performed using the QIIME 2 q2-diveristy plugin (v. 2023.7) (71).

### Harmonization of Cohort-Provided Sample Characteristics

#### Sample Collection and Processing

Cohort sites submitted sample processing metadata as part of the standardized ECHO data protocol, including the extraction kit, primer pair, hypervariable region, sequencing platform, and read length. Primer pair sequences were standardized to reflect a single representative sequence.

#### Antibiotic Use in the Month Prior to Microbiome Sample Collection

A subset of cohort sites provided optional information on selected medication use within one month of microbiome sample collection. A participant was considered exposed to antibiotics if they reported antibiotic use within a month of sample collection. Participants were considered unexposed if: (i) they reported no medication use in the last month; (ii) they reported medication use but no antibiotic use or uncategorized medications in the last month; (iii) they reported medication use and antibiotics were not among the list of specific medications; or (iv) antibiotic use within a month of sample collection was an exclusion criterion.

#### Infant Diet at the Time of Microbiome Sample Collection

Information about infant diet was first ascertained based on an optional cohort site report at the time of sample collection, followed by previously harmonized infant feeding practices. Children were categorized into “exclusive breastmilk,” “any breastmilk,” and “no breastmilk” at the time of sample collection based on: (i) comprehensive feeding practices reported by the cohort site; (ii) binary breastmilk consumption reported by the site; and in the absence of both, (iii) sample collection before the end of exclusive breastfeeding (defined as breastmilk with supplements but no solid foods) or before the end of breastfeeding, with or without solid food according to the harmonized infant feeding practices. Solid food initiation was derived first from cohort site–reported infant diet, and when this was missing, comparing the age at microbiome sample collection to the age at solid food initiation.

### Participant Characteristics

Linkage with participant characteristics, environmental exposures, and health outcomes are described in the Supplemental Methods. Briefly, demographic and anthropometric characteristics, residential history timeline, and birth and health outcomes were harmonized across multiple cohort sites as previously described (42, 94). Participants were considered to have residential data if their address could be geocoded to U.S. Census tract level or better. Targeted assay data (cytokines; tobacco metabolites; metals, including arsenic, zinc, copper, and mercury; and organic contaminants, including phthalates, per- and polyfluoroalkyl substances (PFAS), phenols, organophosphate flame retardants (OPFRs), polycyclic aromatic hydrocarbons (PAHs), and insecticides), genetic data and epigenetic data within ECHO have been previously described (42, 95, 96).

### Statistical Analysis and Visualization

Full analyses are described in the Supplemental Methods. Briefly, the number of participants in each category of participant characteristics and exposures were tabulated; percentage values were calculated, excluding missing values. Continuous variables were summarized as the mean and standard deviation, median and interquartile region, and range, and the count for missing values. Maps were created using Arc-GIS (v 3.3.0; ESRI, Redlands, CA, USA). Base figures were generated with seaborn (v. 0.2.12) and matplotlib (v. 3.6).(97, 98) Figures were assembled; fonts were updated; and icons were added using Adobe Illustrator (San Jose, CA, USA). Icons came from the Font Awesome library (v. 6.0) or the National Institutes of Health (NIH) BioArt Gallery, or were generated for this project.

### Example analysis

To demonstrate the utility of ECHO’s extant microbiome data in analysis, we analyzed the relationship between children’s age at sample collection, diet at sample collection, and birth-associated covariates. Participants were eligible for this complete case analysis if they contributed at least one high quality fecal sample collected between 1 and 5 months of age; had data for sex, delivery mode, and diet at sample collection; were born after at least 37 weeks completed gestation and had not been admitted to the NICU. Shannon alpha diversity and unweighted UniFrac distance were selected as representative metrics for alpha and beta diversity (86, 91). For untargeted genus-level differential abundance, we filtered to retain genera present with a relative abundance of at least 0.001 in at least 5% of samples from at least two cohort sites. Counts were given an offset of 1 and the data was central log ratio transformed. Alpha diversity and genus level CLR measures were modeled using a linear regression adjusted for cohort site with child age (3 month units) and breastmilk consumption at sample collection, sex, and delivery mode. Benjamini-Hochberg multiple hypothesis correction was applied to genus level tests. Modeling was done using statsmodels (v.0.14.1) (99). Beta diversity was modeled using an adonis test in the R vegan library (2.6.1) (100, 101). Cohort site, child age, breastmilk consumption, sex, and delivery mode were included as fixed effects and R^2^ values were estimated by margin. P-values are based on 999 permutations.

## RESULTS

Overall, 22 ECHO cohort sites submitted 16,543 samples from 1,125 pregnant people (1,153 pregnancies), 5,815 children, and 42 sequencing controls. Following quality filtering, denoising, clustering into closed reference OTUs, and filtering to sample types found in multiple cohort sites, we retained 15,509 unique samples from 1,061 pregnant participants (1,086 pregnancies) and 5,701 children at 21 ECHO cohort sites (Figure S4). Samples were collected across three primary body sites, including the gastrointestinal tract (children and pregnant participants), upper airway (children only), and vagina (pregnant participants only; Figure 1). The body site sampled was a major determinant of the microbiome structure (Figure 1).

**Figure 1.**
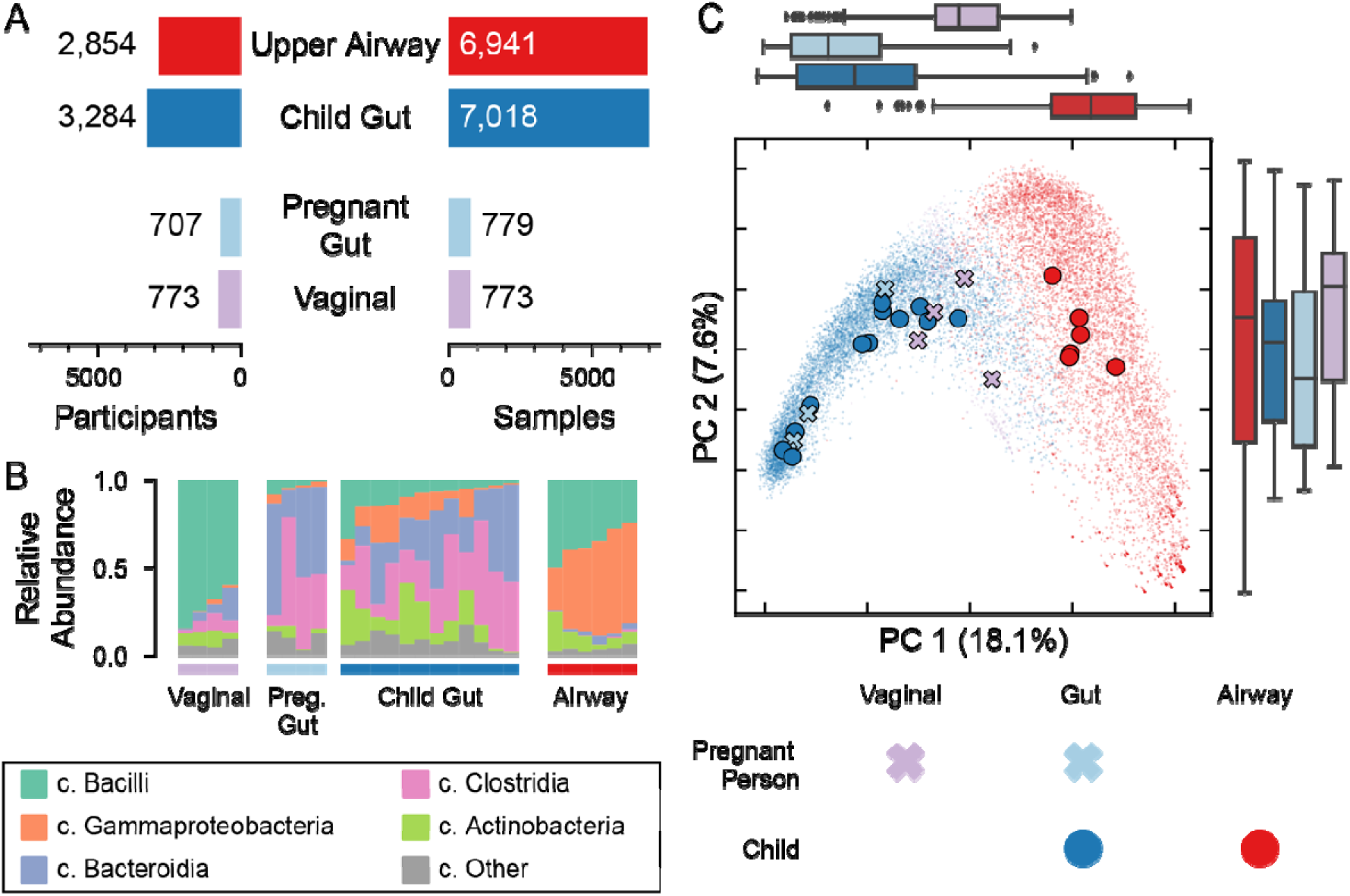
Microbiome samples in ECHO. (A) The number of samples and participants with available microbiome data by participant type and body site. The number of unique pregnancies or children are shown on the left; the number of samples are shown on the right. (B) Bar plot of the relative abundance of class-collapsed taxonomy, grouped by body site and participant type. Each bar represents the average relative abundance within an ECHO cohort site. (C) Principal Coordinates projection of unweighted UniFrac distance between all unique samples with at least 1000 sequences/sample. Each point represents a sample; samples are colored by the body site and participant type. Samples have a relative opacity; darker regions indicate more samples in that location. Marginal plots show the distribution of samples along the two axes. Large points indicate the average position for each ECHO cohort site by participant type and body site. ECHO, Environmental influences on Child Health Outcomes.

### Pregnancy Gut and Vaginal Samples

After quality filtering, we retained 773 vaginal samples from four cohort sites. Each sample was unique to a pregnancy. Samples were collected from a total of 740 people; 16 people were sampled during two or more pregnancies. Additionally, 696 participants with 707 pregnancies provided 779 gut microbiome samples at four ECHO cohort sites. In most pregnancies, gut samples were collected once; 43 (6.1%) pregnancies were sampled two or more times. Two cohort sites submitted both gut and vaginal samples for the same pregnancy (n=394 participants). Table 1 describes the characteristics for all pregnancies with microbiome samples. The observed vaginal communities were primarily *Lactobacillus*-dominant (Figure S5, S6). In both vaginal (Figure S5) and gut samples (Figure S9), the extant microbiome data reflect choices made by the cohort sites regarding the type of sample collected (rectal swab vs. fecal sample), the extraction kit, and the primer pair used (Figure S8, S10) (102–104).

**Table 1.**
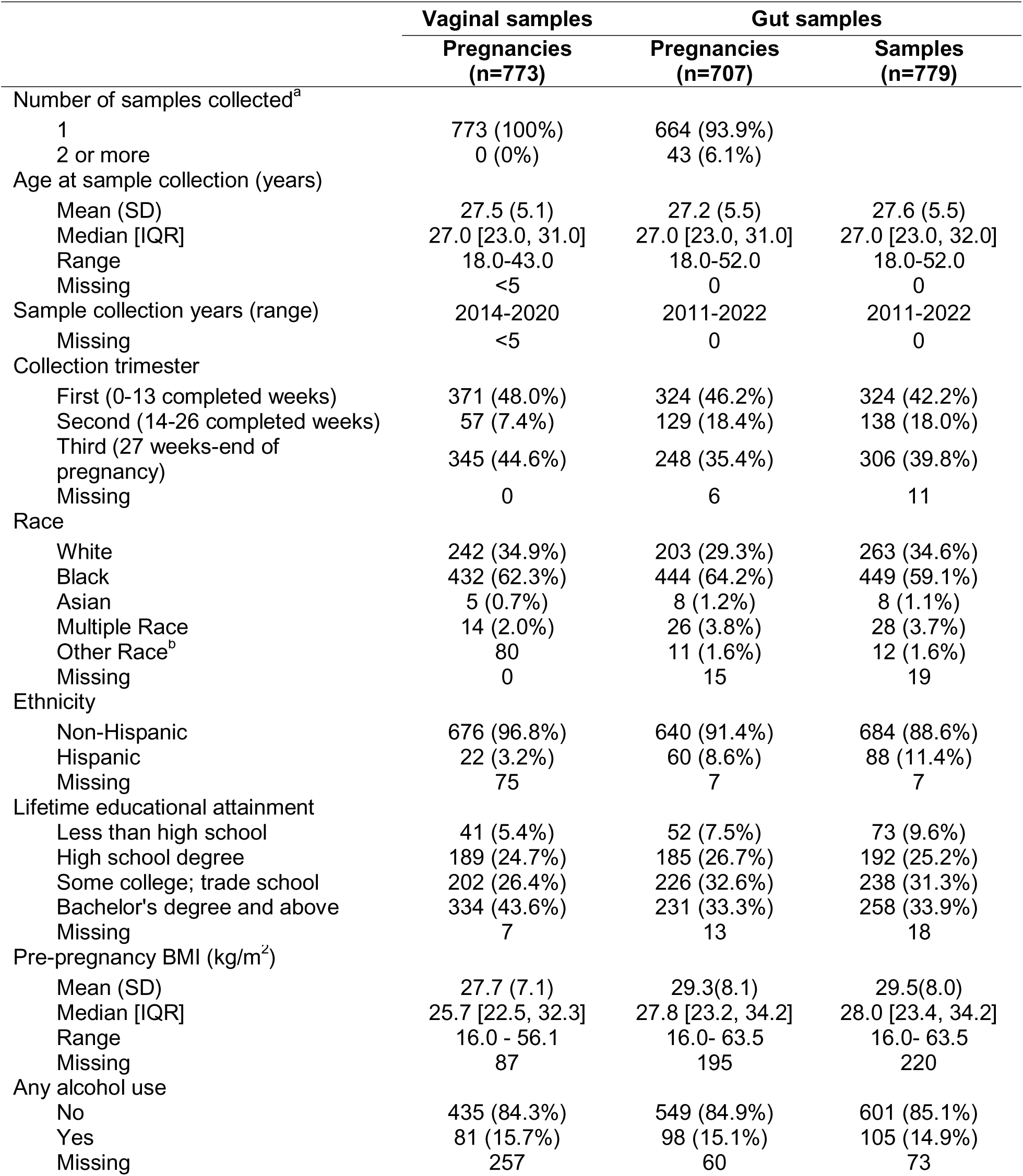
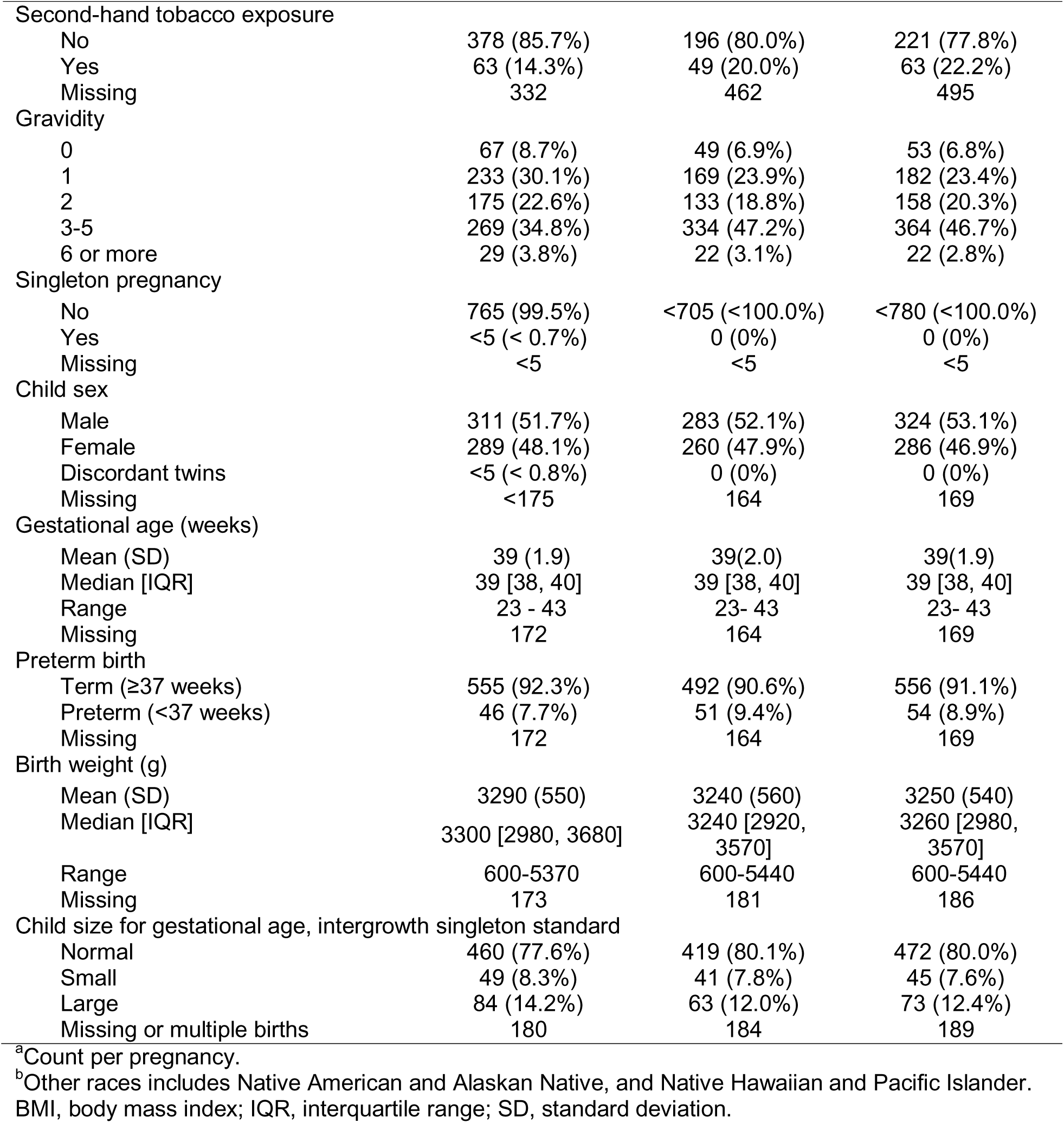
Pregnancy characteristics by sample type.

|  | Vaginal samples | Gut samples |  |
|---|---|---|---|
|  | Pregnancies<br>(n=773) | Pregnancies<br>(n=707) | Samples<br>(n=779) |
| Number of samples collected<sup>a</sup> |  |  |  |
| 1 | 773 (100%) | 664 (93.9%) |  |
| 2 or more | 0 (0%) | 43 (6.1%) |  |
| Age at sample collection (years) |  |  |  |
| Mean (SD) | 27.5 (5.1) | 27.2 (5.5) | 27.6 (5.5) |
| Median [IQR] | 27.0 [23.0, 31.0] | 27.0 [23.0, 31.0] | 27.0 [23.0, 32.0] |
| Range | 18.0-43.0 | 18.0-52.0 | 18.0-52.0 |
| Missing | <5 | 0 | 0 |
| Sample collection years (range) | 2014-2020 | 2011-2022 | 2011-2022 |
| Missing | <5 | 0 | 0 |
| Collection trimester |  |  |  |
| First (0-13 completed weeks) | 371 (48.0%) | 324 (46.2%) | 324 (42.2%) |
| Second (14-26 completed weeks) | 57 (7.4%) | 129 (18.4%) | 138 (18.0%) |
| Third (27 weeks-end of pregnancy) | 345 (44.6%) | 248 (35.4%) | 306 (39.8%) |
| Missing | 0 | 6 | 11 |
| Race |  |  |  |
| White | 242 (34.9%) | 203 (29.3%) | 263 (34.6%) |
| Black | 432 (62.3%) | 444 (64.2%) | 449 (59.1%) |
| Asian | 5 (0.7%) | 8 (1.2%) | 8 (1.1%) |
| Multiple Race | 14 (2.0%) | 26 (3.8%) | 28 (3.7%) |
| Other Race<sup>b</sup> | 80 | 11 (1.6%) | 12 (1.6%) |
| Missing | 0 | 15 | 19 |
| Ethnicity |  |  |  |
| Non-Hispanic | 676 (96.8%) | 640 (91.4%) | 684 (88.6%) |
| Hispanic | 22 (3.2%) | 60 (8.6%) | 88 (11.4%) |
| Missing | 75 | 7 | 7 |
| Lifetime educational attainment |  |  |  |
| Less than high school | 41 (5.4%) | 52 (7.5%) | 73 (9.6%) |
| High school degree | 189 (24.7%) | 185 (26.7%) | 192 (25.2%) |
| Some college; trade school | 202 (26.4%) | 226 (32.6%) | 238 (31.3%) |
| Bachelor's degree and above | 334 (43.6%) | 231 (33.3%) | 258 (33.9%) |
| Missing | 7 | 13 | 18 |
| Pre-pregnancy BMI (kg/m<sup>2</sup>) |  |  |  |
| Mean (SD) | 27.7 (7.1) | 29.3(8.1) | 29.5(8.0) |
| Median [IQR] | 25.7 [22.5, 32.3] | 27.8 [23.2, 34.2] | 28.0 [23.4, 34.2] |
| Range | 16.0 - 56.1 | 16.0- 63.5 | 16.0- 63.5 |
| Missing | 87 | 195 | 220 |
| Any alcohol use |  |  |  |
| No | 435 (84.3%) | 549 (84.9%) | 601 (85.1%) |
| Yes | 81 (15.7%) | 98 (15.1%) | 105 (14.9%) |
| Missing | 257 | 60 | 73 |
| Second-hand tobacco exposure |  |  |  |
| No | 378 (85.7%) | 196 (80.0%) | 221 (77.8%) |
| Yes | 63 (14.3%) | 49 (20.0%) | 63 (22.2%) |
| Missing | 332 | 462 | 495 |
| Gravidity |  |  |  |
| 0 | 67 (8.7%) | 49 (6.9%) | 53 (6.8%) |
| 1 | 233 (30.1%) | 169 (23.9%) | 182 (23.4%) |
| 2 | 175 (22.6%) | 133 (18.8%) | 158 (20.3%) |
| 3-5 | 269 (34.8%) | 334 (47.2%) | 364 (46.7%) |
| 6 or more | 29 (3.8%) | 22 (3.1%) | 22 (2.8%) |
| Singleton pregnancy |  |  |  |
| No | 765 (99.5%) | <705 (<100.0%) | <780 (<100.0%) |
| Yes | <5 (< 0.7%) | 0 (0%) | 0 (0%) |
| Missing | <5 | <5 | <5 |
| Child sex |  |  |  |
| Male | 311 (51.7%) | 283 (52.1%) | 324 (53.1%) |
| Female | 289 (48.1%) | 260 (47.9%) | 286 (46.9%) |
| Discordant twins | <5 (< 0.8%) | 0 (0%) | 0 (0%) |
| Missing | <175 | 164 | 169 |
| Gestational age (weeks) |  |  |  |
| Mean (SD) | 39 (1.9) | 39(2.0) | 39(1.9) |
| Median [IQR] | 39 [38, 40] | 39 [38, 40] | 39 [38, 40] |
| Range | 23 - 43 | 23- 43 | 23- 43 |
| Missing | 172 | 164 | 169 |
| Preterm birth |  |  |  |
| Term (≥37 weeks) | 555 (92.3%) | 492 (90.6%) | 556 (91.1%) |
| Preterm (<37 weeks) | 46 (7.7%) | 51 (9.4%) | 54 (8.9%) |
| Missing | 172 | 164 | 169 |
| Birth weight (g) |  |  |  |
| Mean (SD) | 3290 (550) | 3240 (560) | 3250 (540) |
| Median [IQR] | 3300 [2980, 3680] | 3240 [2920, 3570] | 3260 [2980, 3570] |
| Range | 600-5370 | 600-5440 | 600-5440 |
| Missing | 173 | 181 | 186 |
| Child size for gestational age, intergrowth singleton standard |  |  |  |
| Normal | 460 (77.6%) | 419 (80.1%) | 472 (80.0%) |
| Small | 49 (8.3%) | 41 (7.8%) | 45 (7.6%) |
| Large | 84 (14.2%) | 63 (12.0%) | 73 (12.4%) |
| Missing or multiple births | 180 | 184 | 189 |

<sup>a</sup>Count per pregnancy.
<sup>b</sup>Other races includes Native American and Alaskan Native, and Native Hawaiian and Pacific Islander.
BMI, body mass index; IQR, interquartile range; SD, standard deviation.

One of the ECHO Cohort’s major strengths is the integration of rich, harmonized participant data. ECHO enables integrated microbiome research, including high-quality geocoding for area-level exposures, panels of individual-level targeted assays, and multi-omic profiling (Figure 2). Pregnant participants lived in 10 U.S. states; we successfully geolocated the primary residential locations of 472 (66.8%) participants with gut samples and 440 (59.6%) participants with vaginal samples over the full duration of their pregnancy. Targeted assays were categorized into four main classes: cytokines, tobacco metabolites, metals, and environmental pollutants. At least one targeted assay was available for 582 participants (82.3%) with gut microbiome samples, and 459 (59.3%) participants with vaginal microbiome samples. Additionally, we noted 566 pregnancies where a vaginal or gut sample was collected during pregnancy and the child’s gut microbiome was also profiled. These parent-child dyads have been explored in more detail in a recent publication (105).

**Figure 2.**
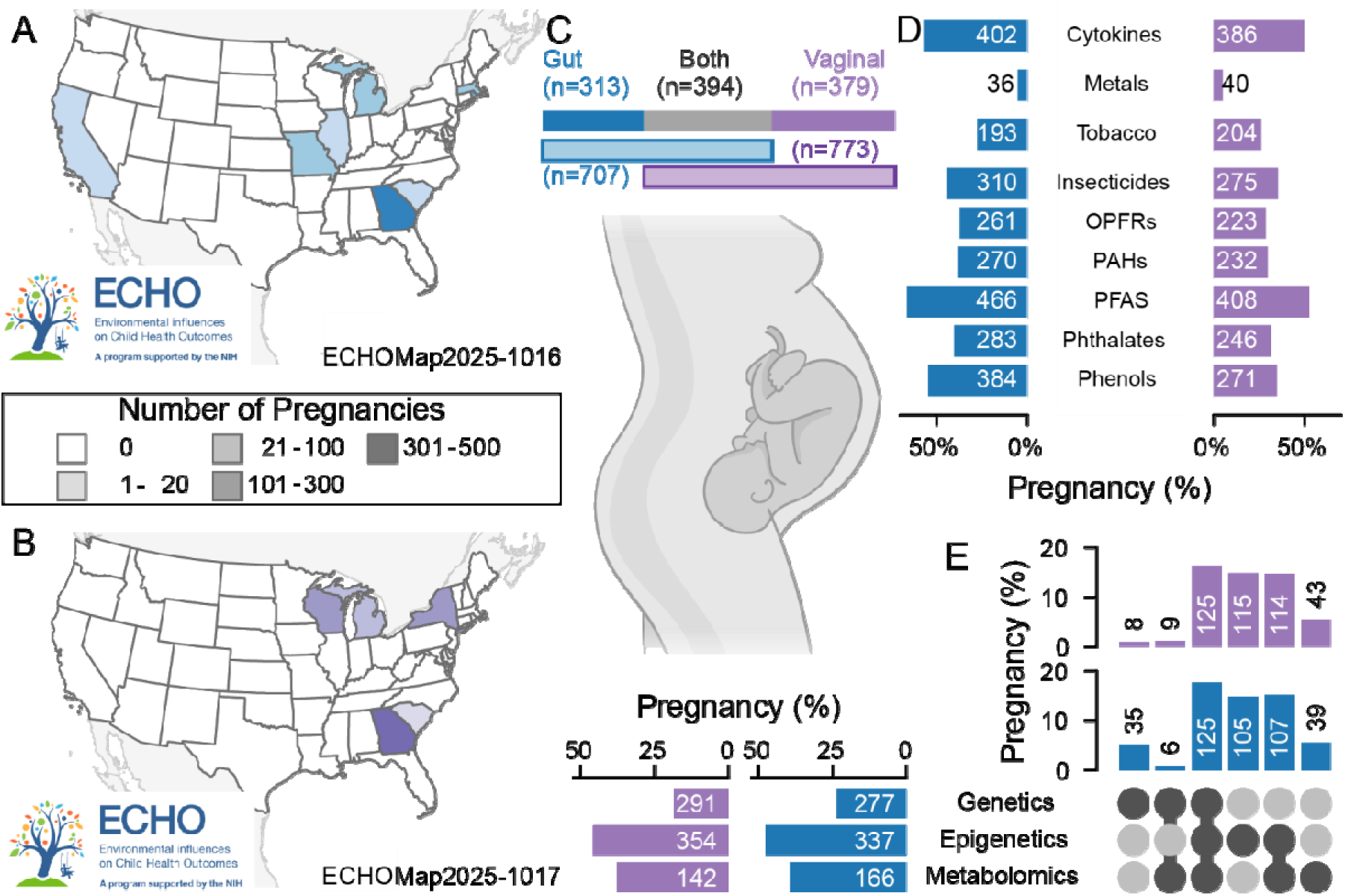
Available environmental and geographic data for pregnant participants with microbiome samples in ECHO. (A, B) The state of residence at the time of microbiome sample collection for (A) gut and (B) vaginal samples. No samples were collected from pregnancies in Alaska, Hawaii, or Puerto Rico. (C) The number of pregnant participants with gut samples only, vaginal samples only, or both. (D) The portion of pregnancies with different types of chemical assays for pregnancies with gut (blue) or vaginal (purple) microbiome samples. The numbers provide the specific count of participants with that assay. (E) Upset plot shows the portion of participants with genetic, epigenetic, or metabolomic data during pregnancy. Horizontal bars show the total number of participants with each assay by sample type (left/top, purple: vaginal; right/lower blue: gut). The dotted plot shows the intersection between the omics; the bars on top show the size of the intersection. ECHO, Environmental influences on Child Health Outcomes.

### Child Gut Samples

Our dataset includes 7,018 unique child gut samples from 3,284 children at 14 ECHO cohort sites. Two cohort sites collected rectal swabs from 326 participants; the remaining 12 sites collected fecal samples from 2,955 participants. Among children with gut samples, 1,473 (44.9%) contributed one sample; 1,096 (33.4%) contributed two, and 712 (21.7%) contributed three or more (Table 2). There were 189 families with multiple children, including 405 children who contributed 1,066 samples. Samples were collected between the first month of life and 16 years old. Notably, 2,151 children provided 3,952 samples before their first birthday (Table S4). We found that sample type (rectal swab or fecal sample) and participant age were major determinants of the observed gut microbiome composition (Figure 3, Figure S11, S12).

**Figure 3.**
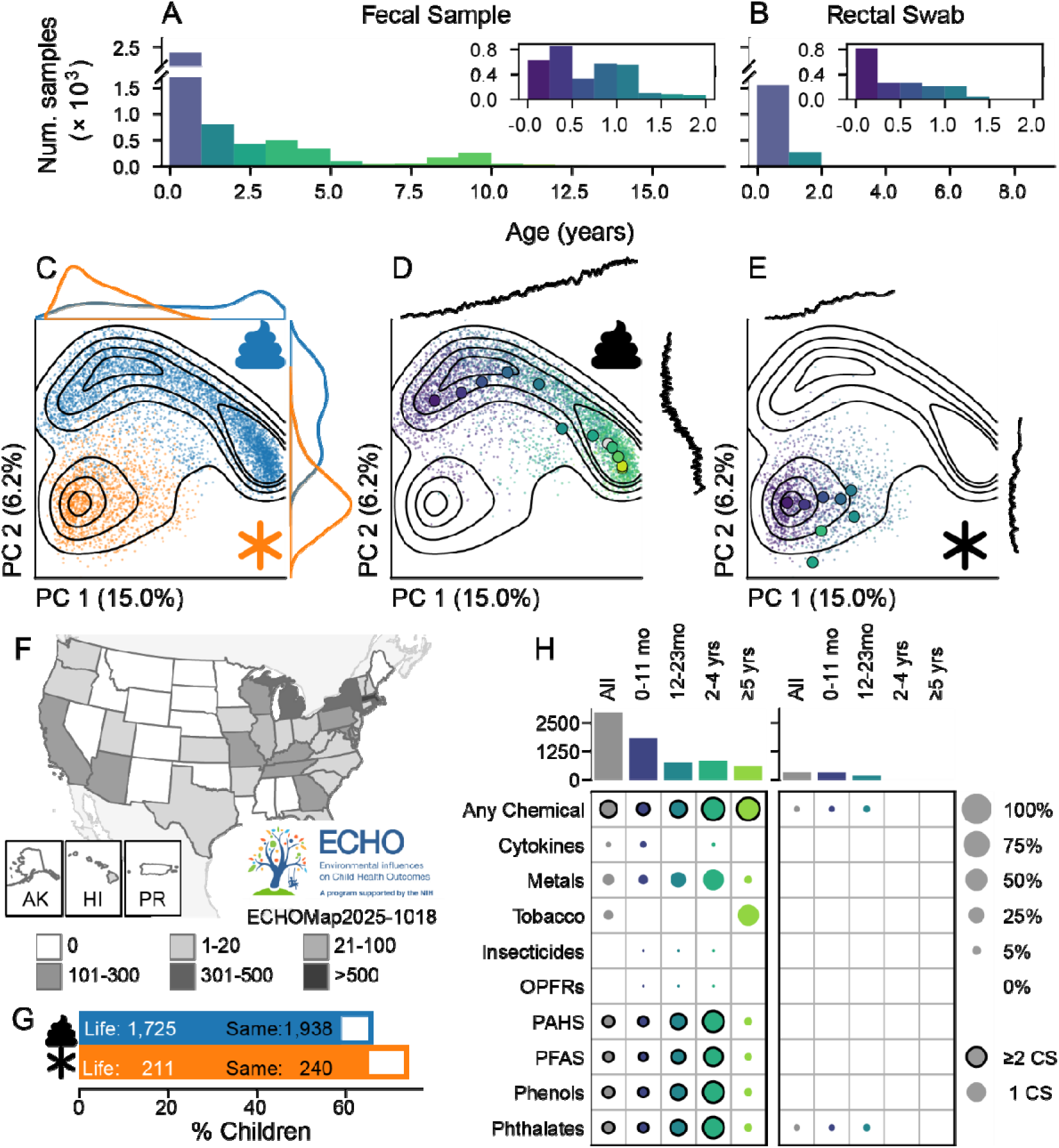
The child gut microbiome in ECHO. (A, B) Distribution of samples by age for participants with (A) fecal samples and (B) rectal swabs. (C-E) Principal coordinates projection of unweighted UniFrac distance. The lines reflect the density of samples used in the projection. Axes are colored by (C) sample type (blue - fecal; orange - rectal swab), (D) fecal samples by age, and (E) rectal swabs by age. Marginal trances on (D and E) show the average age at the same position; marginal axes for age share the same scale. (F) Map of the number of participants with gut or rectal swabs by state. (G) The proportion and number of participants with residential history over the child’s life (solid fill); and at the time of sample collection (white fill). (H) The proportion of children with chemical assays. The bar across the top indicates the number of participants in that age band; each cell gives the portion of participants with results for that assay. ECHO, Environmental influences on Child Health Outcomes.

**Table 2.** Participant characteristics for children with gut samples.

|  | All |  | Fecal sample |  | Rectal swab |  |
|---|---|---|---|---|---|---|
|  | Participants (n=3281) | Samples (n=7015) | Participants (n=2955) | Samples (n=5191) | Participants (n=326) | Samples (n=1824) |
| **Collection characteristics** |  |  |  |  |  |  |
| Number of samples collected |  |  |  |  |  |  |
| Median [IQR] | 2 [1, 2] |  | 2 [1, 2] |  | 3 [2, 4] |  |
| Range | 1-27 |  | 1-6 |  | 1-27 |  |
| Number of samples collected |  |  |  |  |  |  |
| 1 | 1473 (44.9%) |  | 1430 (48.4%) |  | 43 (13.2%) |  |
| 2 | 1096 (33.4%) |  | 1042 (35.3%) |  | 54 (16.6%) |  |
| 3 or more | 712 (21.7%) |  | 483 (16.3%) |  | 229 (70.2%) |  |
| Collection year<sup>a</sup> |  |  |  |  |  |  |
| Median | 2017 | 2016 | 2017 | 2017 | 2018 | 2015 |
| [IQR] | [2014, 2018] | [2014, 2018] | [2014, 2018] | [2014, 2018] | [2016, 2018] | [2014, 2018] |
| Range | 2011-2022 | 2011-2022 | 2011-2022 | 2011-2022 | 2011-2021 | 2011-2021 |
| Missing | 19 | 23 | 19 | 23 | 0 | 0 |
| Child age at collection (years)<sup>a</sup> |  |  |  |  |  |  |
| Mean (SD) | 2.10 (3.27) | 1.91 (2.64) | 2.32 (3.37) | 2.42 (2.89) | 0.11 (0.21) | 0.47 (0.44) |
| Median | 0.45 | 0.82 | 0.51 | 1.03 | 0.03 | 0.34 |
| [IQR] | [0.19, 2.31] | [0.30, 2.89] | [0.27, 2.96] | [0.40, 3.20] | [0.00, 0.09] | [0.09, 0.79] |
| Range | 0.00-16.99 | 0.00-16.99 | 0.01-16.99 | 0.01-16.99 | 0.00-1.39 | 0.00-3.35 |
| Missing | 20 | 26 | 20 | 26 | 0 | 0 |
| **Child demographics** |  |  |  |  |  |  |
| Child sex |  |  |  |  |  |  |
| Male | 1700 (51.8%) | 3622 (51.6%) | 1536 (52.0%) | 2732 (52.6%) | 164 (50.3%) | 890 (48.8%) |
| Female | 1581 (48.2%) | 3393 (48.4%) | 1419 (48.0%) | 2459 (47.4%) | 162 (49.7%) | 934 (51.2%) |
| Child racial group |  |  |  |  |  |  |
| White | 1741 (54.2%) | 3986 (57.7%) | 1527 (52.9%) | 2712 (53.1%) | 214 (66.0%) | 1274 (70.6%) |
| Black | 1013 (31.5%) | 1894 (27.4%) | 953 (33.0%) | 1594 (31.2%) | 60 (18.5%) | 300 (16.6%) |
| Asian | 82 (2.6%) | 166 (2.4%) | <80 (< 2.8%) | 159 (3.1%) | <5 (< 1.5%) | 7 (0.4%) |
| Multiple Races | 324 (10.1%) | 736 (10.6%) | 286 (9.9%) | 555 (10.9%) | 38 (11.7%) | 181 (10.0%) |
| Other Races<sup>b</sup> | 52 (1.6%) | 130 (1.9%) | 44 (1.5%) | <90 (< 1.8%) | 8 (2.5%) | <45 (< 2.5%) |
| Inconsistent | <5 | 22 | <5 | <5 | <5 | <25 |
| Missing | <65 | 81 | <70 | 81 | 0 | 0 |
| Maternal racial group |  |  |  |  |  |  |
| White | 1584 (56.2%) | 3823 (60.7%) | 1358 (54.2%) | 2471 (54.9%) | 226 (72.2%) | 1352 (75.3%) |
| Black | 987 (35.0%) | 1905 (30.2%) | 925 (36.9%) | 1579 (35.0%) | 62 (19.8%) | 326 (18.2%) |
| Asian | 112 (4.0%) | 250 (4.0%) | 99 (4.0%) | 209 (4.6%) | 13 (4.2%) | 41 (2.3%) |
| Multiple Races | 82 (2.9%) | 182 (2.9%) | 73 (2.9%) | 145 (3.2%) | 9 (2.9%) | 37 (2.1%) |
| Other Race<sup>b</sup> | 53 (1.9%) | 140 (2.2%) | <55 (< 2.2%) | 101 (2.2%) | <5 (< 1.6%) | 39 (2.2%) |
| Inconsistent | 31 | 36 | <35 | <35 | <5 | <5 |
| Missing | 432 | 679 | 420 | <655 | 12 | <30 |
| Maternal ethnicity |  |  |  |  |  |  |
| Non-Hispanic | 2539 (86.1%) | 5677 (87.5%) | 2238 (85.4%) | 3984 (85.4%) | 301 (92.3%) | 1693 (92.8%) |
| Hispanic | 409 (13.9%) | 814 (12.5%) | 384 (14.6%) | 683 (14.6%) | 25 (7.7%) | 131 (7.2%) |
| Inconsistent | 5 | 5 | 5 | 5 | 0 | 0 |
| Missing | 328 | 519 | 328 | 519 | 0 | 0 |
| Highest maternal education |  |  |  |  |  |  |
| No bachelor's degree | 1303 (48.1%) | 2663 (43.3%) | <1205 (<50.5%) | <2075 (<47.9%) | <105 (<32.5%) | <595 (<32.7%) |
| Bachelor's degree or higher | 1408 (51.9%) | 3491 (56.7%) | 1188 (49.7%) | 2263 (52.2%) | 220 (68.1%) | 1228 (67.5%) |
| Missing | 570 | 861 | <570 | <860 | <5 | <5 |
| **Birth characteristics and early-life diet** |  |  |  |  |  |  |
| Delivery mode |  |  |  |  |  |  |
| Vaginal | 2241 (69.3%) | 4565 (65.8%) | 2018 (69.3%) | 3601 (70.1%) | 223 (68.8%) | 964 (53.6%) |
| Cesarean | 994 (30.7%) | 2369 (34.2%) | <895 (<30.7%) | 1536 (29.9%) | <105 (<32.4%) | 833 (46.4%) |
| Missing | 46 | 81 | <45 | 54 | <5 | 27 |
| Preterm birth |  |  |  |  |  |  |
| Term (≥37 weeks) | 2938 (90.3%) | 5349 (78.4%) | 2701 (91.8%) | 4719 (91.3%) | 237 (76.2%) | 630 (38.2%) |
| Preterm (<37 weeks) | 315 (9.7%) | 1471 (21.6%) | 241 (8.2%) | 452 (8.7%) | 74 (23.8%) | 1019 (61.8%) |
| Missing | 28 | 195 | 13 | 20 | 15 | 175 |
| Birthweight for gestational age, intergrowth singleton standard<sup>c</sup> |  |  |  |  |  |  |
| Normal | 2477 (79.2%) | 4991 (79.6%) | 2251 (79.2%) | 4017 (79.9%) | 226 (79.0%) | 974 (78.2%) |
| Small | 202 (6.5%) | 420 (6.7%) | 183 (6.4%) | 299 (5.9%) | 19 (6.6%) | 121 (9.7%) |
| Large | 450 (14.4%) | 863 (13.8%) | 409 (14.4%) | 713 (14.2%) | 41 (14.3%) | 150 (12.0%) |
| Missing | 152 | 741 | 112 | 162 | 40 | 579 |
| NICU admission |  |  |  |  |  |  |
| No | 701 (79.2%) | 1133 (45.6%) | 701 (88.0%) | 1133 (87.4%) | 0 (0.0%) | 0 (0.0%) |
| Yes | 184 (20.8%) | 1354 (54.4%) | 96 (12.0%) | 163 (12.6%) | 88 (100.0%) | 1191 (100.0%) |
| Missing | 2396 | 4528 | 2158 | 3895 | 238 | 633 |
| Ever breastfed |  |  |  |  |  |  |
| No | 337 (12.9%) | 497 (8.9%) | <320 (<13.9%) | <410 (<10.9%) | <20 (< 6.2%) | <90 (< 4.9%) |
| Yes | 2284 (87.1%) | 5080 (91.1%) | 1979 (86.2%) | 3347 (89.1%) | 305 (94.1%) | 1733 (95.2%) |
| Missing | 660 | 1438 | <660 | <1440 | <5 | <5 |
| Ever formula fed |  |  |  |  |  |  |
| No | 446 (23.2%) | 892 (22.6%) | 323 (19.9%) | 546 (22.4%) | 123 (41.0%) | 346 (23.0%) |
| Yes | 1474 (76.8%) | 3050 (77.4%) | 1297 (80.1%) | 1893 (77.6%) | 177 (59.0%) | 1157 (77.0%) |
| Missing | 1361 | 3073 | 1335 | 2752 | 26 | 321 |
| Breastfed at 3 months<sup>d</sup> |  |  |  |  |  |  |
| No | 917 (37.7%) | 1694 (32.0%) | <815 (<38.7%) | <1120 (<32.3%) | <110 (<34.0%) | <580 (<31.9%) |
| Yes | 1515 (62.3%) | 3596 (68.0%) | 1296 (61.5%) | 2352 (67.8%) | 219 (67.6%) | 1244 (68.3%) |
| Missing | 849 | 1725 | <850 | <1725 | <5 | <5 |
| Breastfed at 6 months<sup>d</sup> |  |  |  |  |  |  |
| No | 1290 (52.8%) | 2650 (49.9%) | <1140 (<53.7%) | 1688 (48.4%) | <155 (<47.8%) | 962 (52.8%) |
| Yes | 1155 (47.2%) | 2659 (50.1%) | 983 (46.3%) | <1805 (<51.7%) | 172 (53.1%) | <860 (<47.2%) |
| Missing | 836 | 1706 | <835 | <1705 | <5 | <5 |
| **Outcomes** |  |  |  |  |  |  |
| Asthma diagnosis<sup>e</sup> |  |  |  |  |  |  |
| No | 1287 (75.3%) | 3455 (77.7%) | 1030 (71.8%) | 2040 (73.9%) | 257 (93.8%) | 1415 (83.9%) |
| Yes | 422 (24.7%) | 993 (22.3%) | 405 (28.2%) | 721 (26.1%) | 17 (6.2%) | 272 (16.1%) |
| Missing | 1572 | 2567 | 1520 | 2430 | 52 | 137 |
| Asthma diagnosis relative to sample<sup>e</sup> |  |  |  |  |  |  |
| No diagnosis | 1258 (75.2%) | 3455 (77.8%) | 1029 (72.1%) | 2040 (74.1%) | 229 (93.1%) | 1415 (83.9%) |
| Sample before diagnosis | 393 (23.5%) | 934 (21.0%) | 376 (26.3%) | 662 (24.0%) | 17 (6.9%) | 272 (16.1%) |
| Sample after diagnosis | 22 (1.3%) | 52 (1.2%) | 22 (1.5%) | 52 (1.9%) | 0 (0.0%) | 0 (0.0%) |
| Missing | 1608 | 2574 | 1528 | 2437 | 80 | 137 |
| Any food allergy diagnosis |  |  |  |  |  |  |
| No | 223 (43.0%) | 421 (40.1%) | <225 (<46.5%) | <420 (<48.1%) | <5 (<14.3%) | <5 (< 2.8%) |
| Yes | 296 (57.0%) | 628 (59.9%) | <265 (<54.8%) | <460 (<52.7%) | <35 (<100.0%) | <175 (<99.4%) |
| Missing | 2762 | 5966 | 2471 | 4318 | 291 | 1648 |
| Food allergy diagnosis relative to sample |  |  |  |  |  |  |
| No diagnosis | 213 (42.9%) | 421 (40.3%) | <215 (<46.0%) | <420 (<48.4%) | <5 (<16.7%) | <5 (< 2.8%) |
| Sample before diagnosis | 195 (39.2%) | 481 (46.1%) | 175 (37.5%) | 325 (37.4%) | 20 (66.7%) | 156 (88.6%) |
| Sample after diagnosis | 41 (8.2%) | 142 (13.6%) | <40 (< 8.6%) | <130 (<15.0%) | <5 (<16.7%) | <20 (<11.4%) |
| Sampled before and after | 48 (9.7%) |  | 42 (9.0%) |  | 6 (20.0%) |  |
| Missing | 2784 | 5971 | 2488 | 4323 | 296 | 1648 |
| Eczema report |  |  |  |  |  |  |
| No diagnosis | 784 (32.8%) | 1948 (35.7%) | 622 (30.0%) | 1052 (28.4%) | 162 (51.1%) | 896 (51.1%) |
| Self report | 566 (23.7%) | 1213 (22.2%) | 505 (24.3%) | 927 (25.0%) | 61 (19.2%) | 286 (16.3%) |
| Medical diagnosis | 1043 (43.6%) | 2295 (42.1%) | 949 (45.7%) | 1724 (46.6%) | 94 (29.7%) | 571 (32.6%) |
| Missing | 888 | 1559 | 879 | 1488 | 9 | 71 |
| Eczema report relative to sample |  |  |  |  |  |  |
| No diagnosis | 753 (33.5%) | 1948 (36.2%) | 618 (31.3%) | 1052 (29.0%) | 135 (49.6%) | 896 (51.1%) |
| Sample before diagnosis | 445 (19.8%) | 1113 (20.7%) | 439 (22.2%) | 1034 (28.5%) | 6 (2.2%) | 79 (4.5%) |
| Sample after diagnosis | 778 (34.6%) | 2321 (43.1%) | 695 (35.2%) | 1543 (42.5%) | 83 (30.5%) | 778 (44.4%) |
| Sampled before and after | 273 (12.1%) |  | 225 (11.4%) |  | 48 (17.6%) |  |
| Missing | 1032 | 1633 | 978 | 1562 | 54 | 71 |
| Autism diagnosis |  |  |  |  |  |  |
| No | 1817 (96.5%) | 4774 (97.1%) | 1609 (96.2%) | 3252 (96.6%) | 208 (98.6%) | 1522 (98.2%) |
| Yes | 66 (3.5%) | 144 (2.9%) | <65 (< 3.9%) | 116 (3.4%) | <5 (< 2.4%) | 28 (1.8%) |
| Missing | 1398 | 2097 | <1285 | 1823 | <120 | 274 |
| ADHD diagnosis |  |  |  |  |  |  |
| No | 1735 (92.5%) | 4407 (90.0%) | 1540 (92.5%) | 3082 (91.9%) | 195 (92.9%) | 1325 (85.6%) |
| Yes | 140 (7.5%) | 492 (10.0%) | 125 (7.5%) | 270 (8.1%) | 15 (7.1%) | 222 (14.4%) |
| Missing | 1406 | 2116 | 1290 | 1839 | 116 | 277 |
| Ever obese<sup>f</sup> |  |  |  |  |  |  |
| No | 1689 (79.2%) | 4200 (81.9%) | 1484 (78.0%) | 2810 (78.9%) | 205 (89.1%) | 1390 (88.6%) |
| Yes | 444 (20.8%) | 930 (18.1%) | 419 (22.0%) | 751 (21.1%) | 25 (10.9%) | 179 (11.4%) |
| Missing | 1148 | 1885 | 1052 | 1630 | 96 | 255 |
| Obesity relative to sample<sup>f</sup> |  |  |  |  |  |  |
| No diagnosis | 1658 (79.2%) | 4200 (82.0%) | 1454 (78.0%) | 2810 (79.1%) | 204 (89.1%) | 1390 (88.6%) |
| Sample before dx | 435 (20.8%) | 922 (18.0%) | 410 (22.0%) | 743 (20.9%) | 25 (10.9%) | 179 (11.4%) |
| Missing | 1148 | 1885 | 1052 | 1630 | 96 | 255 |

<sup>a</sup>Value for participant reflects the first sample collected.
<sup>b</sup>"Other Races" includes participants who did not identify as White, Black, Asian, or Multiple Races as well as participants who identified as American Indian or Alaskan Native and participants who identified and Native Hawaiian or other Pacific Islander.
<sup>c</sup>This measure excludes non-singleton pregnancies; children of multiple gestation are listed as "missing."
<sup>d</sup>Lifetime duration including participants who were sampled before the age of completion.
<sup>e</sup>Asthma diagnosis was made after age 4; children followed fewer than 4 years cannot have asthma diagnoses.
<sup>f</sup>Obesity was only determined for children with BMI measurements at age 2 years or older; children followed fewer than 2 years cannot have obesity diagnoses.
ADHD, attention-deficit/hyperactivity disorder; IQR, interquartile range; NICU, neonatal intensive care unit; SD, standard deviation

Children with gut samples lived in 37 U.S. states (Figure 3F). There were 2,178 participants (4,781 samples) with residential history geocoded to the Census tract level or better at the time of microbiome sample collection, including 1,938 (65.6%) participants with fecal samples and 240 (73.6%) with rectal swabs (Figure 3G). In total, 1,068 children (2,200 samples) had at least one linked chemical measurement; these were primarily in children with fecal samples (Figure 3H). Genetic profiling was available for 1,377 children with gut samples (1,317 with fecal; 60 with rectal swabs); epigenetic sequencing had been performed for 386 participants, all of whom had fecal samples. Most children who had epigenetics also had genetic data (n=327); there were few children with any other combination of omic measurements.

### Child Upper Airway Samples

Nasal or nasopharyngeal samples were collected at 10 cohort sites; there were 6,945 high-quality samples from 2,956 children. The dataset included multiple children from 27 multi-gestational pregnancies and 175 families with children from multiple pregnancies. There were 404 children from 201 families with samples from multiple children. Most children with upper airway samples were sampled once (n=2,004, 70.2%); among the 503 children (29.8%) who were sampled more than once, children contributed a median of 3 samples (IQR 2, 9; range 2-29). Participant characteristics, outcomes, and sampling characteristics for children with upper airway samples are described in Table 3. The community composition is shown in Figures 4 and S14. Nasal samples were collected from children across 26 states; 1,071 participants (1,272 samples) could be geocoded with Census tract–level or finer accuracy at the time of sample collection. Chemical exposure data were available for 423 participants (14.8%); tobacco metabolites were the most common metabolite, available for 454 (12.4%) children with upper airway samples (Figure 4C). A total of 1,435 children with upper airway samples were genotyped. Epigenetic profiling was conducted on 1,093 children, including 995 participants with epigenetic profiling from nasal tissue, and 619 participants had metabolic measurements (Figure 6E).

**Table 3.**
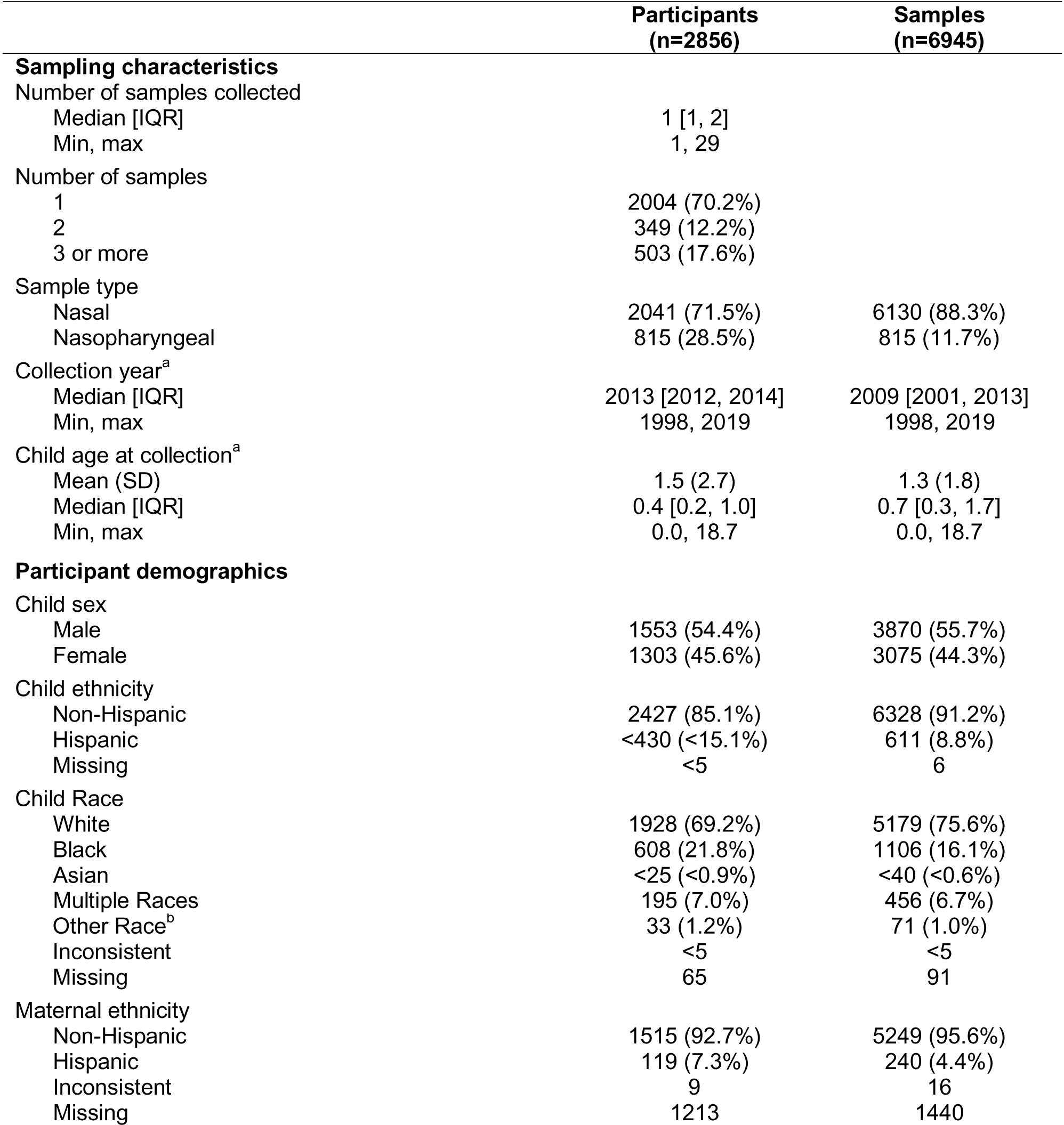
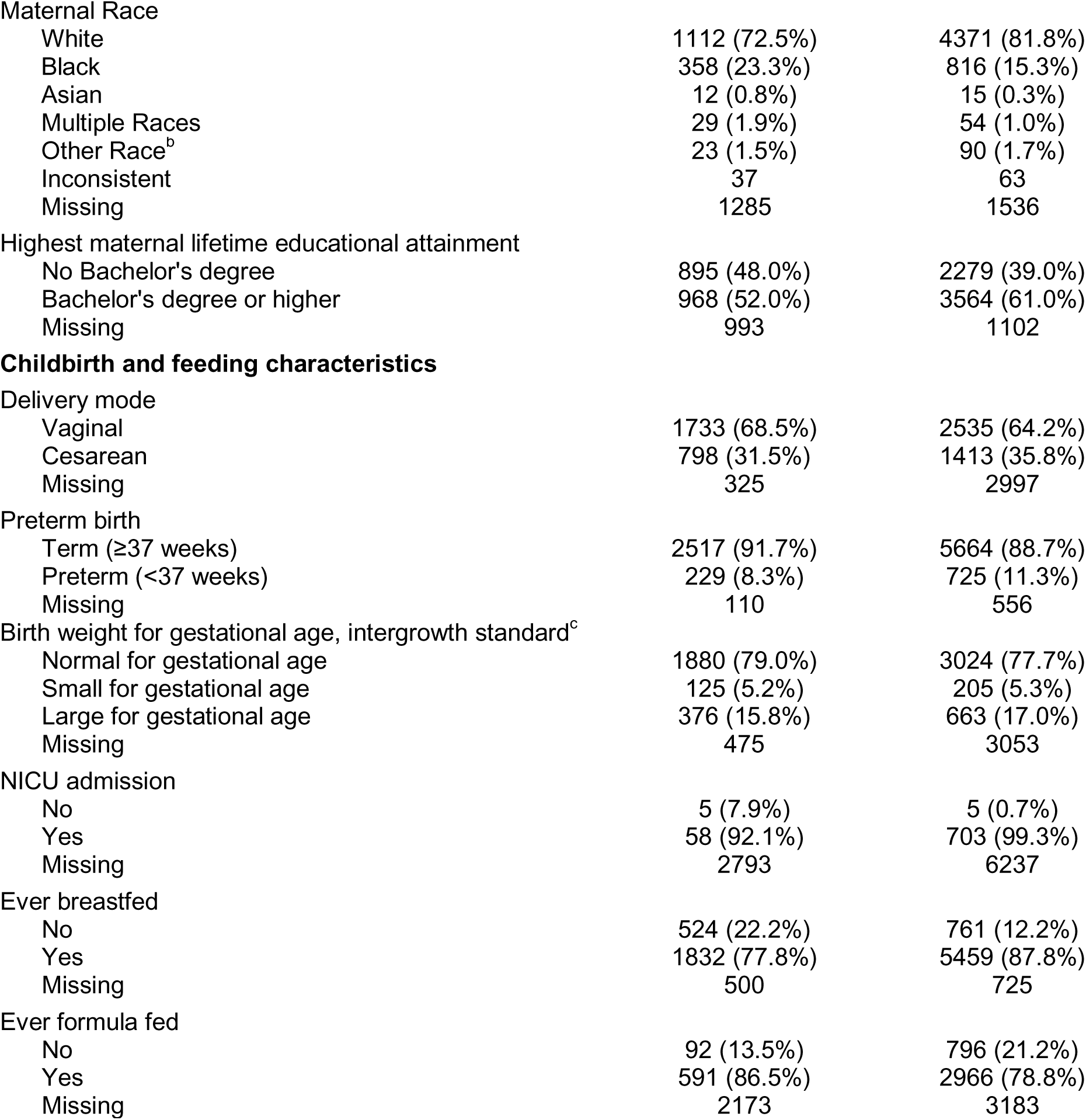
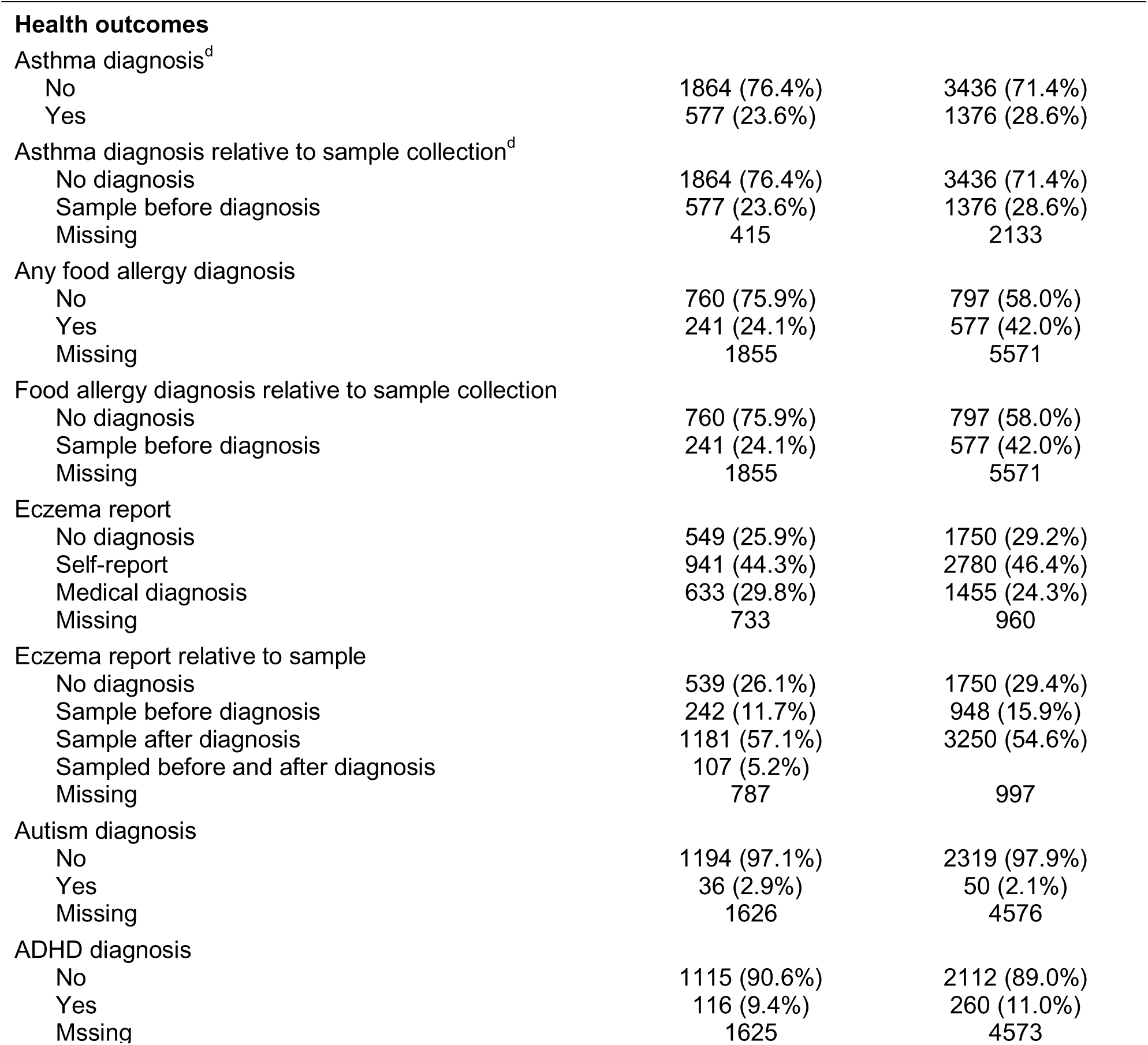
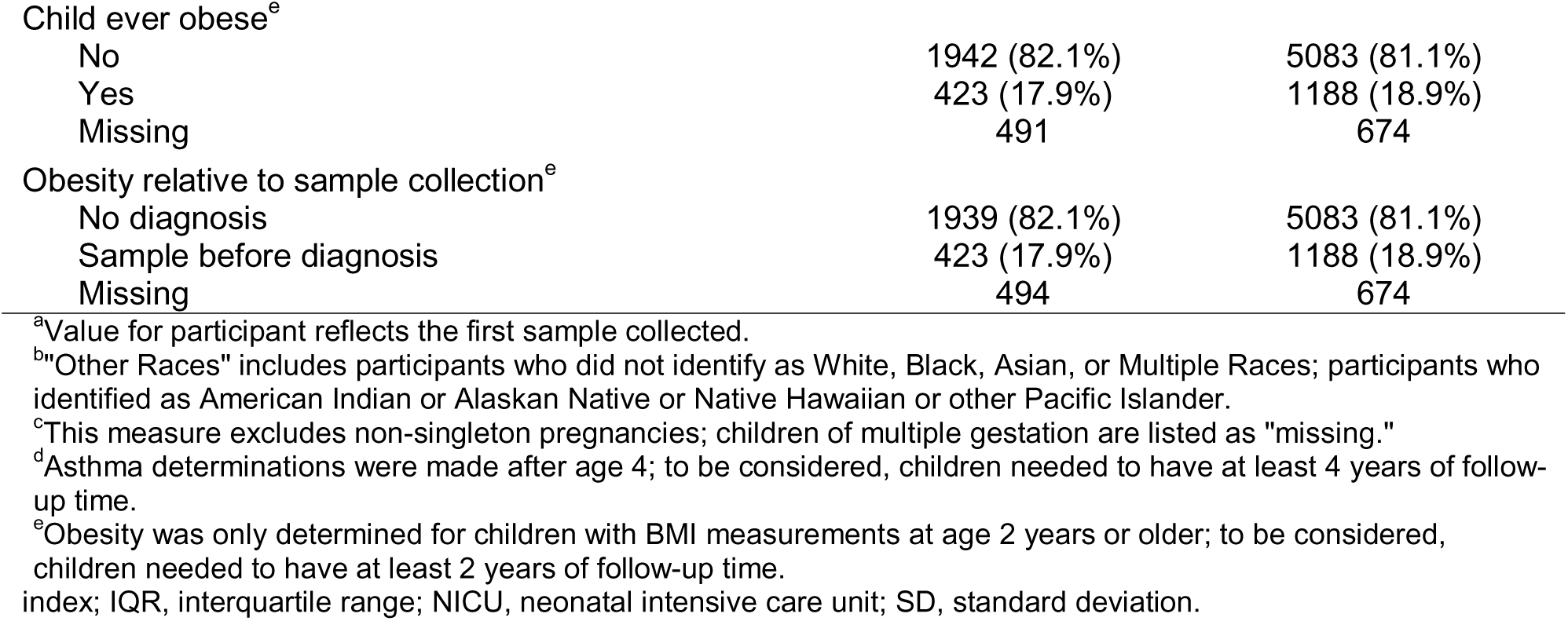
Participant characteristics for children with upper airway samples.

|  | Participants<br>(n=2856) | Samples<br>(n=6945) |
|---|---|---|
| **Sampling characteristics** |  |  |
| Number of samples collected |  |  |
| Median [IQR] | 1 [1, 2] |  |
| Min, max | 1, 29 |  |
| Number of samples |  |  |
| 1 | 2004 (70.2%) |  |
| 2 | 349 (12.2%) |  |
| 3 or more | 503 (17.6%) |  |
| Sample type |  |  |
| Nasal | 2041 (71.5%) | 6130 (88.3%) |
| Nasopharyngeal | 815 (28.5%) | 815 (11.7%) |
| Collection year<sup>a</sup> |  |  |
| Median [IQR] | 2013 [2012, 2014] | 2009 [2001, 2013] |
| Min, max | 1998, 2019 | 1998, 2019 |
| Child age at collection<sup>a</sup> |  |  |
| Mean (SD) | 1.5 (2.7) | 1.3 (1.8) |
| Median [IQR] | 0.4 [0.2, 1.0] | 0.7 [0.3, 1.7] |
| Min, max | 0.0, 18.7 | 0.0, 18.7 |
| **Participant demographics** |  |  |
| Child sex |  |  |
| Male | 1553 (54.4%) | 3870 (55.7%) |
| Female | 1303 (45.6%) | 3075 (44.3%) |
| Child ethnicity |  |  |
| Non-Hispanic | 2427 (85.1%) | 6328 (91.2%) |
| Hispanic | <430 (<15.1%) | 611 (8.8%) |
| Missing | <5 | 6 |
| Child Race |  |  |
| White | 1928 (69.2%) | 5179 (75.6%) |
| Black | 608 (21.8%) | 1106 (16.1%) |
| Asian | <25 (<0.9%) | <40 (<0.6%) |
| Multiple Races | 195 (7.0%) | 456 (6.7%) |
| Other Race<sup>b</sup> | 33 (1.2%) | 71 (1.0%) |
| Inconsistent | <5 | <5 |
| Missing | 65 | 91 |
| Maternal ethnicity |  |  |
| Non-Hispanic | 1515 (92.7%) | 5249 (95.6%) |
| Hispanic | 119 (7.3%) | 240 (4.4%) |
| Inconsistent | 9 | 16 |
| Missing | 1213 | 1440 |
| Maternal Race |  |  |
| White | 1112 (72.5%) | 4371 (81.8%) |
| Black | 358 (23.3%) | 816 (15.3%) |
| Asian | 12 (0.8%) | 15 (0.3%) |
| Multiple Races | 29 (1.9%) | 54 (1.0%) |
| Other Race<sup>b</sup> | 23 (1.5%) | 90 (1.7%) |
| Inconsistent | 37 | 63 |
| Missing | 1285 | 1536 |
| Highest maternal lifetime educational attainment |  |  |
| No Bachelor's degree | 895 (48.0%) | 2279 (39.0%) |
| Bachelor's degree or higher | 968 (52.0%) | 3564 (61.0%) |
| Missing | 993 | 1102 |
| **Childbirth and feeding characteristics** |  |  |
| Delivery mode |  |  |
| Vaginal | 1733 (68.5%) | 2535 (64.2%) |
| Cesarean | 798 (31.5%) | 1413 (35.8%) |
| Missing | 325 | 2997 |
| Preterm birth |  |  |
| Term (≥37 weeks) | 2517 (91.7%) | 5664 (88.7%) |
| Preterm (<37 weeks) | 229 (8.3%) | 725 (11.3%) |
| Missing | 110 | 556 |
| Birth weight for gestational age, intergrowth standard<sup>c</sup> |  |  |
| Normal for gestational age | 1880 (79.0%) | 3024 (77.7%) |
| Small for gestational age | 125 (5.2%) | 205 (5.3%) |
| Large for gestational age | 376 (15.8%) | 663 (17.0%) |
| Missing | 475 | 3053 |
| NICU admission |  |  |
| No | 5 (7.9%) | 5 (0.7%) |
| Yes | 58 (92.1%) | 703 (99.3%) |
| Missing | 2793 | 6237 |
| Ever breastfed |  |  |
| No | 524 (22.2%) | 761 (12.2%) |
| Yes | 1832 (77.8%) | 5459 (87.8%) |
| Missing | 500 | 725 |
| Ever formula fed |  |  |
| No | 92 (13.5%) | 796 (21.2%) |
| Yes | 591 (86.5%) | 2966 (78.8%) |
| Missing | 2173 | 3183 |
| **Health outcomes** |  |  |
| Asthma diagnosis<sup>d</sup> |  |  |
| No | 1864 (76.4%) | 3436 (71.4%) |
| Yes | 577 (23.6%) | 1376 (28.6%) |
| Asthma diagnosis relative to sample collection<sup>d</sup> |  |  |
| No diagnosis | 1864 (76.4%) | 3436 (71.4%) |
| Sample before diagnosis | 577 (23.6%) | 1376 (28.6%) |
| Missing | 415 | 2133 |
| Any food allergy diagnosis |  |  |
| No | 760 (75.9%) | 797 (58.0%) |
| Yes | 241 (24.1%) | 577 (42.0%) |
| Missing | 1855 | 5571 |
| Food allergy diagnosis relative to sample collection |  |  |
| No diagnosis | 760 (75.9%) | 797 (58.0%) |
| Sample before diagnosis | 241 (24.1%) | 577 (42.0%) |
| Missing | 1855 | 5571 |
| Eczema report |  |  |
| No diagnosis | 549 (25.9%) | 1750 (29.2%) |
| Self-report | 941 (44.3%) | 2780 (46.4%) |
| Medical diagnosis | 633 (29.8%) | 1455 (24.3%) |
| Missing | 733 | 960 |
| Eczema report relative to sample |  |  |
| No diagnosis | 539 (26.1%) | 1750 (29.4%) |
| Sample before diagnosis | 242 (11.7%) | 948 (15.9%) |
| Sample after diagnosis | 1181 (57.1%) | 3250 (54.6%) |
| Sampled before and after diagnosis | 107 (5.2%) |  |
| Missing | 787 | 997 |
| Autism diagnosis |  |  |
| No | 1194 (97.1%) | 2319 (97.9%) |
| Yes | 36 (2.9%) | 50 (2.1%) |
| Missing | 1626 | 4576 |
| ADHD diagnosis |  |  |
| No | 1115 (90.6%) | 2112 (89.0%) |
| Yes | 116 (9.4%) | 260 (11.0%) |
| Mssing | 1625 | 4573 |
| Child ever obese<sup>e</sup> |  |  |
| No | 1942 (82.1%) | 5083 (81.1%) |
| Yes | 423 (17.9%) | 1188 (18.9%) |
| Missing | 491 | 674 |
| Obesity relative to sample collection<sup>e</sup> |  |  |
| No diagnosis | 1939 (82.1%) | 5083 (81.1%) |
| Sample before diagnosis | 423 (17.9%) | 1188 (18.9%) |
| Missing | 494 | 674 |

<sup>a</sup>Value for participant reflects the first sample collected.
<sup>b</sup>"Other Races" includes participants who did not identify as White, Black, Asian, or Multiple Races; participants who identified as American Indian or Alaskan Native or Native Hawaiian or other Pacific Islander.
<sup>c</sup>This measure excludes non-singleton pregnancies; children of multiple gestation are listed as "missing."
<sup>d</sup>Asthma determinations were made after age 4; to be considered, children needed to have at least 4 years of follow-up time.
<sup>e</sup>Obesity was only determined for children with BMI measurements at age 2 years or older; to be considered, children needed to have at least 2 years of follow-up time.
index; IQR, interquartile range; NICU, neonatal intensive care unit; SD, standard deviation.

**Figure 4.**
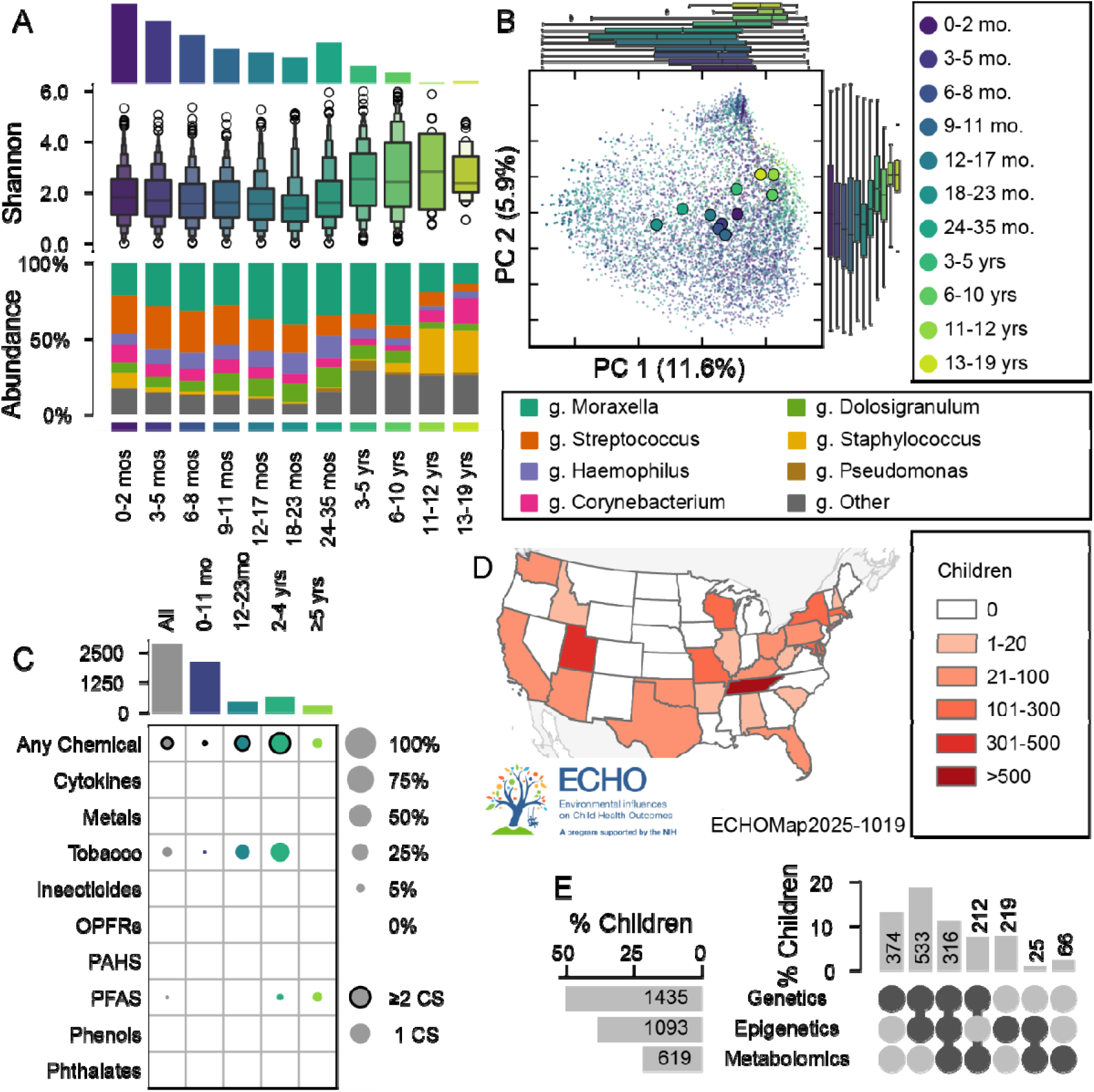
Microbiome composition and environmental data for children with upper airway samples. (A) Age group, alpha diversity, and taxonomic composition. The number of participants is shown in the upper bar. (B) PCoA projection of Jaccard distance, colored by age. Marginal plots show the distribution of participants in that age group over time. (C) The proportion of children with chemical assays. The bar across the top indicates the number of participants in that age band; each cell gives the portion of participants who have a measure in that age band and for that metabolite. Outlined cells indicate that children from two or more cohort sites have the measurement. (D) State of residence for children with upper airway microbiome samples. No samples were collected from children in Alaska, Hawaii, or Puerto Rico. (E) Upset plot shows available omics for children with upper airway samples. The bar plot to the left in each row indicates the total percentage and number of participants who have a given omics measurement; the vertical bars indicate the number and percentage of children with overlaps between omics; overlap combinations are shown in the lower right with darker points indicating the intersection.

### Example Analysis: The effect of diet on the infant fecal microbiome

To demonstrate the utility of our harmonized data, we analyzed the effect of child age and breastmilk consumption on the infant microbiome. We performed a complete case analysis of 828 term children from seven cohort sites with high-quality fecal samples collected between one and five months of age (Figure 5A, Table S5). We modeled the relationship between diet and the fecal microbiome, adjusted for cohort site, age, sex, and delivery mode. Compared to exclusively breastfed infants, infants fed a mixed diet or no breastmilk had higher Shannon diversity: 0.34 (95% confidence interval [CI] 0.22, 0.46) and 0.84 (95% CI 0.71, 0.96), respectively (Figure 5B,D, Table S6). Breastmilk consumption (None, Any, Exclusive) was the primary biological predictor of microbiome composition measured by unweighted UniFrac distance, explaining more than twice the variation as the next most explanatory factor (Figure 5C,E). There were more than 40 differentially abundant genera in infants with no breastmilk compared to those exclusively breastfed (Figure 5F, Table S7). The directionality of three taxa was replicated in the “any” breastmilk group.

**Figure 5.**
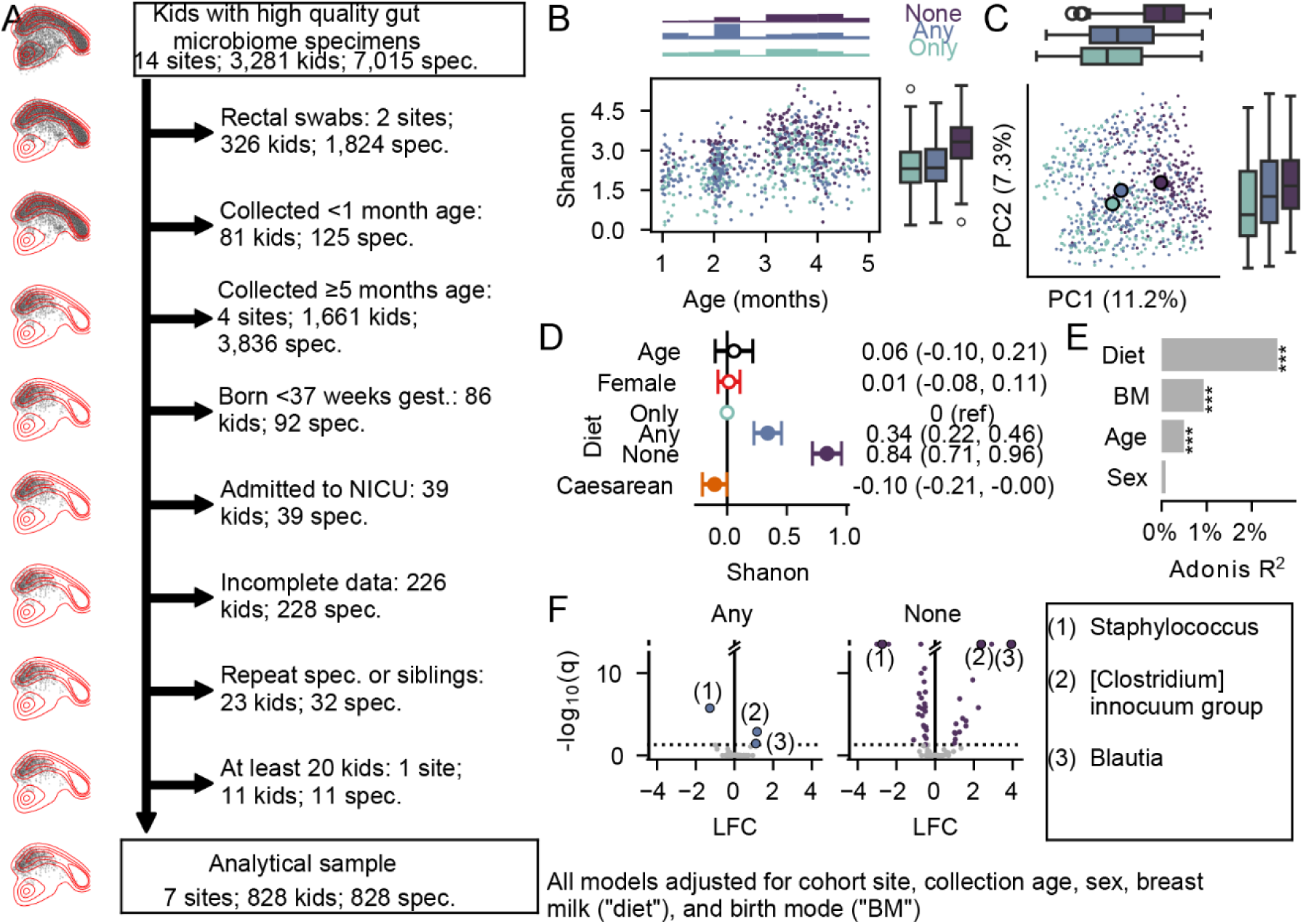
Diet is associated with the fecal microbiome structure of infants. (A) Participant selection from all gut samples (top) to the analytical sample shown here (bottom). Principal coordinates plots (PCoA) on the left show the full original dataset (red) with the retained samples highlighted as points. (D-F) Modeling results for (D) alpha diversity, (E) beta diversity, and (F) untargeted differential abundance; all adjusted for cohort site as a fixed effect and modeling the age, sex (female vs male), diet, and delivery mode (cesarian vs vaginal). (D) Forest plot, showing the change in Shannon diversity per predictor. (E) Adonis modeling results comparing the variation explained by each predictor. Values are ranked based on their variation explained. Permutative p-values calculated from 999 permutations: *** p = 0.001; **p < 0.01, *p < 0.05. (F) Differentially abundant taxa for Any breastmilk and No breastmilk vs Only breastmilk. In both cases, negative values indicate higher relative abundance in infants with only breastmilk and positive values indicate higher values in any or no breastmilk, respectively. Log q values were trimmed at 12; values greater than 12 are shown above the cut axis. Numbered taxa were statistically significant in both groups of infants; direction is replicated in both cases.

## DISCUSSION

The ECHO Cohort has generated and harmonized microbiome data for 15,509 unique, high-quality samples collected from 1,086 pregnancies and 5,701 children across 21 ECHO cohort sites. Samples were collected from the vagina and gut of pregnant participants, and from the gut and upper airway of children. As previously reported, body site was a major determinant of microbiome community composition (106, 107). We were able to link microbiome samples with diverse personal factors, including pregnancy characteristics, demographics, and health outcomes. High-quality residential history information was available for more than 1,000 children with upper airway data, almost 2,000 children with gut data, and more than 60% of pregnancies with microbiome data. Over one-third of children with gut samples had at least one linked chemical measurement while half of the children with upper airway samples had linked genetic data. While cohort-level technical effects are present in the data, we replicated previously reported associations between the microbiome community and both breastfeeding and delivery mode in infants, reflecting the utility of this dataset in child health research (108, 109).

The ECHO Cohort offers several unique opportunities for advancing child health through the integrated study of the microbiome of diverse American children with rich metadata. The Cohort is geographically diverse, with participants recruited from 40 U.S. states. Pregnancy and child gut samples were collected between 2011 and 2022, and upper airway sample collections go back to 2000. A combined analysis of these data will allow for the characterization of geographic and temporal diversity in the microbiome to understand which influences are localized, temporally varying, or generalizable. Our high-quality geocoded data uniquely enable the study of the relationships between the microbiome and area-level exposures across heterogeneous environments in the U.S. Previous ECHO studies have analyzed the associations of area-level exposures, including environmental and social stress (110); drinking water contaminants, including arsenic (111); and airborne lead (112). Extensions of this research may include the consideration of the microbiome as a mediator in the exposure-outcome relationship.

Furthermore, ECHO is one of the few multi-center studies with extant data where harmonization is conducted centrally. Metadata availability and harmonization for re-use is often cited as a barrier to the cross-study comparison of microbiome data (113–115). The ECHO data harmonization model of initial, expert-guided harmonization, followed by the transition to a common data collection protocol in combination with primary microbiome data, facilitates comparisons of both existing measurements not previously directly linked to microbiome data and the long-term follow-up of participants (94, 116). This approach provides more power than what may be achieved in an individual study and more reliable metadata than what may be achieved in individual re-analysis studies (115).

However, microbiome analyses in the ECHO Cohort present several challenges. First, as observed in other studies (102–104), technical decisions regarding study design, sample collection and storage, and DNA extraction, amplification, and sequencing have large effects on the observed microbiome community in our data. Few bioinformatic methods are available to remove these biases; while these could be applied in later analyses, we chose to present the data here without re-normalization. Alpha (within-sample) diversity and differential abundance (taxa-specific comparisons) can be compared using random effects modeling or meta-analysis; beta diversity, which relies on direct sample-to-sample comparison and measures, such as maturation, may be more challenging to meta-analyze. Second, our data are based on 16S rRNA sequencing. While 16S sequencing provides lower taxonomic resolution and does not allow either functional annotation or strain tracking, it remains important for microbiome research and the re-use of data. Amplicon sequencing and paired metagenomic analysis provide similar community-level insights and can help guide more focused research questions (117). While these challenges are difficult to address in our current data, ECHO has banked microbiome samples collected under a common protocol and has continued collection with new participants (116). New techniques allow the integration of 16s rRNA sequencing data and metagenomic results (118); the possibility of sequencing these banked samples in the future presents an opportunity to substantially increase our analytical power while minimizing technical biases (102–104).

## CONCLUSION

Using extant data, the ECHO Cohort harmonized over 15,000 high-quality samples from more than 5,700 children and 1,086 pregnancies. The child samples are primarily collected in the critical first two years of development. The samples are linked to rich metadata, including participant-specific characteristics, chemical measures, and geospatial linkage. ECHO provides an important resource for understanding how environmental exposures influence the microbiome, and ultimately, the health of American children.

## Data Availability

Select de-identified data from the ECHO Program are available through NICHDs Data and Specimen Hub (DASH). Raw sequencing data availability for individual studies is described in Table S1.

## Disclaimer

The content is solely the responsibility of the authors and does not necessarily represent the official views of the National Institutes of Health.

## Author Contributions

The study was conceptualized by JWD, NTM, SSC, ES, KLM, VKC, and AMK. Local sequencing data was contributed by SSC, VCK, SRD, JAE, JEG, TVH, DJJ, JLA, TGO, SC, and CRS. Data harmonization methodology was determined by JWD, NTM, AKM, and ES; data was harmonized by JWD and SX. JWD and SX both wrote software to support the project. GPP provided and supported computational resources. JWD analyzed the data with help from CW. The original manuscript was drafted by JWD, KLM, and MP with critical edits from AKM, SX, NTM, SSC, VCK, and CRS. AKM supervised the project; JWD, ZL, and AKM were responsible for administration. TGO, PAB, MRK, ZZ, LDL, TVH, CAC, JEG, JLA, and DJJ secured funding. All authors reviewed and approved the final manuscript.

## Code and Data Availability

Select de-identified data from the ECHO Program are available through <u>NICHD’s Data</u> <u>and Specimen Hub (DASH)</u>. Information on study data not available on DASH, such as some Indigenous datasets, can be found on the <u>ECHO study DASH webpage</u>. SRA, Import, and dbGap accessions for raw sequencing datasets, where available, can be found in Supplemental Table 1. Code and data for this project are indexed under ECHO project number EC0722; analysis is based on the 20231031 data lock.

## Acknowledgments

The authors wish to thank our ECHO Colleagues; the medical, nursing, and program staff; and the children and families participating in the ECHO cohort. Special thanks to Ann Lindley Gill, Jean M Kerver, Siyuan Ma, Ashley F. Sullivan. The authors wish to acknowledge Samantha Fye and Any Dresen; the Marshfield Clinic Research Institute acknowledges research coordinators Tara Johnson, Terry Foss, Elizabeth Armagost, and Vicki Moon as well as lab personnel Tamara Kronenwetter Koepel, Erin Higdon, Wayne Frome, Carrie Marcis, and Sarah Kohn for their efforts in collecting and processing samples; and the coordinators and lab personnel both past and present in the CREW consortium who made sample collection possible. This research could not have been done without the collaboration of Hurley Medical Center and Hurley Residency Clinic in Flint, MI, Hutzel Medical Center, DMC Center for Obstetrics and Gynecology, and University Health Center in Detroit, MI Munson Hospital and Grand Traverse Women’s Clinic in Traverse City, MI, University of Michigan Hospital, Von Voigtlander Women’s Center, West Ann Arbor Health Center, and Obstetrics and Gynecology at Briarwood Center in Ann Arbor, MI, St. John’s Providence Park, Metro Partners in Women’s Health, and Women’s Health Consultants in Novi, MI, St. Joseph Mercy Hospital, IHA Canton Obstetrics and Gynecology, and IHA Domino Farms in Ann Arbor, MI, Sinai Grace Hospital, Sinai Grace OB-GYN Women’s Health Centers, Northwest Women’s Care, and DMC Northwest Women’s Care in Detroit, MI, Beaumont Dearborn Hospital and Oakwood OB-GYN Associates in Dearborn, MI, Covenant Hospital, Central Michigan University Health in Saginaw, MI, SHMG OB/GYN, Grand Rapids Women’s Health, and Spectrum Health in Grand Rapids, MI, Blue Water OB/GYN, NorthPointe OB/GYN, and McLaren Port Huron Hospital in Port Huron, MI. The authors wish to thank the Alkek Center for Metagenomics and Microbiome Research, Douglas W Fadrosh, Kole V Lynch, Kathryn E McCauley, and Susan V Lynch at the Lynch Lab at the University of California at San Francisco; Kathryn Holt at Department of Infection Biology, London School of Hygiene and Tropical Medicine; Michael Inouye, University of Melbourne and Cambridge University; and Michael Tang, University of Cambridge for their support in sequencing and analysis.

