## Supplementary material for "Harmonization of microbiome data in the Environmental influences on Child Health Outcomes (ECHO) Cohort to analyze environmental influences on the child microbiome": File S1

### Supplemental Materials

**Supplementary Methods**

Data generation by cohort site
Details of Bioinformatic analyses
Details of statistical analyses
Supplemental References

**Supplementary Tables**

Table S1. Contributing cohort sites overview
Table S2. Sample processing by cohort site and sample type
Table S3. Bioinformatic parameters by cohort site and sample type
Table S4. Participant characteristics, infants with gut samples
Table S5. Participant characteristics, example analysis
Table S6. Differentially abundant taxa associated with age, delivery mode, and breast milk consumption (Attached Excel file)

**Supplementary Figures**

Figure S1. Per cohort bioinformatic processing workflow.
Figure S2. All Sample Rarefaction Curve.
Figure S3. Rarefaction Curve by body site.
Figure S4. Cohort, sample, and participant retention during bioinformatic processing
Figure S5. Vaginal microbiome composition by trimester among participants in ECHO using the Silva 138.1 database.
Figure S6. Vaginal microbiome composition by trimester among participants in ECHO using the Optivag database.
Figure S7. Top Lactobacillus species in the Optivag data by cohort and trimester.
Figure S8. Predictors of microbial community structure in vaginal samples.
Figure S9. Maternal gut microbiome is associated with trimester and sample type.
Figure S10. Relationship between gut microbiome community composition and predictors of microbiome community structure.
Figure S11. Within sample diversity and taxonomic composition of child gut samples.
Figure S12. Relationship between gut microbiome community composition and predictors of microbiome community structure.
Figure S14. Upper airway microbiome composition across different metrics and predictors.

### Supplemental Methods

#### Sample collection by cohort site

##### DINE

The DINE (Developmental Impact of NICU Exposures) Study is an ongoing, multicenter prospective cohort of preterm born children developed to study the long-term impact of environmental exposures on child health during the period when preterm infants are in the NICU (1–3). DINE is a part of the Environmental Influences on Child Health Outcomes (ECHO) Consortium and combined data from four existing preterm birth cohorts: Prematurity and Respiratory Outcomes Program (PROP), the Trial of Late Surfactant (TOLSURF) study, the Preterm Erythropoietin Neuroprotection Trial (PENUT), and Neonatal Intensive Care Unit Hospital Exposures and Long-Term Health (NICU-HEALTH) (4–7). Participants were eligible if they were study participants in one of the four extant preterm cohorts and were granted permission to be re-contacted to obtain consent into DINE. Participants were recruited between 2010 and 2016 into their parent cohort study if they were a surviving former preterm infant hospitalized in the NICU at a clinical site participating in one of the four existing preterm birth cohorts. The DINE study was approved by the institutional review boards of all participating clinical sites and parents provided informed consent for infant participation.

In a subset of study participants study as described elsewhere, we sampled gut, nasal, and throat microbiota samples longitudinally from NICU stay through one year of age from pre-term and full-term infants (8). Rectal, nasal, and throat material was collected from pre-term infants from 23 to 37 weeks GA at birth (GAB) weekly until hospital discharge and then monthly through 1 year of age, adjusted for prematurity. Rectal, nasal, and throat samples were collected from full-term infants at enrollment and monthly through 1 year. Each rectal sample was collected by inserting a sterile, normal saline moistened, Copan® flocked nylon swab (Copan Diagnostics, Murrieta, CA) beyond the sphincter into the rectum and twirling prior to removal. Nasal and throat samples were similarly collected by inserting and twirling a sterile, moistened swab into the throat or anterior nostril. Each swab was then immediately placed into sterile buffered saline and stored at 4 °C for no more than 4 h. Samples were processed daily, with the extraction of the fecal, nasal, and throat material from the swabs in a sterile environment and immediate transfer − 80o C storage until DNA extraction.

Total genomic DNA was extracted from the nose, throat, and rectal samples using a modification of the Zymo Fecal/Soil Microbe Miniprep Kit (Zymo Research, Irvine, CA) and FastPrep mechanical lysis (MPBio, Solon, OH). 16S ribosomal DNA (rRNA) was amplified with Phusion High-Fidelity polymerase (Thermo Scientific, Waltham, MA) and dual-indexed primers specific to the V3-V4 (319F: 5′ ACTCCTACGGGAGGCAGCAG 3′; 806R: 3′ ACTCCTACGGGAGGCAGCAG 5′) and V1-V3 (8F: 5′ AGAGTTTGATCCTGGCTCAG 3′; 534R: 3′ ATTACCGCGGCTGCTGG 5′) hypervariable regions (9). Amplicons were pooled and paired-end sequenced on an Illumina MiSeq (Illumina, San Diego, CA) in the University of Rochester Genomics Research Center. Each sequencing run included (1) positive controls consisting of a 1:5 mixture of Staphylococcus aureus, Lactococcus lactis, Porphyromonas gingivalis, Streptococcus mutans, and Escherichia coli genomic DNA and (2) negative controls consisting of sterile saline.

Full detail of specimen processing and DNA sequencing can be found in Grier et al, 2018 (8).

#### MARC-35

Sample collection and sequencing have been previously described (10). Using a standardized protocol, site investigators at 17 sites across 14 US states enrolled infants (age <1 year) hospitalized with an attending physician diagnosis of bronchiolitis during three consecutive bronchiolitis seasons from November 1 to April 30, 2011–2014. We excluded infants with previous enrolment; those who were transferred to a participating hospital 48 hours after the original hospitalization; those who were consented >24 hours after hospitalization; or those with known heart-lung disease, immunodeficiency, immunosuppression or gestational age <32 weeks.

Nasopharyngeal aspirates (NPAs) were performed using a standardized protocol. Designated site personnel were trained using a lecture, written instructions, and video. All of the sites used the same collection equipment (Medline Industries, Mundelein, IL) and collected the samples within 24 hours of a child's arrival on the medical ward or ICU. The nasopharyngeal sample was added to transport medium, immediately placed on ice and then stored at −80°C. Frozen samples were shipped in batches on dry ice to Baylor College of Medicine.

Bacterial genomic DNA was extracted using a MO BIO PowerSoil DNA Isolation Kit (Mo Bio Laboratories, Carlsbad, CA, USA). The 16S rDNA V4 region was amplified by PCR and sequenced in the MiSeq platform (Illumina, SanDiego, CA, USA) using a 2×250 bp paired-end protocol that yields paired-end reads which overlap almost completely. The primers used for amplification contained adapters for MiSeq sequencing and single-end barcodes, allowing pooling and direct sequencing of PCR products.

#### MARC-43

Sample collection and sequencing have been previously described (11). Briefly, using a standardized protocol, site investigators from 5 sites across 4 US states enrolled infants from age 28 days to 11.99 months during well-visit. Cohort exclusion criteria included known heart lung disease, immunodeficiency, immunosuppression, chronic gastrointestinal disorder (except for reflux and constipation), or other major medical disorders; previous hospitalization for a breathing problem; and gestational age <32 weeks; no additional inclusion or exclusion criteria were applied for microbiome sample collection.

In a pilot study, fecal samples were self-collected at home using OMNIGene kits and mailed to MGH, where they were stored at −80°C. In the full cohort, fecal samples were collected from diapers brought to the clinic visit. Diapers containing feces were refrigerated or stored in a cooler by hospital staff immediately after collection. The fecal samples were then placed in sterile Sarstedt feces collection containers (Sarstedt, Nümbrecht, Germany). All fecal samples were immediately stored at −80°C upon receipt at the study center. In a pilot study, fecal samples were self-collected at home using OMNIGene kits and mailed to MGH, where they were stored at −80°C. Frozen samples were then shipped on dry ice to Baylor College of Medicine, where we characterized the microbiota via 16S rRNA gene sequencing.

Bacterial genomic DNA was extracted with a Mo BIO PowerMag DNA Isolation Kit (Mo Bio Laboratories, Carlsbad, CA). The 16S rDNA V4 region was amplified by polymerase chain reaction (PCR) and sequenced in the MiSeq platform (Illumina, San Diego, CA) via the 2x250-bp paired-end protocol, yielding pair-end reads that overlap almost completely. The primers used for amplification contain adapters for MiSeq sequencing and single-end barcodes, allowing pooling and direct sequencing of PCR products.

#### ECHO RESONANCE (MINNIE and BAMBAM)

Stool samples were collected by parents using OMR-200 collection tubes (OMNIgene GUT, DNA Genotek, Ottawa, ON, Canada), placed on ice, and delivered to the laboratory in Rhode Island within 24 hours, where they were immediately stored at –80 °C. Samples were not obtained if participants had taken antibiotics in the preceding two weeks. DNA extractions were carried out at Wellesley College (Wellesley, MA). Nucleic acids were isolated from stool using the RNeasy PowerMicrobiome kit (Qiagen), with the DNA degradation steps omitted. In brief, cells were disrupted by bead beating with the PowerLyzer 24 Homogenizer (Qiagen, Germantown, MD) at 2500 rpm for 45 seconds. The resulting lysates were then processed on a QIAcube (Qiagen, Germantown, MD) to complete the extraction procedure. Purified DNA was subsequently sequenced at the Integrated Microbiome Resource (IMR, Dalhousie University, Halifax, NS, Canada).

The V4–V5 region of the 16S rRNA gene was sequenced according to the protocol described by Comeau et al (12). Briefly, the DNA extracts were amplified using the Phusion High-Fidelity DNA polymerase (Thermo Fisher Scientific, Waltham, MA, United

States) and universal bacterial primers 515FB: 5′-GTGYCAGCMGCCGCGGTAA-3′ and 926R: 5′-CCGYCAATTYMTTTRAGTTT-3′ (13, 14). These primers had appropriate Illumina adapters and error-correcting barcodes unique to each sample to allow up to 380 samples to be simultaneously run per single flow cell. Samples were pooled into a single library and quantified fluorometrically, and then they were cleaned up and normalized using the high-throughput Charm Biotech Just-a-Plate 96-well Normalization Kit (Charm Biotech, Cape Girardeau, MO, United States). The normalized samples were sequenced on the Illumina MiSeq platform (Illumina, San Diego, CA, United States) using 300 + 300 bp paired-end V3 chemistry, producing on average 55,000 raw reads per sample (12).

#### ECHO Emory

Participants for this study were enrolled in the Emory University African American Vaginal, Oral, and Gut Microbiome in Pregnancy Study (15). Briefly, pregnant African American patients were recruited from prenatal clinics of two hospitals in Atlanta, GA. Consenting participants completed biological sample collection upon enrollment at 8-14 weeks' gestation. Participants were provided verbal and pictorial instructions explaining how to obtain self-collected vaginal and rectal swabs. The vaginal collection involved sampling the midportion of the vaginal vault (3–4 inches from introitus). The oral collection involved sampling the tongue, hard palate, and gum line. The rectal sample involved sampling the rectal vault 1 inch beyond the anal sphincter. Consistent with the protocols of the Human Microbiome Project (16), the sampling at each body site used a Sterile Catch-All™ Sample Collection Swab (Epicentre Biotechnologies, Madison WI) that was immediately handed to the study coordinator for placement in MoBio bead tubes (MoBio Laboratories, Inc.,) that were frozen upright on ice until transported to the lab, where they were stored at −80 °C until DNA extraction.

Child fecal samples were collected from the child of African American mothers who were enrolled in our pregnancy study (described above), who re-consented with their infant following the birth of the child (15). Mother-infant pairs were subsequently visited at home at 1-week, and 3-, 6-, 12- and 18-months post-birth for data and sample collection.

Enrolled mothers were provided verbal and pictorial instructions explaining how to obtain the child stool samples from a freshly voided infant diaper. All infant stool samples are obtained by the mother using four Catch All swabs, as recommended per protocols of the Human Microbiome Project (16). The mother used each swab to collect a small amount of stool from the freshly voided diaper stool sample and then places the sample back into pre-labeled hard plastic case. The samples were stored in the home freezer until collection by the study team, ideally within 72 hours. Research staff transferred the samples into labeled tubes at the home or transport them to the research laboratory where the specimens were stored in a −80 freezer.

**DNA Extraction and Sequencing**. DNA was extracted from participant swab samples using the DNeasy PowerSoil Kit (cat# 12888-100, Qiagen). DNA quantification based on a threshold of 5 ug/nL was used to identify samples that were borderline in terms of DNA yield; in cases that were borderline, DNA quality was assessed on a 2% agarose gel and quantitated with the Broad Range Quant-It kit from ThermoFisher Scientific (Q33130). Participant samples with DNA visible on the gel were sequenced as were 30 no-template controls (that contained all assay components except for DNA to verify lack of contamination across reagents and samples) and positive controls (that were a mixture of 20 vaginal specimens of known composition). Microbial composition was characterized by DNA sequencing of the 16S rRNA gene. Amplification of the V3–V4 regions of the 16S rRNA gene was performed using a two step-PCR described previously (17). Briefly, the first PCR used the short 16S rRNA gene specific primers 319F (ACACTGACGACATGGTTCTACA[0–7]ACTCCTRCGGGAGGCAGCAG) and 806R (TACGGTAGCAGAGACTTGGTCT[0–7]GGACTACHVGGGTWTCTAAT) where the underlined sequence is the Illumina sequencing primer sequence and [0–7] indicate the presence of an heterogeneous pad sequence to improve sequencing quality, for a total of 20 cycles (9). The second step extends the amplicon with the Illumina required adaptor sequences and the sample specific dual barcode system via 10 cycles with primers H1 (AATGATACGGCGACCACCGAGATCTACACNNNNNNNNACACTGACGACATGGTTCTACA) and H2 (CAAGCAGAAGACGGCATACGAGATNNNNNNNNTACGGTAGCAGAGACTTGGTCT) where NNNNNN indicates a sample specific barcode sequence and the underlined sequence corresponds to the Illumina sequencing primer for priming to the first step amplicon.4 Amplicons were visualized on a 2% agarose gel, quantified, pooled in equimolar concentration, and purified prior to sequencing on an Illumina HiSeq 2500 (San Diego, CA, USA) modified to generate 300 bp paired-end reads.

#### WISC

**Vaginal sample collection during pregnancy**

Vaginal samples from pregnant mothers were excluded if the mother (i) reported the use of antibiotics (except Group B strep prophylaxis) or corticosteroids in the last month prior to delivery; (ii) works or resides on farm without cattle, cows, pigs or goats, but instead with horses, sheep or poultry ie. sheep-only farms; (iii) working or residing in household with pet farm animals or (iv) residing in nonfarm household < 1/8 mile from animal farm.

Vaginal samples were collected during the mother's OBGYN clinical visit for routine GBS testing. The samples were collected by the OBGYN attending physician, PA or RN using a vaginal swab (Copan ESwab) collection kit. After collection the OBGYN physician or staff send the sample at room temperature via the pneumatic tube system to Marshfield Labs where it is frozen at -80ºC upon arrival. This tube system is monitored 24/7. The sample is stored at -80ºC until shipment to a designated facility, which is completed using dry ice.

Vaginal samples were vortexed with the swab remaining in the tube; 500 µL of transport medium was withdrawn from the tube and added to 500 uL of CTAB extraction buffer

**Fecal sample collection from infants**

There were no additional inclusion or exclusion criteria for microbiome sample collection beyond study enrollment. Samples were collected during the 2, 9, 24, 36, 48, 72 and 96 month home visits. The samples were collected by the caregiver (i.e., self-collection) using diaper liner collection kits for infants and collection “hat” kits for older children. Caregivers are instructed to collect one stool sample from a single bowel movement up to 24 hours before the study staff’s home visit. The diaper or stool should be kept refrigerated at all times and never frozen. The caregiver is asked to record the date and time of collection.

No additional processing was performed on samples collected with the potty hat. For samples collected in diapers, the study staff used a sterile tongue depressor to remove stool from the diaper or, if needed, cut out soiled portions of the diaper liner with a sterile scissors and forceps. Urine stained areas are avoided. The study staff tried not to use any stool that comes in contact with their gloves. The samples are carefully capped and placed into a biohazard bag containing 1 absorbent sheet.

Samples were to the clinic via FedEx, or dropped off at the clinic by the caregiver. The samples were kept on ice during transport and stored at -80ºC when it reached the clinic. The sample was stored at -80ºC until shipment to a designated facility, which is completed using dry ice. At the designated facility, two sub-samples (approximately 0.25 g each) per frozen stool (maintained on dry ice during sub-sampling) were obtained under aseptic conditions in a biosafety cabinet using a sterile punch biopsy prior to being pooled and added to 500 µL of cetyltrimethylammonium bromide (CTAB) extraction buffer (5% CTAB in 0.25 M phosphate buffer and 1M NaCl).

*DNA extraction*

DNA from all samples were extracted using a modified CTAB–polyethylene glycol (PEG) protocol as previously described. Briefly, cells were lysed by bead-beating in Lysis Matrix E tubes (MB Biomedicals) at 5.5 m/s for 30 seconds, phases were separated by centrifugation at 16,000 g for 5 min, and the aqueous layer transferred to a new tube. To improve extraction efficiency, a secondary extraction was performed via addition of 500 µL CTAB to the original extraction tube, bead-beating and phase separation, after which the aqueous layer was mixed with that from the previous extraction. To remove excess phenol, the pooled aqueous layers from both extractions were extracted by vortexing in an equal volume of chloroform followed by centrifugation at 16,000 g for 5 min. The resulting aqueous phase was added to 1mL of PEG precipitating solution (30% PEG 6000 in 1.6M NaCl) and stored overnight at 4°C. Precipitated DNA was recovered by centrifugation for 60 min at 3,000g. DNA pellets were washed twice with 300μL 70% ethanol, air-dried for 10 minutes in a biosafety cabinet, and resuspended in 50 μL of molecular grade water. DNA concentrations were quantified using the Qubit dsDNA HS Assay Kit (ThermoFisher Scientific, MA), diluted to 10 ng/μL−1 and stored at −20°C.

##### *16S rRNA V4 amplicon library preparation and sequencing*

The variable region 4 (V4) of the 16S rRNA gene was amplified in triplicate reactions per sample using 515F/806R primers and PCR conditions previously described. Pooled amplicon reactions with ≥5 ng were purified using the SequalPrep Normalization Plate Kit (ThermoFisher Scientific, MA) according to the manufacturer's specifications, quantified using the Qubit dsDNA HS Assay Kit (ThermoFisher Scientific, MA), and pooled at equimolar concentrations. The amplicon library was concentrated using the Agencourt AMPure XP system (Beckman-Coulter), quantified using the KAPA Library Quantification Kit (APA Biosystems), and diluted to 2 nM. Equimolar PhiX was added at 40% final volume to the amplicon library followed by sequencing on the Illumina NextSeq 500 Platform employing a 2x150 bp sequencing run.

#### COAST

Sample collection and sequencing for this cohort have been previously described (18). Samples were collected from the COAST study participants (19), with no additional inclusion or exclusion criteria within the cohort for sampling. Nasal samples were collected at scheduled visits at 2, 4, 6, 9, 12, 18, and 24 months of age. Samples were taken from mostly well children, but some were sick at their scheduled visit. Additional samples were collected when the children had respiratory illnesses (moderate colds or worse) between birth and 36 months. Nasal lavage samples collected by study

coordinators using a kit (bulb/syringe device) developed and assembled in lab. Samples were either stored in saline or M4RT viral transport media. All samples were stored in -80C freezers.
Samples were shipped on dry ice via courier who replenished dry ice as needed. All samples were confirmed frozen upon arrival. Samples had not been freeze/thawed prior to DNA extraction. Samples were prepared and extracted using the FastPrep Bacterial Genomic DNA Purification protocol. The V4 region (515F-806R) was amplified before sequencing on the MiSeq using a 300 cycle kit.

#### URECA

##### *Stool samples*

Samples were collected at 9- and 10- year visits with no additional inclusion or exclusion beyond cohort enrollment. Stool samples were collected at home using a stool sample collection hat. Sample was placed into a collection cup using a scoop attached to the lid of the cup. Samples were brought to the clinic or mailed after being frozen at home for at least 24 hours. Upon receipt at the clinic, samples were frozen at -80C. Sample collection tubes shipped frozen at -80C.
At the lab, two sub-samples (approximately 0.25 g each) per frozen stool (maintained on dry ice during sub-sampling) were obtained under aseptic conditions in a biosafety cabinet using a sterile punch biopsy prior to being pooled and added to 500 µL of cetyltrimethylammonium bromide (CTAB) extraction buffer (5% CTAB in 0.25 M phosphate buffer and 1M NaCl).

##### *Nasal Samples*

Samples were collected at 12 and 36 months with no additional inclusion or exclusion criteria. Some children were sick at the time of collection; they were included in the study. Nasal mucus was collected by study personnel by nasal lavage using a modified bulb syringe. Approximately 2 mL of saline was squirted into the nose, and lavage fluid was obtained with gentle suction. The samples were kept on ice and brought to the site laboratory where the samples were divided into aliquots and cryopreserved (-80C) pending analysis.

##### *Extraction and Sequencing*

DNA from all samples and several extraction blanks were extracted using a modified CTAB–polyethylene glycol (PEG) protocol as previously described. Briefly, cells were lysed by bead-beating in Lysis Matrix E tubes (MB Biomedicals) at 5.5 m/s for 30 seconds, phases were separated by centrifugation at 16,000 g for 5 min, and the aqueous layer transferred to a new tube. To improve extraction efficiency, a secondary extraction was performed via addition of 500 µL CTAB to the original extraction tube, bead-beating and phase separation, after which the aqueous layer was mixed with that from the previous extraction. To remove excess phenol, the pooled aqueous layers from both extractions were extracted by vortexing in an equal volume of chloroform followed by centrifugation at 16,000 g for 5 min. The resulting aqueous phase was added to 1mL of PEG precipitating solution (30% PEG 6000 in 1.6M NaCl) and stored overnight at 4°C. Precipitated DNA was recovered by centrifugation for 60 min at 3,000g. DNA pellets were washed twice with 300µL 70% ethanol, air-dried for 10 minutes in a biosafety cabinet, and resuspended in 50 µL of molecular grade water. DNA concentrations were quantified using the Qubit dsDNA HS Assay Kit (ThermoFisher Scientific, MA), diluted to 10 ng/µL−1 and stored at −20°C.

The variable region 4 (V4) of the 16S rRNA gene was amplified in triplicate reactions per sample using 515F/806R primers and PCR conditions previously described. Pooled amplicon reactions with ≥5 ng were purified using the SequalPrep Normalization Plate Kit (ThermoFisher Scientific, MA) according to the manufacturer's specifications, quantified using the Qubit dsDNA HS Assay Kit (ThermoFisher Scientific, MA), and pooled at equimolar concentrations. The amplicon library was concentrated using the Agencourt AMPure XP system (Beckman-Coulter), quantified using the KAPA Library Quantification Kit (APA Biosystems), and diluted to 2 nM. Equimolar PhiX was added at 40% final volume to the amplicon library followed by sequencing on the Illumina NextSeq 500 Plat-form employing a 2x150 bp sequencing run.

#### INSPIRE

**Study population**

The Infant Susceptibility to Pulmonary Infections and Asthma Following Respiratory Syncytial Virus Exposure study (INSPIRE) is a large, population-based, birth cohort of previously healthy, term, non-low-birth-weight infants born between June and December of 2012 and 2013. In brief, eligible children were enrolled near birth (median [interquartile range] age at enrollment = 55 [16-78] days) at one of 11 participating pediatric practices throughout the middle Tennessee region. The catchment zone encompassed urban, suburban, and rural areas. In children enrolled during the 2nd year of the study, the enrollment visit included the collection of a nasal filter and stool sample for human microbiome studies. Yearly follow-up for the development of common childhood respiratory illnesses is ongoing. One parent of each children provided informed consent for participation. The Institutional Review Board of Vanderbilt University approved this study. The detailed methods for INSPIRE have been previously reported (20, 21).

The inclusion criteria included: 1) singleton birth to a mother age ≥18 years, 2) estimated gestational age ≥37 weeks, 3) birth weight ≥2,250 grams, 4) birth between June 1 and December 31 of 2012 or 2013, 5) parent or guardian able to understand and provide informed consent, and 6) no intent to relocate from middle Tennessee within 5 years of enrollment. The exclusion criteria included: 1) bronchopulmonary dysplasia, 2) cystic fibrosis, 3) primary or acquired immunodeficiency (including maternal infection with human immunodeficiency virus), 4) significant pulmonary, cardiovascular, or neurological disease, 5) need for mechanical ventilation prior to enrollment, and 6) any medical condition that may jeopardize the integrity of the data to be collected.

In addition to the yearly follow-up, we conducted in-person respiratory illness visits during each child's first respiratory syncytial virus season (November to March in our region) in those who met pre-specified criteria for an acute respiratory infection. These respiratory illness visits included a parental questionnaire, physical exam, and the collection of a nasal wash for human microbiome studies.

Children enrolled were born between June 1 and December 31 of 2012 or 2013. The respiratory illness visits were conducted during the 2012-2013 and 2013-2014 RSV seasons.

**Specimen collection**

Nasal filters were collected at enrollment by trained research staff using an epithelial lining fluid using filter paper made of synthetic absorptive matrix (Leukosorb, Pall Life Sciences). For this, we introduced ~⅓ to ½ of a filter paper measuring ~ 15 x 25 mm into one of the nostrils and placed it laterally against the anterior portion of the inferior nasal turbinate. We then briefly compressed the nasal alae together to facilitate absorption of the URT epithelial lining fluid. The filter paper was left in the nostril for a minimum of 30 seconds and up to 2 minutes. Once removed, it was placed into a sterile container. We subsequently collected a 2nd nasal filter from the other nostril using the same method. Nasal washes were collected during respiratory illness visits by gently flushing and then collecting 5 ml of sterile saline solution (0.9%) into one of the infant's nares using a bulb syringe.

Stool samples were collected at enrollment by trained research staff directly from a fresh diaper during an in-person follow-up visit or from frozen stool collected by parents using a home kit and later shipped to our laboratory in dry ice.

Biospecimens were all transferred to a freezer and stored at -80°C until further processing.

**DNA Extraction, Amplification, and Sequencing**

The methods used to characterize the human microbiome from children enrolled in INSPIRE have been previously described in detail (22–25). Laboratory assays were performed at the Das Laboratory at the J. Craig Venter Institute (Maryland, United States).

Bacterial DNA was extracted from all biospecimens using the PowerSoil Kit (MOBIO/Qiagen), In addition, mechanical lysis of bacterial cell walls was performed by shaking the samples on a vortex for a total of 20 minutes.

For the nasal filters and washes, amplicons targeting the V4 region of the bacterial 16S ribosomal RNA (rRNA) were generated by combining 7 µl of template, 12.5 µl MyTaq HS Mix (Bioline), 0.75 µl DMSO (Sigma), 1 µl PCR Certified Water (Teknova), and 2 µl of each 10 µM primer before each round of PCR. During the first round of PCR, the target region was amplified with primers 515F 5'-GTGCCAGCHGCYGCGGT-3' and 806R 5'-GGACTACNNGGGTWTCTAAT-3', with an initial denaturing step at 95°C for 3 min. This was followed by 10 cycles of 95°C for 30 seconds, 50°C for 30 seconds, and 72°C for 1 second, and a final extension at 72°C for 5 minutes. During the second round of PCR, 20 cycles with the same cycling condition as before were performed to add Illumina adaptors, standard Illumina sequence primer region, a 12 base pair barcode, and random nucleotides to increase sequence diversity. Each amplified sample was run on a 1.2% agarose gel to confirm reaction success. Libraries were cleaned and normalized with the SequalPrep Kit (Thermo Fisher). Normalized amplicons were pooled and cleaned with 1X AMPure XP beads (Beckman Coulter). The pool was run on a 1.5% agarose gel and the target size band was extracted and cleaned with the NucleoSpin Gel and PCR cleanup kit (Macherey-Nagel). Two negative controls (a PCR and a extraction negative control) and 2 positive controls (with known taxonomic composition [provided by the NIH/NIAID Biological and Emerging Infections (BEI) Program]) were amplified and sequenced concurrently along with the participant samples to assess data quality and levels of background contamination. The 2 BEI control reagents obtained through BEI Resources included: 1) Genomic DNA from Microbial Mock Community B (Staggered, Low Concentration), v5.2L, for 16S rRNA Gene Sequencing, HM- 783D, and 2) Genomic DNA from Microbial Mock Community B (Even, Low Concentration), v5.1L, for 16S rRNA Gene Sequencing, HM-782D.

For the stool samples, dual-indexed universal primers appended with Illumina-compatible adapters were used to amplify the hypervariable V4 region of the bacterial 16S rRNA gene. The PCR mix for each library contained 12.5 µl of MyTaq Mix (Bioline), 0.75 µl DMSO, 1 µl of forward primer, 1 µl of reverse primer, 4 µl of sample, and PCR Certified water (Teknova) to achieve a final volume of 25.25 µl. DNA was denatured at 95°C for 2 min, and then 30 cycles of 95°C for 20 seconds, 55°C for 15 seconds, and 72°C were performed. Samples were then incubated at 72°C for 10 min and held at 4°C until removal from the thermocycler. Each amplified sample was run on a 1% agarose gel to verify reaction success. Libraries were cleaned and normalized with the SequalPrep Kit (Thermo Fisher). Following normalization to 1-2 ng/µl, 10 µl of each sample was combined to create the sequencing pool. The pool was cleaned with 1X AMPure XP beads (Beckman Coulter). Multiple negative controls (PCR and extraction negative controls) and a positive control (with known taxonomic composition [ZYMOBiomics]) were amplified and sequenced concurrently along with the participant samples to assess data quality and levels of background contamination. Libraries were sequenced on an Illumina MiSeq with 2x300 base pair reads for all biospecimens.

#### ECHO EGDS

Study recruiters contacted parents in a subsample of linked families, whereby there was at least one additional sibling in the adoptive or biological family home, to collect fecal samples from all siblings to measure microbiome diversity in each sibling. All study procedures were approved by the University of Oregon Internal Review Board (IRB) and all caregivers provided informed consent and youth age 7 and older provided assent. To collect the fecal sample, parents were asked to place a provided paper seat cover on the toilet when the child indicates they need to defecate. For children in diapers, a sample was collected from the diaper. The OMNIgene-gut fecal collection kit was used (which contains a nucleic acid preservative) following the kit's instructions (DNA Genotek; OMR-200). Samples were returned via standard mail at ambient temperature to the University of Oregon. Samples were stored at −20°C upon receipt and transferred to −80°C for long-term storage within 4 weeks of receipt, until DNA extraction.

DNA extraction was conducted using the MoBio PowerFecal DNA isolation kit. Each sample was amplified in triplicate using a standard PCR protocol targeting the 16S rRNA variable region V4 (Illumina 515F and 806R). Samples and no-template negative controls were sequenced on the Illumina HiSeq4000 sequencing platform (paired-end 150-bp reads) with a target sequencing depth of 50,000 reads per sample.

#### VDAART

Stool samples were self-collected by participants from mothers during the 3rd trimester of pregnancy and from offspring at age 3 to 6 months, age 1 year, age 3 years, and annually after that to date. Stool was not collected if participants had used antibiotics in the past 7 days and otherwise there were no additional inclusion/exclusion criteria. Subjects were asked to collect a 0.5 teaspoon-sized sample of stool in a potty hat or from diaper for infants in a preservative-free tube 1 to 2 days before a study visit. Subjects store the sample in a home freezer before transport with a freezer pack to the study site. After delivery to the study site, stool samples were immediately stored at -80° C and then shipped on dry ice to the Channing Division of Network Medicine laboratory at Brigham and Women's Hospital. Stool samples are stored there in aliquots with buffer added to the first aliquot to prep the sample for downstream microbiome sequencing.

For microbiome sequencing, DNA extraction and sequencing of the bacterial 16S V4 hyper-variable region was performed using the NEXTflex 16S Amplicon-Seq Kit 2.0 (515F/806R primers, Bioo Scientific Corp, Austin, TX). Sequencing of samples was performed on the Illumina MiSeq platform (San Diego, CA) at Partners Healthcare Personalized Medicine (Boston, MA).

#### Rochester

Women with normal risk pregnancies were recruited at <14 weeks gestation at the University of Rochester Medical Center, Rochester, NY. Eligibility criteria for pregnant participants included: <14 weeks gestation, age 18 or older, singleton pregnancy, no known substance problems or a history of psychotic illness, and ability to communicate in English. Exclusion criteria included women with major endocrine disorders (e.g., polycystic ovary syndrome, pre-pregnancy diabetes), high-risk pregnancies, or significant obstetrics problems. From the 326 women that were initially enrolled, n=18 participants were subsequently excluded (e.g., multiple pregnancies, miscarriage, medical screen failure). Infants born before 37 weeks of gestation were excluded from the postnatal study phases. Study visits were conducted in a private clinic room and consisted of extensive biospecimen and questionnaire data collection, supplemented with health information abstracted from medical records. All study procedures were approved by the University of Rochester Medical Center (URMC) Internal Review Board (IRB) and all subjects (or caregivers) provided informed consent. Study visits occurred in each trimester with cervicovaginal fluid collected in the third trimester. Child visits occurred at birth and 1, 6, and 12 months of age, at which time rectal swabs were obtained. Collection kits were assembled by clinical coordinators at the University of Rochester. All swabs were stored as quickly as possible at -80oC on site.

Total genomic DNA was extracted immediately after thawing on ice using mechanical lysis with a FastPrep-24 instrument(MP Biomedicals, Irvine, CA) and UV-treated ZymoResearch Quick-DNA Fecal/Soil Microbe Miniprep Kit(ZymoResearch, Irvine, CA). DNA was amplified using Phusion High-Fidelity DNA Polymerase(New England BioLabs, Ipswich, MA) and 16S v3-v4 region specific primers (319F: 5′ ACTCCTACGGGAGGCAGCAG 3′; 806R: 3′GGACTACHVGGGTWTCTAAT 5′) with linkers, followed by dual-indexed linker-sequencing primers compatible with Illumina NexteraXT. Amplicons were pooled and paired-end sequenced on an Illumina MiSeq (Illumina, San Diego, CA) at the University of Rochester Genomics Research Center. Each sequencing run included (1) positive controls consisting of a 1:5 mixture of Staphylococcus aureus, Lactococcus lactis, Porphyromonas gingivalis, Streptococcus mutans, and Escherichia coli genomic DNA and (2) negative controls consisting of sterile water. Additionally, the background microbiome was monitored at multiple stages of sample collection and processing. All sterile saline, buffers, reagents, plasticware, and flocked nylon swabs used for sample collection, extraction, and amplification of nucleic acid were UV irradiated to eliminate possible DNA background contamination. Elimination of potential background from the irradiated buffers, reagents, plasticware, and swabs was confirmed by 16S rDNA amplification. After sample collection, multiple aliquots of sterile saline with swabs used for sample collection were carried through our entire protocol as individual samples, including DNA extraction, 16S rDNA amplification, library construction, and sequencing to monitor potential background microbiome.

#### MARCH

The study population was drawn from the Michigan Archive for Research on Child Health (MARCH) cohort, an ongoing population-based pregnancy and birth cohort set in Michigan's lower peninsula (26). The purpose of the MARCH study is to store biological specimens and other health information in pregnancy and early life that can be used to better understand the causes of problems in pregnancy and optimize the health of children.

##### *Pregnancy Fecal Samples*

Our analysis included pregnant women who provided informed consent for providing stool samples during their pregnancy, typically during the third trimester. MARCH participants who have stated they are willing to participate in additional ancillary studies are contacted to ask if they want to participate in a pregnancy fecal sample collection. Fecal collection kits were sent by mail, and participants return the samples by mail. Typical transit time is 3-4 days.

*Infant Fecal Samples*

Pregnant women provided informed consent for providing infant stool samples to the study. During the MARCH 3 month phone interview mothers confirmed their interest in participating in this sample collection. Fecal collection kits were sent by mail. Fecal samples are collected from infants aged from 3 to 9 months old. Fecal collection kits were sent by mail, and participants return the samples by mail. Typical transit time is 3-4 days.

*Fecal sample sequencing*

Once received in the lab, the fecal samples are aliquoted into sterile tubes and stored at -80°C. DNA is extracted following a modified version of the Human Microbiome Project's protocol. DNA extractions are performed using the MoBio (now Qiagen DNeasy) Powersoil DNA Isolation kit (Qiagen MoBio, Carlsbad, CA) per the Human Microbiome Project's protocol with these alterations: after incubating in C3 solution, the samples were centrifuged for 2 minutes and the DNA was eluted from the spin columns with 50 µL of low EDTA TE buffer (IDT, Coralville, IA) heated to 55°C. Barcoded primers are used to amplify the V4 region of the 16S rRNA gene following the mothur wet lab documentation (27). PCR amplification also follows the wet lab protocol outlined in the mothur documentation. The resulting 16S rRNA libraries are sequenced using 250 base pair Illumina MiSeq with V2 chemistry at the Michigan State University (MSU) genomics core.

*Pregnancy Vaginal Samples*

Our analysis included pregnant women who provided informed consent for providing vaginal swab samples during their pregnancy, typically during the third trimester. MARCH participants who have stated they are willing to participate in additional ancillary studies are contacted to ask if they want to participate in a pregnancy vaginal swab sample collection. Vaginal swab collection kits were sent by mail. Participants self-collected the vaginal swab sample, and participants returned the samples by mail. Typical transit time is 3-4 days.

Samples were processed at two different labs, one at Michigan State University, East Lansing, MI and one at the University of Michigan, Ann Arbor, MI. Following are the sample collection and processing methods at each university.

1. Michigan State University (MSU):
Once received in the lab, the vaginal swabs are aliquoted into sterile tubes and stored at -80°C. DNA is extracted following a modified version of the Human Microbiome Project's protocol. DNA extractions are performed using the MoBio (now Qiagen DNeasy) Powersoil DNA Isolation kit (Qiagen MoBio, Carlsbad, CA) per the Human Microbiome Project's protocol with these alterations: after incubating in C3 solution, the samples were centrifuged for 2 minutes and the DNA was eluted from the spin columns with 50 µL of low EDTA TE buffer (IDT, Coralville, IA) heated to 55°C. Barcoded primers are used to amplify the V4 region of the 16S rRNA gene following the mothur wet lab documentation (27). PCR amplification also follows the wet lab protocol outlined in the mothur documentation. The resulting 16S rRNA libraries are sequenced using 250 base pair Illumina MiSeq with V2 chemistry at the MSU genomics core.

2. University of Michigan (U of M):
DNA is extracted at MSU and transferred to the U of M. At U of M, DNA samples are quantified using the Quant-iT PicoGreen dsDNA Assay kit (cat#: P7589). The V4 region of the 16S rRNA gene was amplified from each sample using the Dual indexing sequencing strategy developed by Dr. Patrick D. Schloss's MiSeq SOP (27).. Sequencing was done on the Illumina MiSeq platform, using a MiSeq Reagent Kit V2 500 cycles (Illumina cat# MS102-2003), according to the manufacturers instructions with modifications found in the Schloss SOP. Accuprime High Fidelity Taq (Life Technologies cat # 12346094 ) was used instead of Accuprime Pfx supermix. PCR was performed using the following conditions (Standard or Touch Down) PCR amplification follows the wet lab protocol outlined in the mothur documentation. The resulting 16S

rRNA libraries are sequenced using 250 base pair Illumina MiSeq with V2 chemistry at the U of M microbiome core.

##### Utah Children's Project

Participants in the Utah children's projects were eligible for microbiome collection with no exclusion criteria. Participant caregivers answered a microbiome sample collection survey on behalf of their child regarding travel, antibiotic, antifungal or antiviral use in the past month, use of nasal medication, exposure to tobacco smoke or vaping, and incidence of runny nose or congestion. Information was documented but participants were not excluded based on responses. Samples were extracted using Zymo DNA Miniprep Plus Kit (Zymo Cat#11-297B) according to manufacturer instructions. Samples were amplified using 515F-806R primers and sequenced using a 2x150nt paired end strategy.

#### Supplemental Bioinformatic Methods

##### Silva database preparation

We selected the Silva 138.1 reference database due to its popularity and broad environmental coverage (28). The database was downloaded using the RESCRIPt plugin for QIIME 2 (29). Briefly, the aligned RNA sequences and corresponding taxa with species level annotation were downloaded for the Silva 138.1 release SSURef 99% database. The sequences were reverse transcribed and filtered to remove sequences with more than five degenerate nucleotides (nt) or homo polymers of more than 8nt in length. Archea under 900nt, Bacteria under 1200nt and Eukeryota under 1400nt were discarded. Full-length sequences were dereplicated and taxonomic assignments were made using the lowest common ancestor.

#### Silva Reference taxonomy for OTU clustering

We constructed a phylogeny for the reference database by aligning the full-length representative sequences using MAFTT (q2-alignment, MAFFT v. 7.490) (30). The data were masked to remove any positions with more than 75% gaps. The reference tree was built using FastTree, and we performed midpoint rooting (q2-phylogeny; FastTree v. 2.1.10) (31).

##### *Optivag database preparation*

Vaginal communities require species-level annotation, within the limits available to a primer pair. To achieve this, we used the Optivag database (v.0.3), a manually curated database of vaginal species with a harmonized 16S and metagenomic reference (32). Database sequences and taxonomy were downloaded from GitHub and imported into QIIME 2 (33).

##### *ASV classification and species-level collapse*

One cohort site had both forward and reverse amplicons where the reverse amplicon was loaded on the sequencer from the 3' end (Table S2). We reverse complemented the "reversed" reads using scikit-bio, and re-imported the amplicons into QIIME 2 (33, 34). We treated the forward and reverse complemented reads as separate cohort sites for the purpose of processing. For each unique amplicon within cohorts with vaginal data, we performed taxonomic classification in a two-step procedure. First, we used RESCRIPt to extract reference from the Silva database matching our specific ASV; extracted amplicons less than 100nt were excluded (28, 29). This provides an exact sequence-length match, rather than a primer-specific classifier. The extracted amplicons were dereplicated with RESCRIPt using default parameters. We built a region-specific naive Bayesian classifier for each table using q2-feature-classifer. These were used to filter mitochondrial sequences out of the vaginal data because mitochondria are not included in the Optivag database. We then performed vaginal classification. Again, region-specific amplicons were extracted from the Optivag reference database and dereplicated using the "super" consensus mode for taxonomy (lowest common ancestor followed by preferentially selecting annotated genera) (32). This was used to build a naive Bayesian classifier to classify the region-specific sequences. Taxonomic annotations were propagated forward, so ASVs that could not be classified to species level were given a species name based on the genus (e.g. an ASV classified as g__Lactobacillus without a species assignment would be give a species assignment of g__Lactobacillus as well.) All ASV tables were collapsed to species level and combined to provide a harmonized table. Taxonomic assignments are given by feature (species) names; phylogenetic comparisons are not possible for taxonomic assignments from the Optivag database.

#### Supplemental Statistical Methods

##### Data description

The number of participants in each category were tabulated; percent values were calculated excluding missing values. Count values less than five were masked using custom suppression code. Differences in count data were tested using a chi-square test excluding missing values. Continuous variables were summarized as mean and standard deviations, median and interquartile region, and range as well as the count of missing values. Differences in distributions were tested using the Kruskal-Wallis test.

##### Alpha Rarefaction Curve

We generated rarefaction datasets by randomly selecting 100 samples with at least 1,000 sequence/sample (or the entire cohort if there were fewer than 100 samples) from each cohort for each body site/participant type combination. Samples were rarefied to 100, 200, 300, 400, 500, 600, 700, 800, 900, 1000, 2000, 2500, 3000, 4000, 5000, 6000, 7500, 7500, 8000, 9000, 10000, 15,000 and 20,000 sequences/sample and per-sample depth Shannon diversity was calculated as the average of 15 rarefactions using QIIME 2 (33). Shannon diversity for a data set was calculated as the average of all samples in the data set at a given depth; the standard deviation was the pooled standard deviations (35). The portion of samples retained overall was calculated by determining the number of samples retained at each depth where a sample was lost; samples, lost in the rarefied subset were calculated based on the rarefaction threshold.

#### Taxonomy bar plots

The target clade level for each taxonomy bar plot was determined based on the resolution needed for comparison at the body site. At each body site (Vaginal, Gut, Upper Airway), all samples from a cohort site were averaged by participant type (e.g., a cohort with both maternal gut and child gut samples would contribute two averages.). For all body sites, the top five classes are shown (5.0% average relative abundance or higher); all other taxa were collapsed into a category labeled as "other". Taxonomic levels were colored using the "Set2" colormap from colorbrewer2. Vaginal samples were collapsed to genus level for both the Silva and Optivag table; genera were retained if they had an average relative abundance of at least 1% in either table. Top genera for upper airway samples were selected based on the 7 most abundant genera with an average relative abundance of at least 1%.

Taxonomic categories for gut samples were determined across all cohorts except DIN. The top five most abundant phyla with an average relative abundance of at least 1% across pooled cohorts were identified. Taxa from any other phylum was collapsed into an "Other Bacteria" category. Within each abundant phylum, the four most abundant families with an average pooled relative abundance of at least 1% were also identified. All other families in these phyla were collapsed into a fifth category, designated as other organisms within that phylum. Fecal data were colored based on the Microshades CVD palette; each phylum was assigned a hue and families within the phylum were assigned a shade, with darker shades indicating more abundant families (36). The "other" category in each phylum was always assigned a lighter shade than explicitly defined clades.

#### Beta Diversity Principal Coordinates Plots

Distance matrices calculated at 1000 sequences/sample rarefaction depth were used. Distances were projected in principal coordinates (PCoA) ordination space using the QIIME 2 q2-diveristy plugin (33). Two dimensional PCoAs with marginal plots were created using matplotlib (v. 3.6.0) and seaborn (v. 0.12.2) (37, 38).

#### Primer position along the E. coli 16S sequence

To display cohort-specific amplicons, we downloaded the 16S sequences from *Escherichia coli* from several ATCC isolates (ATCC catalogue numbers 11775, 19138, 25922, 35218, 35421, 700414, 700415, 700416, 700417, 700973; 700928, and BAA_2216) through the ATCC Genome Portal (39). The sequences were imported into QIIME 2 and dereplicated using the RESCRIPt plugin (29).

The overall entropy at a position was determined by generating a multiple sequence alignment for a randomly 5% (n= 21775) of the Silva 138.1 reference database using MAFFT (v. 7.52) implemented in the q2-alignment plugin (28, 30, 33). The ATCC *E. coli* sequences were added to this alignment, and the data was filtered to exclude gap positions in the *E. coli* sequences. Shannon Entropy at each non-gap position was calculated using scikit-bio and divided by the maximum entropy possible for the position (34, 35).

For each cohort, primer pair, and sequencing direction, we created a representative *E. coli* amplicon, designed to mimic what the *E. coli* reference sequence would look like if it had been amplified with the primer pair, sequenced, and processed with our data. One cohort had mixed orientation reads, which were not clearly labeled at submission. In this case, we built a cohort-specific phylogenetic tree using the QIIME 2 alignment MAFFT to FastTree Pipeline based on ASVs with at least 100 counts present in at least 5 samples (30, 31, 33). ASVs were portioned based on the root split in the tree; we verified that ASVs across the main split did not co-occur in any samples. ASVs were split into cluster one or cluster two, representing different sequencing directions.

Representative sequences from each cohort, primer, and direct cluster pair were used as "lures" for the RESCRIPt extract-seq-segments function (29, 40). If this was unsuccessful, we reverse completed the lure sequences. Extracted *E. coli* amplicons were dereplicated to identify unique sequences and each sample was mapped to a set of amplicon fragments. These were combined into a consensus sequence with scikit-bio. Representative *E. coli* amplicons and representative primer sequences were aligned to the full-length *E. coli* sequences using MAFTT with the "add-fragment" mode.

#### Overlaps with other measurements and other omics

Overlap UpSet plots are based on a visualization by Lex et al (41). Plots were built using custom code based on the matplotlib and seaborn plotting libraries (37, 38).

20. Larkin EK, Gebretsadik T, Moore ML, Anderson LJ, Dupont WD, Chappell JD, Minton PA, Peebles RS, Moore PE, Valet RS, Arnold DH, Rosas-Salazar C, Das

SR, Polack FP, Hartert TV. 2015. Objectives, design and enrollment results from the Infant Susceptibility to Pulmonary Infections and Asthma Following RSV Exposure Study (INSPIRE). BMC Pulm Med 15:45.

26. Elliott MR, Kerver JM, Drew A, Watson K, Kornatowski B, Norman GS, Copeland GE, Leissou E, Ridenour T, Kruger-Ndiaye S, Ma T, Ruden D, Barone C, Keating DP, Sokol RJ, Johnson CC, Paneth N. 2025. Obtaining a Probability Sample of a Pregnancy Cohort of Births: A Review of the Problem and a Practical Solution. Am J Epidemiol kwaf110.

32. Hugerth LW, Pereira M, Zha Y, Seifert M, Kaldhusdal V, Boulund F, Krog MC, Bashir Z, Hamsten M, Fransson E, Svarre-Nielsen H, Schuppe-Koistinen I, Engstrand L. 2020. Assessment of In Vitro and In Silico Protocols for Sequence-Based Characterization of the Human Vaginal Microbiome. mSphere 5.

33. Bolyen E, Rideout JR, Dillon MR, Bokulich NA, Abnet CC, Al-Ghalith GA, Alexander H, Alm EJ, Arumugam M, Asnicar F, Bai Y, Bisanz JE, Bittinger K, Brejnrod A, Brislawn CJ, Brown CT, Callahan BJ, Caraballo-Rodríguez AM, Chase J, Cope EK, Da Silva R, Diener C, Dorrestein PC, Douglas GM, Durall DM, Duvallet C, Edwardson CF, Ernst M, Estaki M, Fouquier J, Gauglitz JM, Gibbons SM, Gibson DL, Gonzalez A, Gorlick K, Guo J, Hillmann B, Holmes S, Holste H, Huttenhower C, Huttley GA, Janssen S, Jarmusch AK, Jiang L, Kaehler BD, Kang KB, Keefe CR, Keim P, Kelley ST, Knights D, Koester I, Kosciolek T, Kreps J, Langille MGI, Lee J, Ley R, Liu Y-X, Loftfield E, Lozupone C, Maher M, Marotz C, Martin BD, McDonald D, McIver LJ, Melnik AV, Metcalf JL, Morgan SC, Morton JT, Naimey AT, Navas-Molina JA, Nothias LF, Orchanian SB, Pearson T, Peoples SL, Petras D, Preuss ML, Pruesse E, Rasmussen LB, Rivers A, Robeson MS, Rosenthal P, Segata N, Shaffer M, Shiffer A, Sinha R, Song SJ, Spear JR, Swafford AD, Thompson LR, Torres PJ, Trinh P, Tripathi A, Turnbaugh PJ, Ul-Hasan S, van der Hooft JJJ, Vargas F, Vázquez-Baeza Y, Vogtmann E, von Hippel M, Walters W, Wan Y, Wang M, Warren J, Weber KC, Williamson CHD, Willis AD, Xu ZZ, Zaneveld JR, Zhang Y, Zhu Q, Knight R, Caporaso JG. 2019. Reproducible, interactive, scalable and extensible microbiome data science using QIIME 2. 8. Nature Biotechnology 37:852–857.

### Supplemental tables

#### Table S1. Contributing cohort sites overview

| Cohort Name | Cohort site study population, design, and inclusion | Specimen type | Microbiome Study Design | Sample Collection Period | Sampling age median [IQR]; range | Num. participants (Num. samples) | Data Accessions |
|---|---|---|---|---|---|---|---|
| Developmental Impact of NICU Exposures (DINE) | Participants were enrolled perinatally from a hospital-based Included children were admitted to the NICU (either <1500g or <32 weeks gestation) with no congenital abnormalities | Child gut (rectal swab)<br>Child upper airway (nasal) | | 2011 - 2017<br>2013 - 2017 | 0.3 [0.1, 0.8] 0.0-3.4 years<br>0.5 [0.2, 0.9] 0.0-3.4 years | 88 (1191)<br>57 (702) | dbGaP phs001347.v3.p2 dbGaP phs001297.v1.p1<br>dbGaP phs001347.v3.p2 |
| 35th Multicenter Airway Research Collaboration (MARC-35) | Hospital-based prospective cohort study of infants admitted to the hospital with bronchiolitis. Participants with known heart-lung disease, immunodeficiency, immunosuppression, or gestational age <32 weeks were excluded. | Child upper airway (nasopharyngeal) | Samples collected during index bronchiolitis hospitalization | 2011 - 2014 | 0.3 [0.1, 0.5] 0.0-1.0 years | 815 (815) | ImmPort SDY1182 |
| 43rd Multicenter Airway Research Collaboration (MARC-43) | Clinic-based enrollment of healthy infants at well visits. Children were excluded if they had been diagnosed with certain comorbidities (heart-lung disease, immunodeficiency, immunosuppression, or chronic gastrointestinal disorder); had a fever, respiratory or gastrointestinal illness; or had received antibiotic treatment within the 7 d prior to enrollment | Child gut (feces) | Samples collected from diapers brought to clinical visit or self-collected at home. Time points include infancy, 1yr, 2yr, and 3yr visits. | 2013 - 2019 | 0.8 [0.3, 1.1] 0.0-5.0 years | 696 (1329) | ImmPort SDY1646 Samples collected 2015-2019 |
| Brown university Assessment of Myelination and Behavior Across normal Maturation (BAMBAM) | General population-based prospective cohort. Participants were healthy neurotypically developing children with no personal or family history of learning, intellectual or psychiatric disabilities less than 12 years of age at the time of enrollment. | Child gut (feces) | Samples collected from diapers brought to clinical visit. Participants were excluded if they had taken antibiotics within the two weeks prior to sampling | 2017 - 2022 | 5.3 [2.6, 8.5] 0.2-17.0 years | 306 (506) | SRA PRJNA695570 |

| Cohort Name | Cohort site study population, design, and inclusion | Specimen type | Microbiome Study Design | Sample Collection Period | Sampling age median [IQR]; range | Num. participants (Num. samples) | Data Accessions |
|---|---|---|---|---|---|---|---|
| | Children were excluded if they had any major risk factors of abnormal in-utero or brain development; prenatal exposure to alcohol, tobacco, or drugs; were born at less than 32 weeks gestation; had a birth weight of less than 1500g or were admitted to the NICU. | | | | | | |
| Maternal health Influences and Nutrition in Neonatal and Infant dEvelopment (MINNIE) | Prenatal enrollment Community-based recruitment of healthy singleton pregnancies from neurotypical families. Pregnancies were excluded if there was any history of psychiatric disorder in the mother or any history of intellectual, learning, or psychiatric disability in first degree relatives. Children were excluded if they were born before 32 weeks gestation, admitted to the NICU, or had epilepsy | Pregnancy gut (feces) | Samples self-collected at home and brought to the clinic the next day. Participants were excluded if they had taken antibiotics within the two weeks prior to sampling | 2018 - 2022 | Second and third trimester Maternal age: 30 [27, 34] 19-43 years | 152 (206) | SRA PRJNA695570 |
| | | Child gut (feces) | Samples collected from diapers brought to clinical visit. Participants were excluded if they had taken antibiotics within the two weeks prior to sampling | 2018 - 2022 | 0.5 [0.3, 1.1] 0.1-3.6 years | 132 (195) | SRA PRJNA695570 |
| Atlanta ECHO Cohort of Emory University (ECHO Emory) | General population pregnancy cohort of English speaking African American women with healthy singleton gestations enrolled at 8-14 weeks gestation. Pregnancies were excluded if the participant had chronic medical condition or use of perspective | Pregnancy Vaginal | Self collected sample at clinic visit upon enrollment | 2014 - 2018 | First and second trimester (6 - 17 weeks gestation) Maternal age: 24 [22, 28] 18-40 years | 393 (393) | SRA PRJNA725416 Vaginal microbiome potential and preterm birth |

| Cohort Name | Cohort site study population, design, and inclusion | Specimen type | Microbiome Study Design | Sample Collection Period | Sampling age median [IQR]; range | Num. participants (Num. samples) | Data Accessions |
|---|---|---|---|---|---|---|---|
| | medication at the time of enrollment | Pregnancy gut (rectal) | Self collected sample at clinic visit upon enrollment | 2014 - 2018 | First and second trimester (6 - 17 weeks gestation)<br>Maternal age: 24 [22, 28]<br>18-40 years | 373 (373) | No |
| | | Child gut (feces) | Longitudinal collection of fecal samples at 1 week, 3, 6, 12, and 18 months post birth | 2015 - 2018 | 1.1 [0.3, 2.1]<br>0.0-3.8 years | 144 (286) | No |
| Wisconsin Infant Study Cohort (WISC) | Clinic-based and community-based enrollment of healthy pregnancies and children residing in Wisconsin with and without exposure to farm animals.<br>Exposure to farm animals was defined as residing or working on a farm with cattle, cows, or goats. Pig-only farms or farms with pigs and non-cattle, cow, or goat livestock were enrolled at the PI's discretion<br>Pregnancies with prenatal infection and children born before 34 weeks were excluded. | Pregnancy vaginal | Sample collection: Sample is collected during the mother's OBGYN clinical visit for routine GBS testing.<br>Samples were excluded if the participant used any corticosteroids or antibiotic except group B strep prophylaxis in the month prior to delivery | 2014 - 2020 | 3rd trimester<br>Maternal age: 30 [27, 33]<br>18-41 years | 144 (144) | SRA PRJEB46659 |
| Wisconsin Infant Study Cohort (WISC) | | Child gut (feces) | Longitudinal sample collection during the 2, 9, 24, 36, 48, 72 and 96 month home visits. | 2014 - 2021 | 0.4 [0.2, 0.8]<br>0.1-1.0 years | 249 (451) | SRA PRJEB46659<br>2-month infant stool samples only |

| Cohort Name | Cohort site study population, design, and inclusion | Specimen type | Microbiome Study Design | Sample Collection Period | Sampling age median [IQR]; range | Num. participants (Num. samples) | Data Accessions |
|---|---|---|---|---|---|---|---|
| Childhood Origins of Asthma Study (COAST) | Clinic-based recruitment of healthy pregnancies from families with a history of asthma or allergies. Children were excluded if they were preterm (less than 37 weeks completed gestation) | Child upper airway (nasal) | Nasal samples were collected at scheduled visits at 2, 4, 6, 9, 12, 18, and 24 months of age. Samples were taken from mostly well children. Additional samples were collected when the children had respiratory illnesses between birth and 36 months. | 1998 - 2003 | 1.0 [0.5, 1.8] 0.0-3.6 years | 277 (2927) | No |
| Urban Environment and Childhood Asthma (URECA) | A clinic and community-based prospective cohort study of children in urban areas with a familiar risk for asthma. English or Spanish speaking participants were enrolled prenatally if they planning to delivery at a hospital, a family history of asthma, allergic rhinitis, eczema, and reside in a census tract with at least 20% of the population below the federal poverty line with plans to remain in the area. Pregnancies were excluded if the mother was living with HIV. Children were included if they were born after 34 weeks or more completed gestation and provided a suitable cord blood sample. | Child gut (feces) | Samples were collected cross sectionally at 9- and 10- year visits | 2014 - 2017 | 9.1 [9.0, 9.2] 8.9-10.5 years | 366 (366) | No |
| Urban Environment and Childhood Asthma (URECA) | Children were excluded if they experienced respiratory distress requiring ventilation for more than 4 hours or requiring supplemental oxygen or CPAP for more than 4 days; pneumonia | Child upper airway (nasal) | Samples were collected at 12 and 36 months. No additional inclusion or exclusion criteria | 2006 - 2010 | 2.8 [2.0, 3.1] 0.9-3.5 years | 356 (598) | No |

| Cohort Name | Cohort site study population, design, and inclusion | Specimen type | Microbiome Study Design | Sample Collection Period | Sampling age median [IQR]; range | Num. participants (Num. samples) | Data Accessions |
|---|---|---|---|---|---|---|---|
| | requiring antibiotic use for one week or more; had significant congenital abnormalities; or were administrated palivizumab (Synagis) for RSV prophylaxis. | | | | | | |
| Infant Susceptibility to Pulmonary Infections and Asthma Following RSV Exposure (INSPIRE) | Community-based enrollment of healthy infants<br>Eligible children were recruited and enrolled near birth from 11 participating pediatric practices across middle Tennessee. Inclusion required: singleton birth to mother >= 18 years; infant being overall healthy, term, and born between June and December of 2012 and 2013; parent or guardian able to understand and provide informed consent; and no intent to relocate from middle Tennessee within 5 years of enrollment | Child gut (feces) | Stool samples were collected during health at the time of enrollment. | 2013 - 2014 | 0.1 [0.0, 0.2]<br>0.0-0.5 years | 113 (113) | No |
| | | Child upper airway (nasal) | Nasal samples were collected from healthy children at the time of enrollment<br>In addition, nasal samples were collected during an acute respiratory illness using a nasal wash | 2012 - 2014 | 0.3 [0.2, 0.5]<br>0.0-0.8 years | 916 (1468) | No |
| Early Growth and Development Study Pediatric Cohort | Domestic adoptions of term children without congenital abnormalities adopted within 3 months of birth placed with a non-biologically related family, and the biological and unrelated siblings of the adopted child (reared either by biological parent or by adoptive parents) | Child gut (feces) | Children, their biological, and adoptive siblings were sampled cross sectionally | 2016 - 2017 | 9.9 [8.6, 12.0]<br>6.0-13.6 years | 33 (33) | No |
| Vitamin D Antenatal Asthma Reduction Trial (VDAART) | Clinic based randomized control trial of vitamin D exposure in pregnancy. Participants were enrolled prenatally | Pregnancy gut (feces) | Sample collected in third trimester. Participants were excluded if antibiotics were taken within 7 days of sampling | 2011 - 2011 | third trimester (29-40 weeks gestation)<br>Maternal age: 30 [23, 32]<br>18-40 years | 101 (119) | SRA PRJNA1074705 |

| Cohort Name | Cohort site study population, design, and inclusion | Specimen type | Microbiome Study Design | Sample Collection Period | Sampling age median [IQR]; range | Num. participants (Num. samples) | Data Accessions |
|---|---|---|---|---|---|---|---|
| | | Child gut (feces) | longitudinal sampling at 3 months, 6 months, 1 year, 3 years, and annual after 3 years of age | 2011 - 2016 | 3.0 [1.0, 4.0] 0.2-5.3 | 566 (1484) | |
| Rochester | Clinic-based enrollment of women with healthy singleton pregnancies Eligibility criteria for pregnant participants included: <14 weeks gestation, age 18 or older, singleton pregnancy, no known substance problems or a history of psychotic illness, and ability to communicate in English. Exclusion criteria included women with major endocrine disorders (e.g., polycystic ovary syndrome, pre-pregnancy diabetes), high-risk pregnancies, or significant obstetrics problems | Pregnancy vaginal | Cervicovaginal fluid collected in the third trimester | 2016 - 2019 | Gestational age: third trimester (34-40 weeks) Maternal age: 30 [26, 33] 19-41 years | 168 (168) | SRA PRJNA1099167 |
| | | Child gut (rectal swab) | Rectal swabs were collected birth and 1, 6, and 12 months of age | 2016-2021 | 0.5 [0.1, 1.0] 0.0-1.5 years | 238 (633) | SRA PRJNA1099167 |
| Michigan Archive for Research in Child Health (MARCH) | Clinic-based enrollment of women with healthy singleton pregnancies<br><br>English-speaking participants who were at least 18 years of age at their first prenatal visit in the sampled clinics | Pregnancy vaginal | Vaginal samples were collected at home using a mailed kit Mother opted-in to collection during pregnancy via separate consent (MSU IRB STUDY00000383) | 2018 - 2019 | First, second, third trimester (8-38 weeks gestation) Maternal age: 31 [29, 34]; 20-43 years | 68 (68) | SRA PRJNA1218274 A subset of mother-child dyads |
| MARCH | | Pregnancy gut (feces) | Samples were collected using a kit by mail. Sampling was primary performed in the third trimester. | 2018 - 2020 | second, third trimester (25-41 weeks gestation) Maternla age: 31 [28, 34] 20-52 years | 81 (81) | SRA PRJNA1218274 A subset of mother-child dyads |

| Cohort Name | Cohort site study population, design, and inclusion | Specimen type | Microbiome Study Design | Sample Collection Period | Sampling age median [IQR]; range | Num. participants (Num. samples) | Data Accessions |
|---|---|---|---|---|---|---|---|
| | | | Mother opted-in to collection during pregnancy via separate consent (MSU IRB STUDY00005020, LEGACY17-1352) | | | | |
| | | Child gut (feces) | Fecal collection kits were sent by mail. Fecal samples are collected from infants aged from 3 to 9 months old. In some cases, children were sampled longitudinally. Parents opted in to collection during 3 month visit | 2018 - 2022 | 0.3 [0.3, 0.5] 0.2-4.3 years | 350 (428) | SRA PRJNA1218274 A subset of mother-child dyads |
| Utah Children's Project (National Children's study Salt Lake; Nation Children's Study Cache County; HOPE) | Population-based sampling of women who are pregnant, trying to conceive, or those who may become pregnant; community-based enrollment of women who are pregnant or trying to connive; clinic-based recruitment of women with singleton pregnancies | Child upper airway (nasal) | There were no additional inclusion or exclusion criteria | 2017 - 2019 | 6.5 [4.5, 8.0] 0.2-18.7 years | 435 (435) | No |

**Table S2. Sample processing by cohort site and sample type**

| Cohort Site | Sample Type | Collection protocol | DNA extraction | 16S primer pair | sequencing strategy | Num. sequencing batches | Num. samples |
|---|---|---|---|---|---|---|---|
| DINE | Child gut (rectal swab) | Collected on site by trained staff without preservative | Qiagen Fecal kit; single tube extraction | V3-V4 (319F-805R) | Illumina MiSeq 2x300 loaded from 3' | 5 | 289 |
| DINE | Child gut (rectal swab) | Collected on site by trained staff without preservative | Zymo Fec So Quick DNA; single tube extraction | V3-V4 (319F-805R) | Illumina MiSeq 2x300 loaded from 3' | 23 | 902 |
| DINE | child upper airway (nasal) | Collected on site by trained staff without preservative | Zymo Fec So Quick DNA; single tube extraction | V1-V3 (27F-539R) | Illumina MiSeq 2x300 loaded from 3' | 18 | 702 |
| MARC-35 | Child upper airway (nasopharyngeal) | Samples were collected on site by trained staff using a storage kit with preservative | MoBio PowerSoil Kit | V4 (515F-806R) | Illumina MiSeq 2x250 | 1 | 815 |
| MARC-43 | Child gut (fecal sample) | Samples were self-collected at home using a collection kit with preservative | MoBio PowerMag Kit | V4 (515F-806R) | Illumina MiSeq 2x250 | 1 | 73 |
| MARC-43 | Child gut (fecal sample) | Samples were collected on site by trained staff using a kit without preservative | MoBio PowerMag Kit | V4 (515F-806R) | Illumina MiSeq 2x250 | 4 | 1256 |
| BAMBAM | Child gut (fecal sample) | Samples were collected at home using a kit with preservative and transported on dry ice | RNeasy PowerMicrobiome QIAcube | V4-V5 (515F-926R) | Illumina NextSeq 2x300 | 9 | 506 |
| MINNIE | Pregnancy gut (fecal sample) | Samples were collected at home using a kit with preservative and transported on dry ice | RNeasy PowerMicrobiome QIA cube | V4-V5 (515F-926R) | Illumina NextSeq 2x300 | 6 | 206 |
| MINNIE | Child gut (fecal sample) | Samples were collected at home using a kit with preservative and transported on dry ice. | RNeasy PowerMicrobiome QIAcube | V4-V5 (515F-926R) | Illumina NextSeq 2x300 | 7 | 195 |

| Cohort Site | Sample Type | Collection protocol | DNA extraction | 16S primer pair | sequencing strategy | Num. sequencing batches | Num. samples |
|---|---|---|---|---|---|---|---|
| ECHO Emory | Pregnancy gut (rectal swab) | Samples were self collected on site using a kit with preservative | MoBio PowerSoil Kit | V3-V4 (319F-805R) | Illumina HiSeq 2x300 | 32 | 373 |
| ECHO Emory | Pregnancy vaginal | Samples were self collected on site using a kit with preservative | MoBio PowerSoil Kit | V3-V4 (319F-805R) | Illumina HiSeq 2x300 | 32 | 393 |
| ECHO Emory | Child gut (fecal sample) | Samples were collected on site by the family using a kit with preservative | MoBio PowerSoil Kit | V3-V4 (319F-805R) | Illumina HiSeq 2x300 | 32 | 286 |
| WISC | Pregnancy vaginal | Samples were collected on site by trained staff using a collection kit for culturing | In house chemical based extraction; 96-well plate format | V4 (515F-806R) | Illumina NextSeq 2x150 | 2 | 144 |
| WISC | Child gut (fecal sample) | Samples were collected on site by trained staff using a kit without preservative | In house chemical based extraction; 96-well plate format | V4 (515F-806R) | Illumina NextSeq 2x150 | 3 | 451 |
| COAST | Child upper airway (nasal) | Samples were collected on site by the family using a kit without preservative | FastPrep Bacterial Genomics DNA kit; 96-well plate format | V4 (515F-806R) | Illumina NextSeq 2x150 | 35 | 2927 |
| URECA | Child gut (fecal sample) | Samples were collected on site by the family using a kit without preservative. | In house chemical based extraction; 96-well plate format | V4 (515F-806R) | Illumina NextSeq 2x150 | 1 | 366 |
| URECA | Child upper airway (nasal) | Samples were collected on site by the family using a collection kit intendent for culturing | In house chemical based extraction; 96-well plate format | V4 (515F-806R) | Illumina NextSeq 2x150 | 2 | 598 |
| INSPIRE | Child gut (fecal sample) | Samples were collected by the family without preservative | MoBio PowerSoil Kit; 96-well plate format | V4 (515F-806R) | Illumina MiSeq 2x300 | 1 | 113 |

| Cohort Site | Sample Type | Collection protocol | DNA extraction | 16S primer pair | sequencing strategy | Num. sequencing batches | Num. samples |
|---|---|---|---|---|---|---|---|
| INSPIRE | Child upper airway (nasal) | Samples were collected on site by trained staff | MoBio PowerSoil Kit; 96-well plate format | V4 (515F-806R) | Illumina MiSeq 2x300 | 1 | 1468 |
| EDGS | Child gut (fecal sample) | Samples were collected at home using a kit with preservative. Samples were sent by mail. | Qiagen QIAmp kit; single tube extraction | V4-V5 (515F-926R) | Illumina MiSeq 2x300 | 1 | 33 |
| VDAART | Pregnancy Gut (fecal sample) | Samples were self-collected on site without preservative | Qiagen QIAmp kit | V4 (Schloss V4) | Illumina MiSeq 2x250 | 7 | 119 |
| VDAART | Child Gut (fecal sample) | Samples were collected on site by the family without preservative | Qiagen QIAmp kit | V4 (Schloss V4) | Illumina MiSeq 2x250 | 7 | 1264 |
| VDAART | Child Gut (fecal sample) | Samples were collected on site by the family without preservative | Qiagen QIAmp kit | V4 (Schloss V4) | Illumina MiSeq 2x250 | 8 | 220 |
| Rochester | Pregnancy vaginal | Samples were self collected on site without preservative | Zymo Fec So Quick DNA; 96-well plate format | V3-V4 (319F-805R) | Illumina MiSeq 2x300 loaded from 3' | 1 | 5 |
| Rochester | Pregnancy vaginal | Samples were self collected on site without preservative | Zymo Fec So Quick DNA; 96-well plate format | V3-V4 (319F-805R) | Illumina MiSeq 2x300 | 1 | 163 |
| Rochester | Child gut (rectal swab) | Samples were collected on site by trained staff without preservative | Zymo Fec So Quick DNA; 96-well plate format | V3-V4 (319F-805R) | Illumina MiSeq 2x300 loaded from 3' | 2 | 327 |
| Rochester | Child gut (rectal swab) | Samples were collected on site by trained staff and stored without preservative prior to freezing. | Zymo Fec So Quick DNA; 96-well plate format | V3-V4 (319F-805R) | Illumina MiSeq 2x300 | 2 | 306 |

| Cohort Site | Sample Type | Collection protocol | DNA extraction | 16S primer pair | sequencing strategy | Num. sequencing batches | Num. samples |
|---|---|---|---|---|---|---|---|
| MARCH | Pregnancy Gut (fecal sample) | Samples were collected at home using a kit with preservative and sent by mail | MoBio PowerSoil Kit; single tube extraction | V4 (Schloss V4) | Illumina MiSeq 2x250 | 1 | 81 |
| MARCH | Pregnancy Vaginal | Samples were collected at home using a kit with preservative and sent by mail | MoBio PowerSoil Kit; single tube extraction | V4 (Schloss V4) | Illumina MiSeq 2x250 | 1 | 29 |
| MARCH | Pregnancy Vaginal | Samples were collected at home using a kit with preservative and sent by mail | MoBio PowerSoil Kit; single tube extraction | V4 (Schloss V4) | Illumina MiSeq 2x250 | 1 | 39 |
| MARCH | Child Gut (fecal sample) | Samples were collected at home using a kit with preservative and sent by mail | MoBio PowerSoil Kit; single tube extraction | V4 (Schloss V4) | Illumina MiSeq 2x250 | 6 | 428 |
| Utah Children's Project | Child Upper airway (nasal) | Samples were collected on site by the family under observation by trained staff and stored in DNA/RNA preservative. | Zymo BIOMICs DNA kit | V4 (515F-806R) | Illumina NextSeq 2x150 | 4 | 435 |

Table S3. Bioinformatic parameters by cohort site and sample type

| Cohort Site | Participant Type and body site | 16S primer pair and sequencing length | Discard read without primers | Read joining | Forward trimming | ASV trim length | Clustering strand | Silva 138.1 97% OTU table | EMP 515F 150nt ASV table | Optivag Species Table |
|---|---|---|---|---|---|---|---|---|---|---|
| DINE | Child gut (rectal swab) | V3-V4 (319F-805R) 2x300 loaded from 3' | No | Single[1] | 0 | 250 | Both | Yes | No | No |
| DINE | Child upper airway (nasal) | V1-V3 (27F-539R) 2x300 loaded from 3' | No | Single[1] | 0 | 275 | Both | Yes | No | No |
| MARC-35 | Child upper airway (nasopharyngeal) | V4 (515F-806R) 2x250 | No | Single[2] | 0 | 150 | Plus | Yes | Yes | No |
| MARC-43 | Child gut (fecal sample) | V4 (515F-806R) 2x250 | No | Single[2] | 0 | 150 | Plus | Yes | Yes | No |
| BAMBAM | Child gut (fecal sample) | V4-V5 (515F-926R) 2x300 | No | Single[2] | 0 | 150 | Plus | Yes | Yes | No |
| MINNIE | Pregnancy gut (fecal sample) | V4-V5 (515F-926R) 2x300 | Yes | Single[2] | 0 | 150 | Plus | Yes | Yes | No |

[1]High quality portion of reverse read was too short for read joining
[2]Started with 515F primer; we used only forward reads for all 515F data sets

| Cohort Site | Participant Type and body site | 16S primer pair and sequencing length | Discard read without primers | Read joining | Forward trimming | ASV trim length | Clustering strand | Silva 138.1 97% OTU table | EMP 515F 150nt ASV table | Optivag Species Table |
|---|---|---|---|---|---|---|---|---|---|---|
| MINNIE | Child gut (fecal sample) | V4-V5 (515F-926R) 2x300 | Yes | Single[2] | 0 | 150 | Plus | Yes | Yes | No |
| ECHO Emory | Pregnancy gut (rectal swab) | V3-V4 (319F-805R) 2x300 | No | Paired | 0 | 425 | Plus | Yes | No | No |
| ECHO Emory | Pregnancy vaginal | V3-V4 (319F-805R) 2x300 | No | Paired | 0 | 425 | Plus | Yes | No | Yes |
| ECHO Emory | Child gut (fecal sample) | V3-V4 (319F-805R) 2x300 | No | Paired | 0 | 425 | Plus | Yes | No | No |
| WISC | Pregnancy vaginal | V4 (515F-806R) 2x150 | No | Single[2] | 0 | 150 | Plus | Yes | Yes | Yes |
| WISC | Child gut (fecal sample) | V4 (515F-806R) 2x150 | No | Single[2] | 0 | 150 | Plus | Yes | Yes | No |
| COAST | Child upper airway (nasal) | V4 (515F-806R) 2x150 | No | Single[2] | 0 | 150 | Plus | Yes | Yes | No |
| URECA | Child gut (fecal sample) | V4 (515F-806R) 2x150 | No | Single[2] | 0 | 150 | Plus | Yes | Yes | No |

[1]High quality portion of reverse read was too short for read joining
[2]Started with 515F primer; we used only forward reads for all 515F data sets

| Cohort Site | Participant Type and body site | 16S primer pair and sequencing length | Discard read without primers | Read joining | Forward trimming | ASV trim length | Clustering strand | Silva 138.1 97% OTU table | EMP 515F 150nt ASV table | Optivag Species Table |
|---|---|---|---|---|---|---|---|---|---|---|
| INSPIRE | Child gut (fecal sample) | V4 (515F-806R) 2x300 | No | Single[2] | 0 | 150 | Plus | Yes | Yes | No |
| INSPIRE | Child upper airway (nasal) | V4 (515F-806R) 2x300 | No | Single[2] | 0 | 150 | Plus | Yes | Yes | No |
| ECHO EGDS | Child gut (fecal sample) | V4-V5 (515F-926R) 2x300 | No | Single[2] | 0 | 150 | Plus | Yes | Yes | No |
| VDAART | Pregnancy gut (fecal sample) | V4 (Schloss V4) 2x250 | No | Single[1] | 15 | 225 | Plus | Yes | No | No |
| VDAART | Child gut (fecal sample) | V4 (Schloss V4) 2x250 | No | Single[1] | 15 | 225 | Plus | Yes | No | No |
| VDAART | Child gut (fecal sample) | V4 (Schloss V4) 2x250 | No | Single[1] | 15 | 225 | Plus | Yes | No | No |
| Rochester | Pregnancy vaginal | V3-V4 (319F-805R) 2x300 loaded from 3' | No | Single[1] | 0 | 225 | Both | Yes | No | Yes |
| Rochester | Pregnancy vaginal | V3-V4 (319F-805R) 2x300 | No | Single[1] | 0 | 225 | Both | Yes | No | Yes |

[1]High quality portion of reverse read was too short for read joining
[2]Started with 515F primer; we used only forward reads for all 515F data sets

| Cohort Site | Participant Type and body site | 16S primer pair and sequencing length | Discard read without primers | Read joining | Forward trimming | ASV trim length | Clustering strand | Silva 138.1 97% OTU table | EMP 515F 150nt ASV table | Optivag Species Table |
|---|---|---|---|---|---|---|---|---|---|---|
| Rochester | Child gut (rectal swab) | V3-V4 (319F-805R) 2x300 loaded from 3' | No | Single[1] | 0 | 225 | Both | Yes | No | No |
| Rochester | Child gut (rectal swab) | V3-V4 (319F-805R) 2x300 | No | Single[1] | 0 | 225 | Both | Yes | No | No |
| MARCH | Pregnancy gut (fecal sample) | V4 (Schloss V4) 2x250 | No | Paired | 15 | 225 | Plus | Yes | No | No |
| MARCH | Pregnancy vaginal | V4 (Schloss V4) 2x250 | No | Paired | 15 | 225 | Plus | Yes | No | Yes |
| MARCH | Pregnancy vaginal | V4 (Schloss V4) 2x250 | No | Paired | 15 | 225 | Plus | Yes | No | Yes |
| MARCH | Child gut (fecal sample) | V4 (Schloss V4) 2x250 | No | Paired | 15 | 225 | Plus | Yes | No | No |
| Utah | Child upper airway (nasal) | V4 (515F-806R) 2x150 | No | Single[2] | 0 | 150 | Plus | Yes | Yes | No |

[1]High quality portion of reverse read was too short for read joining
[2]Started with 515F primer; we used only forward reads for all 515F data sets

**Table S4. Participant characteristics, infants with gut samples**

| | All | | Fecal Samples | | Rectal Swabs | |
|---|---|---|---|---|---|---|
| | participant (n=2151) | sample (n=3952) | participant (n=1828) | sample (n=2405) | participant (n=323) | sample (n=1547) |
| Collection Characteristics | | | | | | |
| *Number of samples collected* | | | | | | |
| median [IQR] | 1 [1, 2] | | 1 [1, 2] | | 3 [2, 4] | |
| range | 1 -22 | | 1 - 4 | | 1 -22 | |
| *Number of samples collected* | | | | | | |
| 1 | 1349 (62.7%) | | 1282 (70.1%) | | 67 (20.7%) | |
| 2 | 599 (27.8%) | | 517 (28.3%) | | 82 (25.4%) | |
| 3 or more | 203 (9.4%) | | 29 (1.6%) | | 174 (53.9%) | |
| *Collection year[a]* | | | | | | |
| median [IQR] | 2017 [2014, 2018] | 2017 [2014, 2018] | 2017 [2014, 2018] | 2017 [2015, 2018] | 2018 [2016, 2018] | 2015 [2014, 2018] |
| min, max | 2011 - 2022 | 2011 - 2022 | 2011 -2022 | 2011 -2022 | 2011 - 2021 | 2011 - 2021 |
| *Child age at collection[a], years* | | | | | | |
| mean (std) | 0.32 (0.24) | 0.41 (0.29) | 0.35 (0.22) | 0.45 (0.27) | 0.10 (0.18) | 0.34 (0.30) |
| median [IQR] | 0.28 [0.16, 0.43] | 0.34 [0.17, 0.64] | 0.31 [0.18, 0.46] | 0.36 [0.23, 0.75] | 0.03 [0.00, 0.09] | 0.22 [0.08, 0.55] |
| min, max | 0.00, 1.00 | 0.00, 1.00 | 0.01, 1.00 | 0.01, 1.00 | 0.00, 0.86 | 0.00, 1.00 |
| Child Demographics | | | | | | |
| *Child sex* | | | | | | |
| Male | 1126 (52.3%) | 2029 (51.3%) | 963 (52.7%) | 1273 (52.9%) | 163 (50.5%) | 756 (48.9%) |
| Female | 1025 (47.7%) | 1923 (48.7%) | 865 (47.3%) | 1132 (47.1%) | 160 (49.5%) | 791 (51.1%) |
| *Child ethnicity* | | | | | | |
| non-Hispanic | 1847 (86.0%) | 3477 (88.1%) | 1559 (85.4%) | 2093 (87.2%) | 288 (89.2%) | 1384 (89.5%) |
| Hispanic | <305 (<14.2%) | <475 (<12.0%) | <270 (<14.8%) | <310 (<12.9%) | 35 (10.8%) | 163 (10.5%) |
| missing | <5 | <5 | <5 | <5 | 0 | 0 |

[a]Value for participant reflects the first sample collected
[b]"Other Races" includes participants who did not identify as White, Black, Asian, or Multiple Races as well as participants who identified as American Indian or Alaskan Native and participants who identified and Native Hawaiian or other Pacific Islander.
[c]This measure excludes non-singleton pregnancies; children of multiple gestation are listed as "missing"
[d]Lifetime duration including participants who were sampled before the age of completion.
[e]Asthma diagnosis was made after age 4; children followed fewer than 4 years cannot have asthma diagnoses
[f]Obesity was only determined for children with BMI measurements at age two years or older; children followed fewer than 2 years cannot have obesity diagnoses

| | All | | Fecal Samples | | Rectal Swabs | |
|---|---|---|---|---|---|---|
| | participant (n=2151) | sample (n=3952) | participant (n=1828) | sample (n=2405) | participant (n=323) | sample (n=1547) |
| | Child Demographics (ctd) | | | | | |
| *Child Race* | | | | | | |
| White | 1337 (62.5%) | 2560 (65.3%) | 1124 (61.8%) | 1484 (62.0%) | 213 (66.4%) | 1076 (70.4%) |
| Black | 509 (23.8%) | 861 (21.9%) | 450 (24.8%) | 606 (25.3%) | 59 (18.4%) | 255 (16.7%) |
| Asian | 66 (3.1%) | 85 (2.2%) | <65 (<3.6%) | 78 (3.3%) | <5 (<1.6%) | 7 (0.5%) |
| Multiple Races | 197 (9.2%) | 354 (9.0%) | 160 (8.8%) | 201 (8.4%) | 37 (11.5%) | 153 (10.0%) |
| Other Race[b] | 30 (1.4%) | 63 (1.6%) | 22 (1.2%) | <30 (<1.3%) | 8 (2.5%) | <40 (<2.6%) |
| inconsistent | <5 | 20 | <5 | <5 | <5 | <20 |
| missing | <10 | 9 | <10 | 9 | 0 | 0 |
| *Maternal ethnicity* | | | | | | |
| non-Hispanic | 1706 (88.9%) | 3318 (91.1%) | 1407 (88.2%) | 1884 (89.9%) | 299 (92.6%) | 1434 (92.7%) |
| Hispanic | 212 (11.1%) | 325 (8.9%) | 188 (11.8%) | 212 (10.1%) | 24 (7.4%) | 113 (7.3%) |
| missing | 233 | 309 | 233 | 309 | 0 | 0 |
| *Maternal Race* | | | | | | |
| White | 1216 (64.7%) | 2430 (67.8%) | 992 (63.2%) | 1289 (62.5%) | 224 (72.3%) | 1141 (74.9%) |
| Black | 499 (26.5%) | 873 (24.4%) | 438 (27.9%) | 600 (29.1%) | 61 (19.7%) | 273 (17.9%) |
| Asian | 81 (4.3%) | 120 (3.3%) | 68 (4.3%) | 83 (4.0%) | 13 (4.2%) | 37 (2.4%) |
| Multiple Races | 51 (2.7%) | 89 (2.5%) | 42 (2.7%) | 53 (2.6%) | 9 (2.9%) | 36 (2.4%) |
| Other Race[b] | 33 (1.8%) | 73 (2.0%) | <35 (<2.2%) | 37 (1.8%) | <5 (<1.6%) | 36 (2.4%) |
| inconsistent | 5 | 8 | <5 | <10 | <5 | <5 |
| missing | 266 | 359 | 254 | <340 | 12 | <25 |
| *Highest maternal education* | | | | | | |
| No Bachelor's | 783 (44.2%) | 1406 (40.4%) | <685 (<47.1%) | <905 (<46.7%) | <105 (<32.8%) | <505 (<32.7%) |
| Bachelor's or higher | 990 (55.8%) | 2073 (59.6%) | 772 (53.1%) | 1033 (53.4%) | 218 (68.1%) | 1040 (67.4%) |
| missing | 378 | 473 | <380 | <470 | <5 | <5 |

[a]Value for participant reflects the first sample collected
[b]"Other Races" includes participants who did not identify as White, Black, Asian, or Multiple Races as well as participants who identified as American Indian or Alaskan Native and participants who identified and Native Hawaiian or other Pacific Islander.
[c]This measure excludes non-singleton pregnancies; children of multiple gestation are listed as "missing"
[d]Asthma diagnosis was made after age 4; children followed fewer than 4 years cannot have asthma diagnoses
[e]Obesity was only determined for children with BMI measurements at age two years or older; children followed fewer than 2 years cannot have obesity diagnoses

| | All | | Fecal Samples | | Rectal Swabs | |
|---|---|---|---|---|---|---|
| | participant (n=2151) | sample (n=3952) | participant (n=1828) | sample (n=2405) | participant (n=323) | sample (n=1547) |
| | | | Birth characteristics | | | |
| *Singleton Pregnancy* | | | | | | |
| No | 66 (3.1%) | 374 (9.5%) | <45 (<2.5%) | 59 (2.5%) | <25 (<7.8%) | 315 (20.5%) |
| Yes | 2078 (96.9%) | 3556 (90.5%) | 1778 (97.6%) | 2338 (97.5%) | 300 (93.2%) | 1218 (79.5%) |
| missing | 7 | 22 | <10 | 8 | <5 | 14 |
| *Delivery mode* | | | | | | |
| vaginal | 1471 (69.2%) | 2472 (63.3%) | 1251 (69.3%) | 1661 (69.8%) | 220 (68.5%) | 811 (53.2%) |
| cesarean | 655 (30.8%) | 1434 (36.7%) | <555 (<30.7%) | 720 (30.2%) | <105 (<32.7%) | 714 (46.8%) |
| missing | 25 | 46 | <25 | 24 | <5 | 22 |
| *Preterm birth* | | | | | | |
| Term (≥ 37 weeks) | 1910 (89.7%) | 2711 (71.6%) | 1676 (92.0%) | 2198 (91.7%) | 234 (76.0%) | 513 (37.0%) |
| Preterm (<37 weeks) | 219 (10.3%) | 1074 (28.4%) | 145 (8.0%) | 199 (8.3%) | 74 (24.0%) | 875 (63.0%) |
| missing | 22 | 167 | 7 | 8 | 15 | 159 |
| *Birth weight for gestational age*[c] | | | | | | |
| Normal | 1623 (79.4%) | 2650 (78.8%) | 1400 (79.5%) | 1838 (79.3%) | 223 (78.8%) | 812 (77.8%) |
| Small | 115 (5.6%) | 228 (6.8%) | 96 (5.4%) | 119 (5.1%) | 19 (6.7%) | 109 (10.4%) |
| Large | 307 (15.0%) | 485 (14.4%) | 266 (15.1%) | 362 (15.6%) | 41 (14.5%) | 123 (11.8%) |
| missing | 106 | 589 | 66 | 86 | 40 | 503 |
| *NICU admission* | | | | | | |
| No | 593 (77.7%) | 756 (40.0%) | 593 (87.9%) | 756 (88.1%) | 0 (0.0%) | 0 (0.0%) |
| Yes | 170 (22.3%) | 1134 (60.0%) | 82 (12.1%) | 102 (11.9%) | 88 (100.0%) | 1032 (100.0%) |
| missing | 1388 | 2062 | 1153 | 1547 | 235 | 515 |

[a]Value for participant reflects the first sample collected
[b]"Other Races" includes participants who did not identify as White, Black, Asian, or Multiple Races as well as participants who identified as American Indian or Alaskan Native and participants who identified and Native Hawaiian or other Pacific Islander.
[c]This measure excludes non-singleton pregnancies; children of multiple gestation are listed as "missing"
[d]Asthma diagnosis was made after age 4; children followed fewer than 4 years cannot have asthma diagnoses
[e]Obesity was only determined for children with BMI measurements at age two years or older; children followed fewer than 2 years cannot have obesity diagnoses

| | All | | Fecal Samples | | Rectal Swabs | |
|---|---|---|---|---|---|---|
| | participant (n=2151) | sample (n=3952) | participant (n=1828) | sample (n=2405) | participant (n=323) | sample (n=1547) |
| Early Life Diet | | | | | | |
| *Ever breastfed* | | | | | | |
| No | 165 (8.9%) | 250 (7.0%) | <150 (<9.8%) | <175 (<8.7%) | <20 (<6.2%) | <80 (<5.2%) |
| Yes | 1681 (91.1%) | 3312 (93.0%) | 1379 (90.4%) | 1844 (91.4%) | 302 (94.1%) | 1468 (95.1%) |
| missing | 305 | 390 | <305 | <390 | <5 | <5 |
| *Ever formula fed* | | | | | | |
| No | 325 (26.8%) | 557 (21.9%) | 204 (22.3%) | 281 (22.0%) | 121 (40.7%) | 276 (21.7%) |
| Yes | 887 (73.2%) | 1990 (78.1%) | 711 (77.7%) | 997 (78.0%) | 176 (59.3%) | 993 (78.3%) |
| missing | 939 | 1405 | 913 | 1127 | 26 | 278 |
| *Breastfed at sample collection* | | | | | | |
| No | 613 (34.6%) | 1209 (39.0%) | 559 (38.5%) | 786 (42.2%) | 54 (17.0%) | 423 (34.3%) |
| Any breastmilk | 645 (36.4%) | 1166 (37.6%) | 572 (39.4%) | 741 (39.8%) | 73 (23.0%) | 425 (34.4%) |
| Exclusive breastmilk | 513 (29.0%) | 722 (23.3%) | 322 (22.2%) | 335 (18.0%) | 191 (60.1%) | 387 (31.3%) |
| missing | 380 | 855 | 375 | 543 | 5 | 312 |
| *Solid food consumption at sample collection* | | | | | | |
| No solids | 568 (47.0%) | 1539 (66.5%) | 470 (52.5%) | 665 (59.7%) | 98 (31.2%) | 874 (72.7%) |
| Any solids | 641 (53.0%) | 777 (33.5%) | 425 (47.5%) | 449 (40.3%) | 216 (68.8%) | 328 (27.3%) |
| missing | 942 | 1636 | 933 | 1291 | 9 | 345 |
| *TPN Use at sample collection* | | | | | | |
| No TPN | 713 (96.7%) | 1452 (96.5%) | 429 (100.0%) | 453 (100.0%) | 284 (92.2%) | 999 (95.1%) |
| Yes, TPN | 24 (3.3%) | 52 (3.5%) | 0 (0.0%) | 0 (0.0%) | 24 (7.8%) | 52 (4.9%) |
| missing | 1414 | 2448 | 1399 | 1952 | 15 | 496 |

[a]Value for participant reflects the first sample collected
[b]"Other Races" includes participants who did not identify as White, Black, Asian, or Multiple Races as well as participants who identified as American Indian or Alaskan Native and participants who identified and Native Hawaiian or other Pacific Islander.
[c]This measure excludes non-singleton pregnancies; children of multiple gestation are listed as "missing"
[d]Asthma diagnosis was made after age 4; children followed fewer than 4 years cannot have asthma diagnoses
[e]Obesity was only determined for children with BMI measurements at age two years or older; children followed fewer than 2 years cannot have obesity diagnoses

| | All | | Fecal Samples | | Rectal Swabs | |
|---|---|---|---|---|---|---|
| | participant (n=2151) | sample (n=3952) | participant (n=1828) | sample (n=2405) | participant (n=323) | sample (n=1547) |
| | | | Outcomes | | | |
| *Asthma diagnosis* | | | | | | |
| No | 827 (87.4%) | 1986 (84.1%) | 571 (84.8%) | 778 (84.6%) | 256 (93.8%) | 1208 (83.8%) |
| Yes | 119 (12.6%) | 375 (15.9%) | 102 (15.2%) | 142 (15.4%) | 17 (6.2%) | 233 (16.2%) |
| missing | 1205 | 1591 | 1155 | 1485 | 50 | 106 |
| *Any food allergy diagnosis* | | | | | | |
| No | 208 (57.3%) | 236 (43.1%) | <210 (<64.0%) | <235 (<60.1%) | <5 (<14.3%) | <5 (<3.2%) |
| Yes | 155 (42.7%) | 311 (56.9%) | <125 (<38.1%) | <160 (<40.9%) | <35 (<100.0%) | <155 (<99.4%) |
| missing | 1788 | 3405 | 1500 | 2014 | 288 | 1391 |
| *Eczema report* | | | | | | |
| No diagnosis | 571 (37.6%) | 1337 (43.0%) | 409 (34.0%) | 558 (34.9%) | 162 (51.3%) | 779 (51.6%) |
| Self report | 404 (26.6%) | 686 (22.1%) | 344 (28.6%) | 443 (27.7%) | 60 (19.0%) | 243 (16.1%) |
| Medical diagnosis | 544 (35.8%) | 1087 (35.0%) | 450 (37.4%) | 600 (37.5%) | 94 (29.7%) | 487 (32.3%) |
| missing | 632 | 842 | 625 | 804 | 7 | 38 |
| *Autism diagnosis* | | | | | | |
| No | 1269 (97.1%) | 2734 (97.4%) | 1063 (96.8%) | 1438 (96.8%) | 206 (98.6%) | 1296 (98.1%) |
| Yes | 38 (2.9%) | 73 (2.6%) | <40 (<3.6%) | 48 (3.2%) | <5 (<2.4%) | 25 (1.9%) |
| missing | 844 | 1145 | <735 | 919 | <115 | 226 |
| *ADHD diagnosis* | | | | | | |
| No | 1229 (95.1%) | 2530 (90.6%) | 1036 (95.5%) | 1403 (95.1%) | 193 (92.8%) | 1127 (85.4%) |
| Yes | 64 (4.9%) | 264 (9.4%) | 49 (4.5%) | 72 (4.9%) | 15 (7.2%) | 192 (14.6%) |
| missing | 858 | 1158 | 743 | 930 | 115 | 228 |
| *Obesity* | | | | | | |
| No | 961 (85.4%) | 2189 (86.4%) | 757 (84.4%) | 1013 (84.1%) | 204 (89.5%) | 1176 (88.4%) |
| Yes | 164 (14.6%) | 346 (13.6%) | 140 (15.6%) | 192 (15.9%) | 24 (10.5%) | 154 (11.6%) |
| missing | 1026 | 1417 | 931 | 1200 | 95 | 217 |

[a]Value for participant reflects the first sample collected
[b]"Other Races" includes participants who did not identify as White, Black, Asian, or Multiple Races as well as participants who identified as American Indian or Alaskan Native and participants who identified and Native Hawaiian or other Pacific Islander.
[c]This measure excludes non-singleton pregnancies; children of multiple gestation are listed as "missing"
[d]Asthma diagnosis was made after age 4; children followed fewer than 4 years cannot have asthma diagnoses
[e]Obesity was only determined for children with BMI measurements at age two years or older; children followed fewer than 2 years cannot have obesity diagnoses

Table S5. Participant characteristics by breastmilk consumption, example analysis

| | All (n=828) | None (n=255) | Any (n=319) | Only (n=254) | p-value |
|---|---|---|---|---|---|
| Age at collection (months) | | | | | >0.001 |
| mean (std) | 3.1 (1.1) | 3.4 (0.9) | 2.8 (1.1) | 3.00 (1.1) | |
| median [IQR] | 3.3 [2.1, 4.0] | 3.6 [3.0, 4.1] | 2.4 [2.0, 3.9] | 3.3 [2.0, 3.9] | |
| range | 1.0; 5.0 | 1.0; 5.0 | 1.0; 5.0 | 1.0; 5.0 | |
| Child sex | | | | | 0.75 |
| Male | 423 (51.1%) | 134 (52.5%) | 164 (51.4%) | 125 (49.2%) | |
| Female | 405 (48.9%) | 121 (47.5%) | 155 (48.6%) | 129 (50.8%) | |
| Delivery mode | | | | | 0.01 |
| Vaginal | 582 (70.3%) | 167 (65.5%) | 219 (68.7%) | 196 (77.2%) | |
| Cesarean | 246 (29.7%) | 88 (34.5%) | 100 (31.3%) | 58 (22.8%) | |
| Primer pair | | | | | >0.001 |
| Schloss V4 | 320 (38.6%) | 135 (52.9%) | 68 (21.3%) | 117 (46.1%) | |
| 515F-806R | 462 (55.8%) | 105 (41.2%) | 228 (71.5%) | 129 (50.8%) | |
| 515F-926R | 46 (5.6%) | 15 (5.9%) | 23 (7.2%) | 8 (3.1%) | |

#### Supplemental Figures

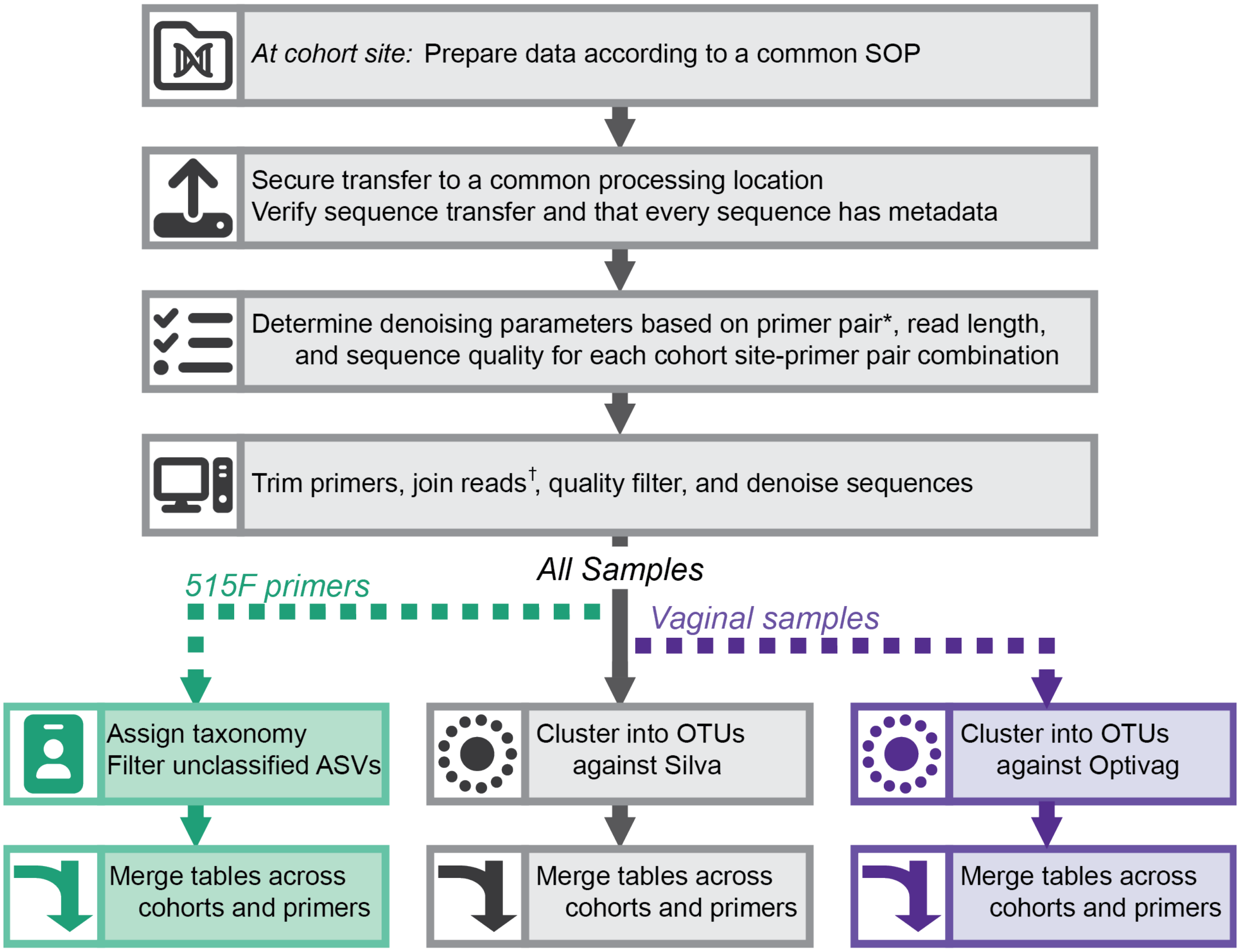


*All 515F-806R and 515F-926R were processed using the same parameters; Schloss V4 were trimmed consistently
†If appropriate to quality, read length, and primer pair

**Figure S1. Per cohort bioinformatic processing workflow.** Schematic shows the process of uploading and processing samples. All samples went through grey steps; teal steps were applied to samples amplified with 515F primers (515F-806R or 515F-926R); purple steps were applied to vaginal samples.

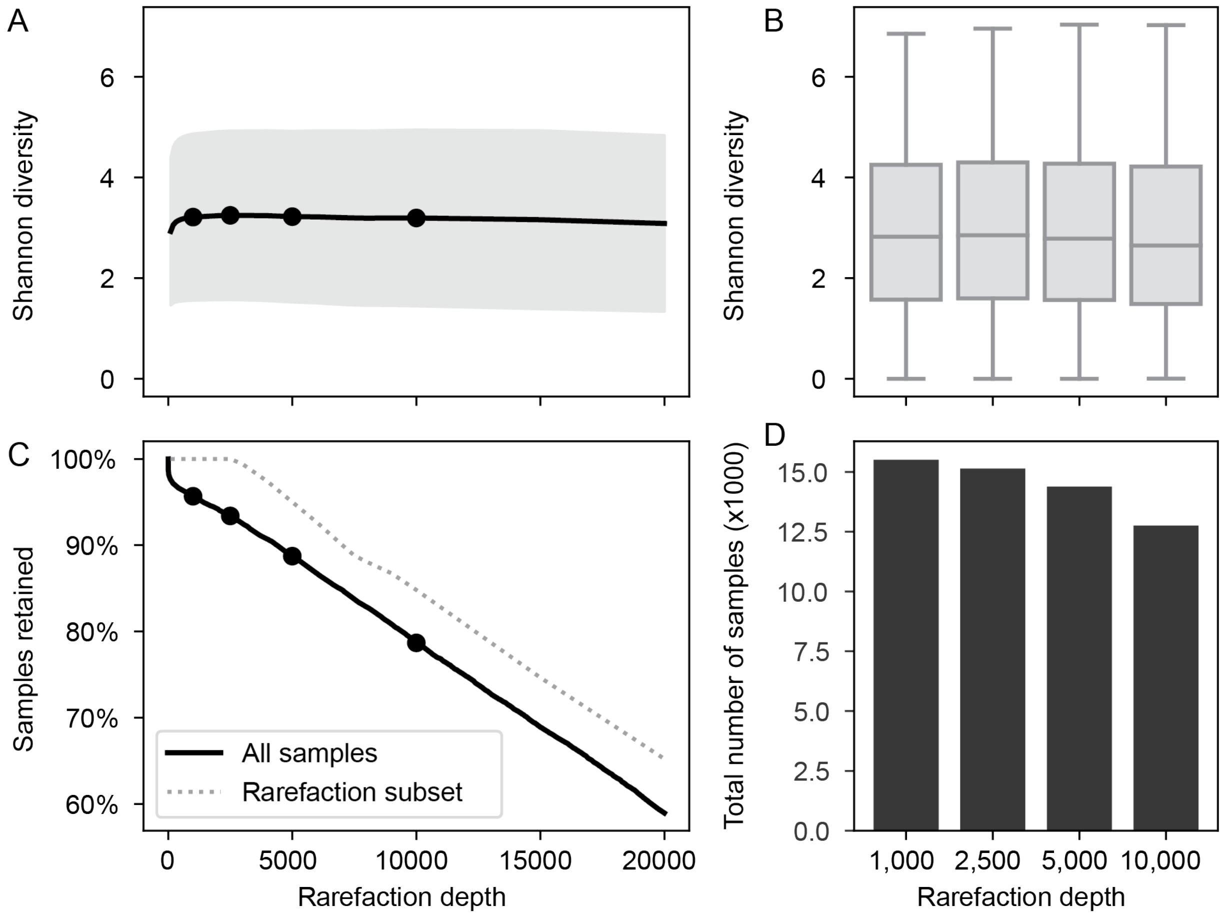


**Figure S2. All Sample Rarefaction Curve.** (A) Rarefaction curve showing the relationship between Shannon Diversity and sequencing depth for a subset of microbiome samples (n= 2482). The four pre-calculated rarefaction depths are highlighted as black dots. (B) The Shannon diversity across 15,139 samples at all four pre-calculated rarefaction depths. Not all samples are reflected in all boxplots. (C) The portion of samples retained as a function of sequencing depth. The solid black lines reflects all extant microbiome samples, the dashed line reflects the number of samples retained in the data set in panel (A). The four pre-calculated rarefaction depths are highlighted as black dots. (D) The number of samples retained as a function of rarefaction depth for all samples (black bars) and the rarefaction subset (gray bars).

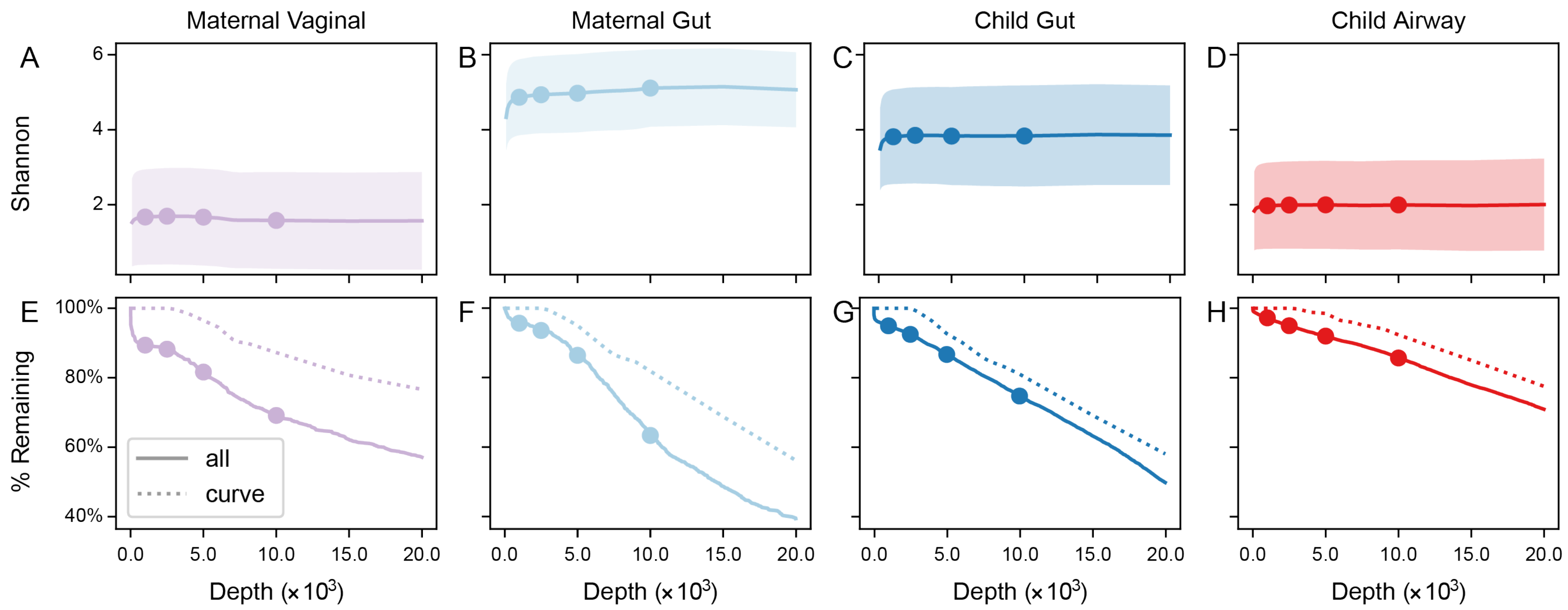


**Figure S3. Rarefaction Curve by body site.** (A-D) The average Shannon diversity as a function of depth for each body site. Shaded area shows the 95% confidence interval for the calculated depth. Points show the 4 pre-calculated rarefaction depths available in ECHO data: 1000, 2500, 5000, and 10,000 sequences/sample. (E-H) Gives the percentage of submitted samples remaining as a function of depth. The solid line shows all samples, the dashed line gives the percentage for the subset used to generate the rarefaction curve. Points highlight the four pre-calculated rarefaction depths. Body sites: (A,E) Maternal Vaginal, (B,F) Maternal Gut, (C,G) Child Gut, and (D,H) Child fecal.

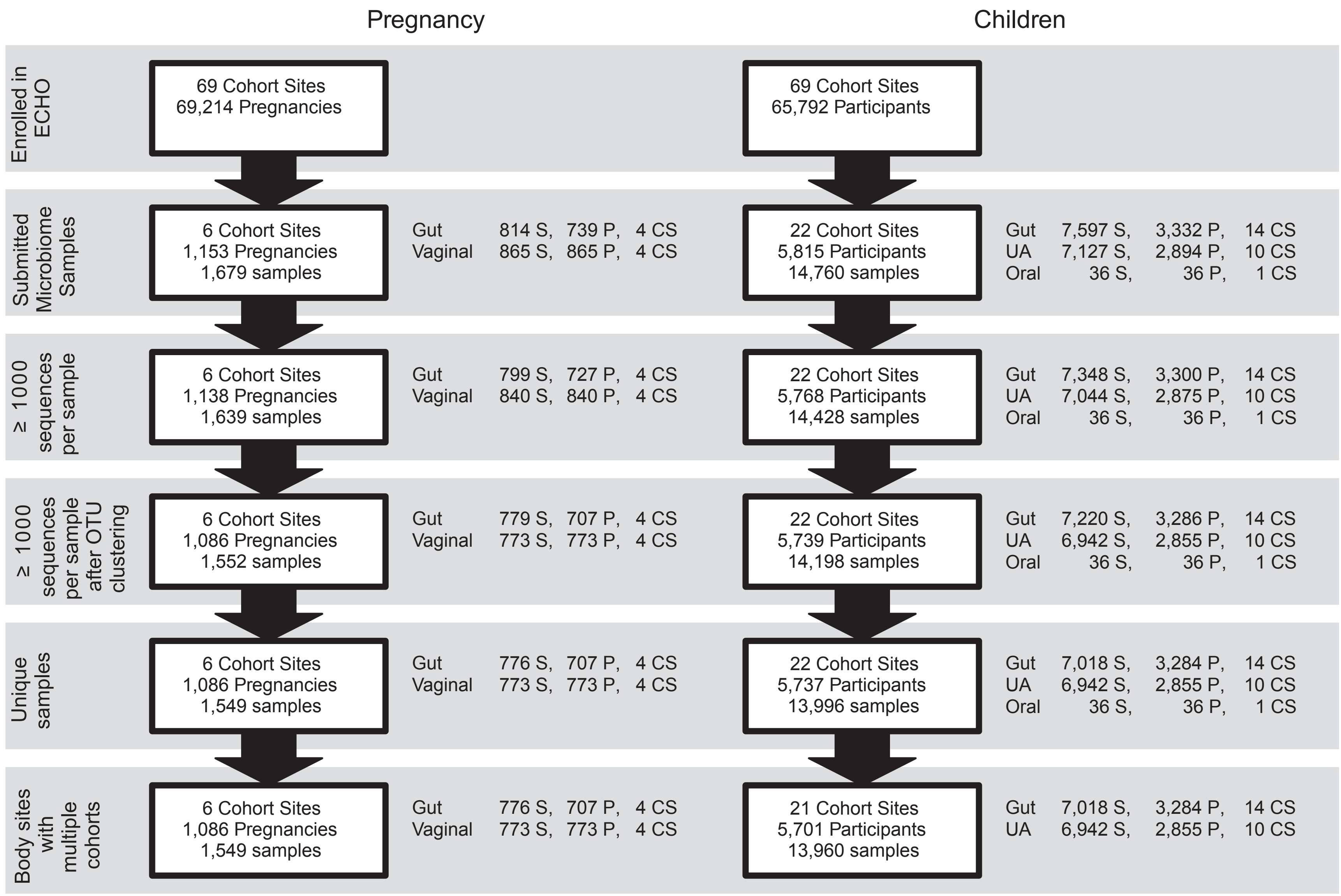


**Figure S4. Cohort, sample, and participant retention during bioinformatic processing.**

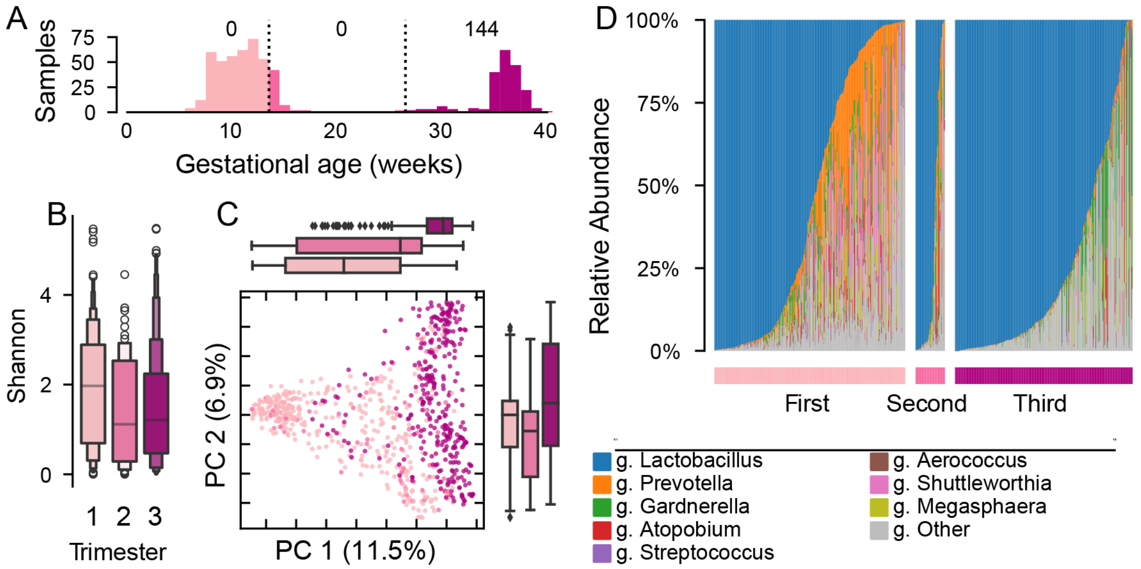


**Figure S5. Vaginal microbiome composition by trimester among participants in ECHO.** (A) Sample collection by week of gestation. Weeks are colored by trimester designation (light pink: first, mid-pink: second, dark pink: third); trimester periods are labeled with the number of samples in the trimester where the week is unknown. (B) Shannon diversity by trimester. (C) Principal components (PC) projection of Jaccard distance colored by trimester. (D) Genus-level taxonomic composition of vaginal samples.

ECHO, Environmental influences on Child Health Outcomes.

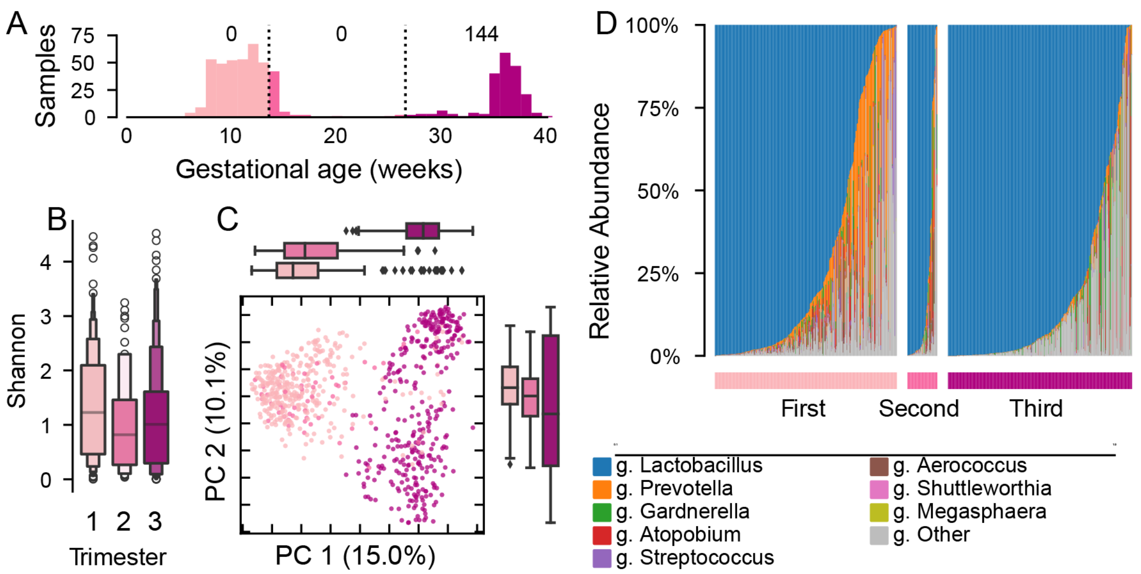


**Figure S6. Vaginal microbiome composition by trimester among participants in ECHO using the Optivag database.** There were 733 samples annotated with the Optivag database which retained at least 1000 sequences after clustering at 99% identity. (A) Sample collection by week of gestation. Weeks are colored by trimester designation (light pink: first, mid-pink: second, dark pink: third); trimester periods are labeled with the number of samples in the trimester where the week is unknown. (B) Shannon diversity by trimester. (C) Principal Components projection of Jaccard distance colored by trimester. (D) Genus-level taxonomic composition of vaginal samples.

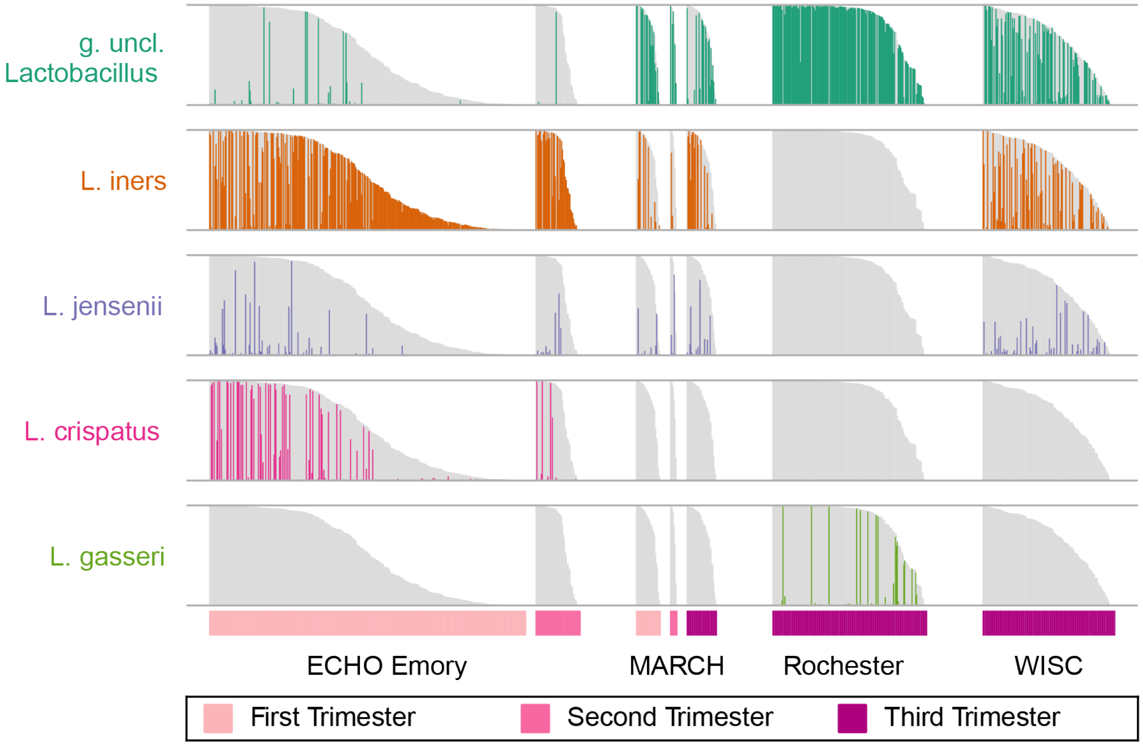


**Figure S7. Top Lactobacillus species in the Optivag data by cohort and trimester.** Each row shows the abundance of the 8 most abundant (cohort normalized mean abundance ≥ 1%) Lactobacillus species, organized by ECHO cohort site, trimester, and the Lactobacillus abundance. The gray bar in each row shows the total Lactobacillus in a given sample; the colored portions shows the abundance of the specific Lactobacillus species within the sample.

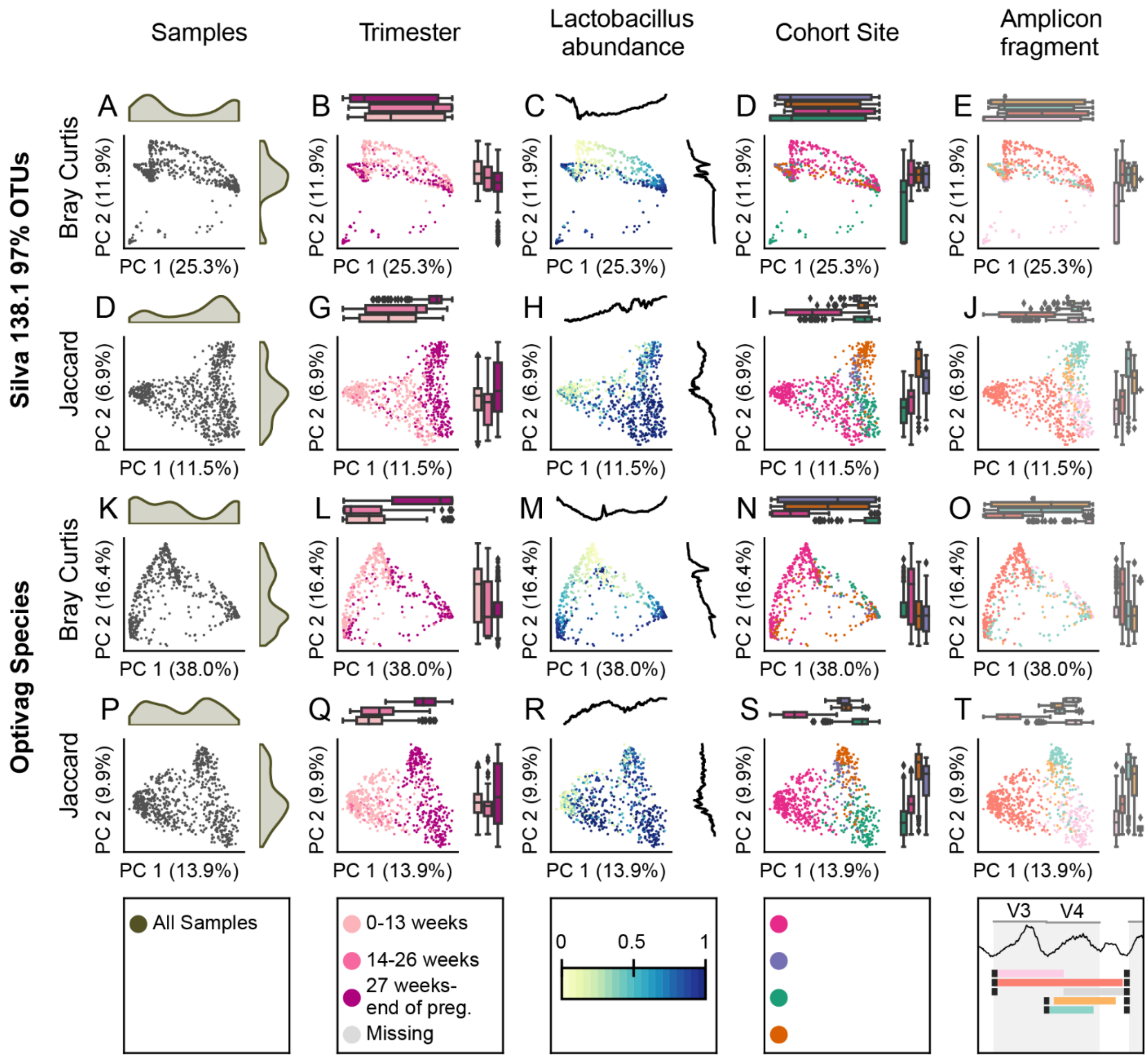


**Figure S8. Predictors of microbial community structure in vaginal samples.** Principal coordinates projections of (A-E) Silva 138.1 97% OTU Bray Curtis dissimilarity, (F-J) Silva 138.1 97% OTU Jaccard distance, (K-O) Optivag species Bray Curtis dissimilarity and (P-T) Optivag Species Jaccard distance. Each row reflects a metric; each column shows the relationship with a property of the microbiome or the data. The distribution of samples along the PC (A,F,K,P); the trimester of sample collection (B,G,L,Q); the relative abundance of L*actobacillus* (C,H,M,R); the cohort site which generated the sample (D,I,N,S); and the amplicon fragment used in the sample (E,J,O,T).

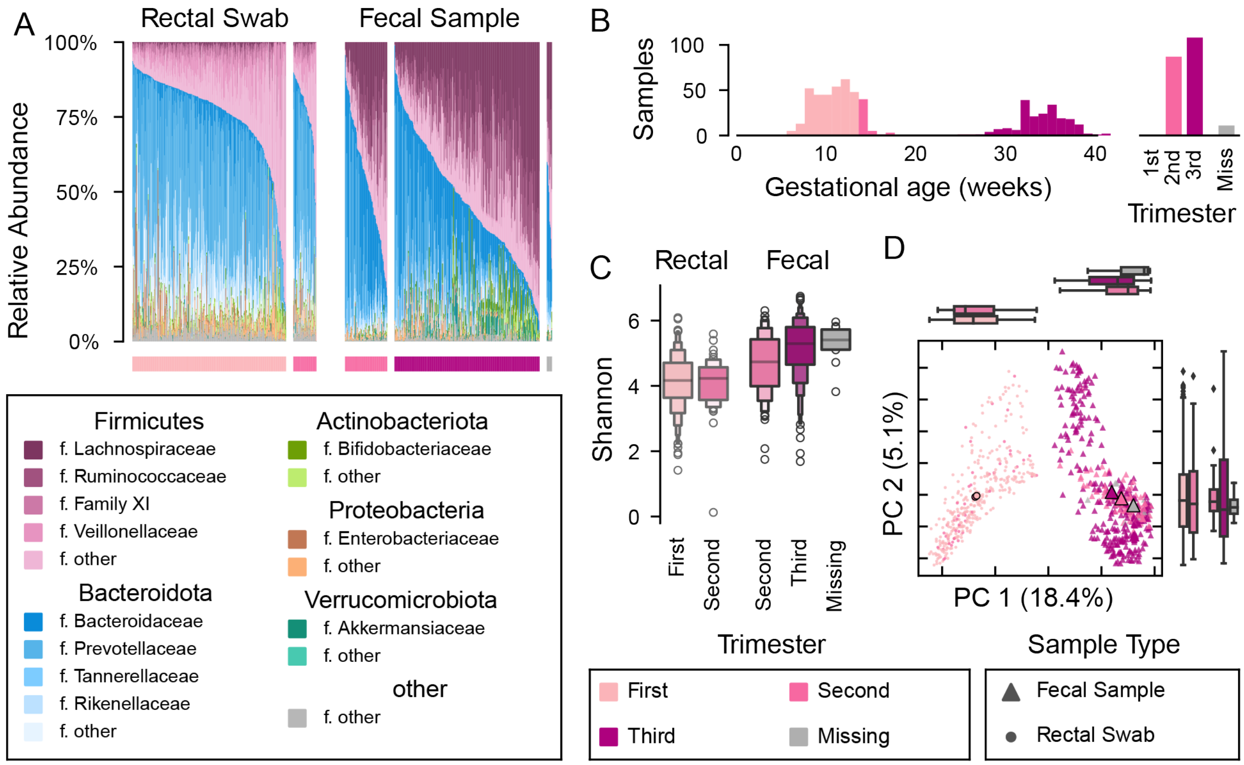


**Figure S9. Maternal gut microbiome is associated with trimester and sample type.** (A) Abundant phyla and families in maternal gut samples. Each bar represents a sample, organized by sample type, collection trimester, and Firmicutes abundance. The hue indicates a phylum level grouping; the shade indicates the family. (B) Sample collection by week of gestation. Weeks are colored by trimester designation (light pink: first, mid-pink: second, dark pink: third; gray missing). Counts for samples where the week of gestation was unknown are shown on an axis to the right. (C) Shannon diversity by sample type and trimester. (D) Principal components projection of Bray Curtis dissimilarity, colored by trimester. Rectal swabs are small circles; fecal samples are triangles. Larger symbols show the mean position for the sample type and trimester; marginal boxplots show the distribution of samples along the axis.

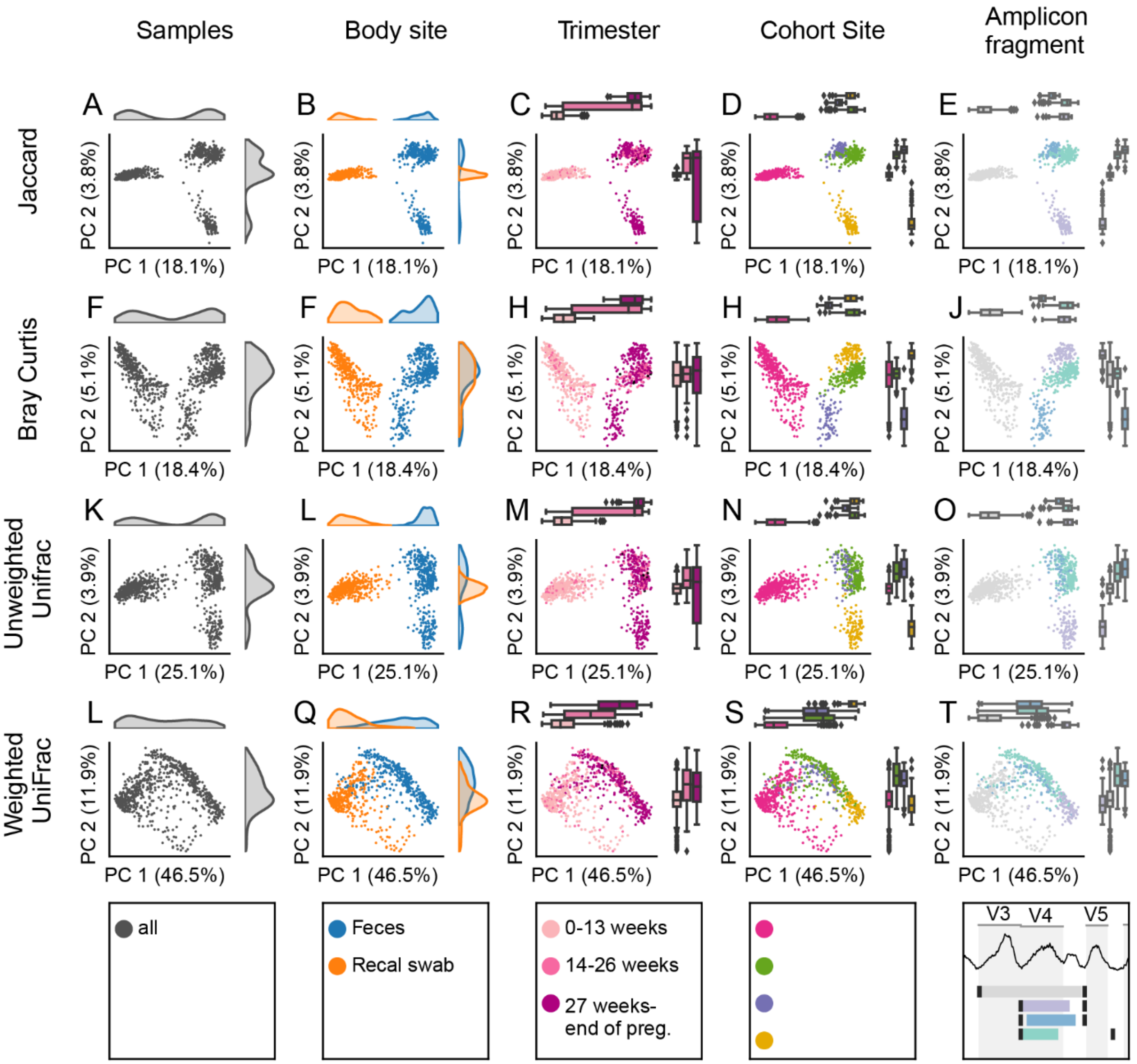


**Figure S10. Relationship between gut microbiome community composition and predictors of microbiome community structure.** PCoA of different measures of between community dissimilarity or distance are shown on each row: (A-E) Jaccard distances, (F-J) Bray Curtis dissimilarity; (K-O) unweighted UniFrac and (L-T) weighted UniFrac distance. (A,F,K,L) Density of samples along the axes; (B,F,L,Q) distribution of samples by specimen type; (C,H,M,R) colored by trimester; (D,H,N,S) colored by cohort; (E, J, O, T) colored by amplicon.

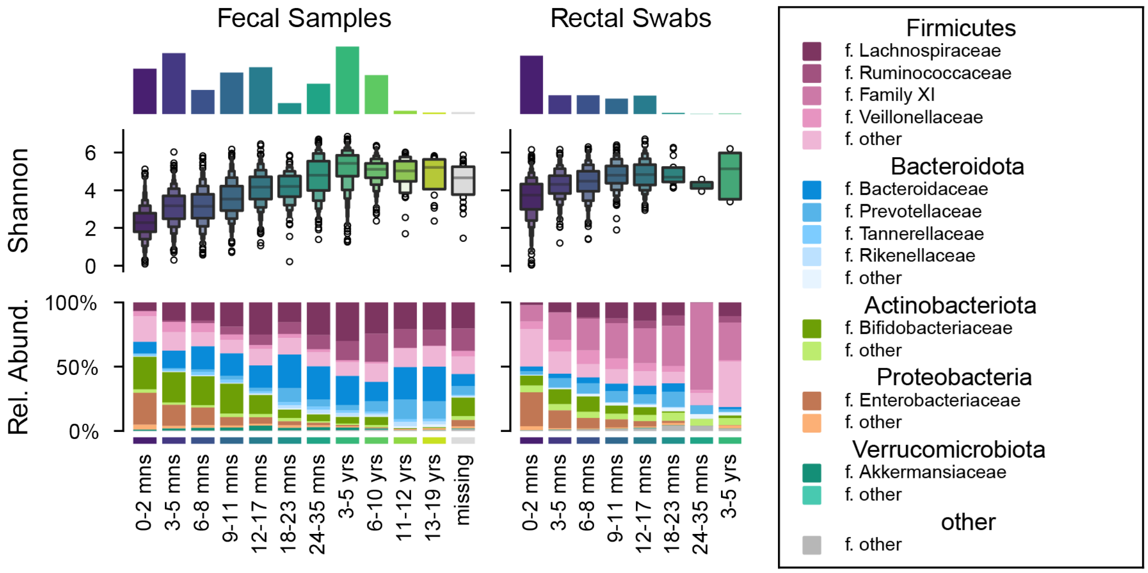


**Figure S11. Within sample diversity and taxonomic composition of child gut samples.** Top row shows the number of samples in each age group; the middle row shows the Shannon diversity; the bottom row shows the taxonomic composition.

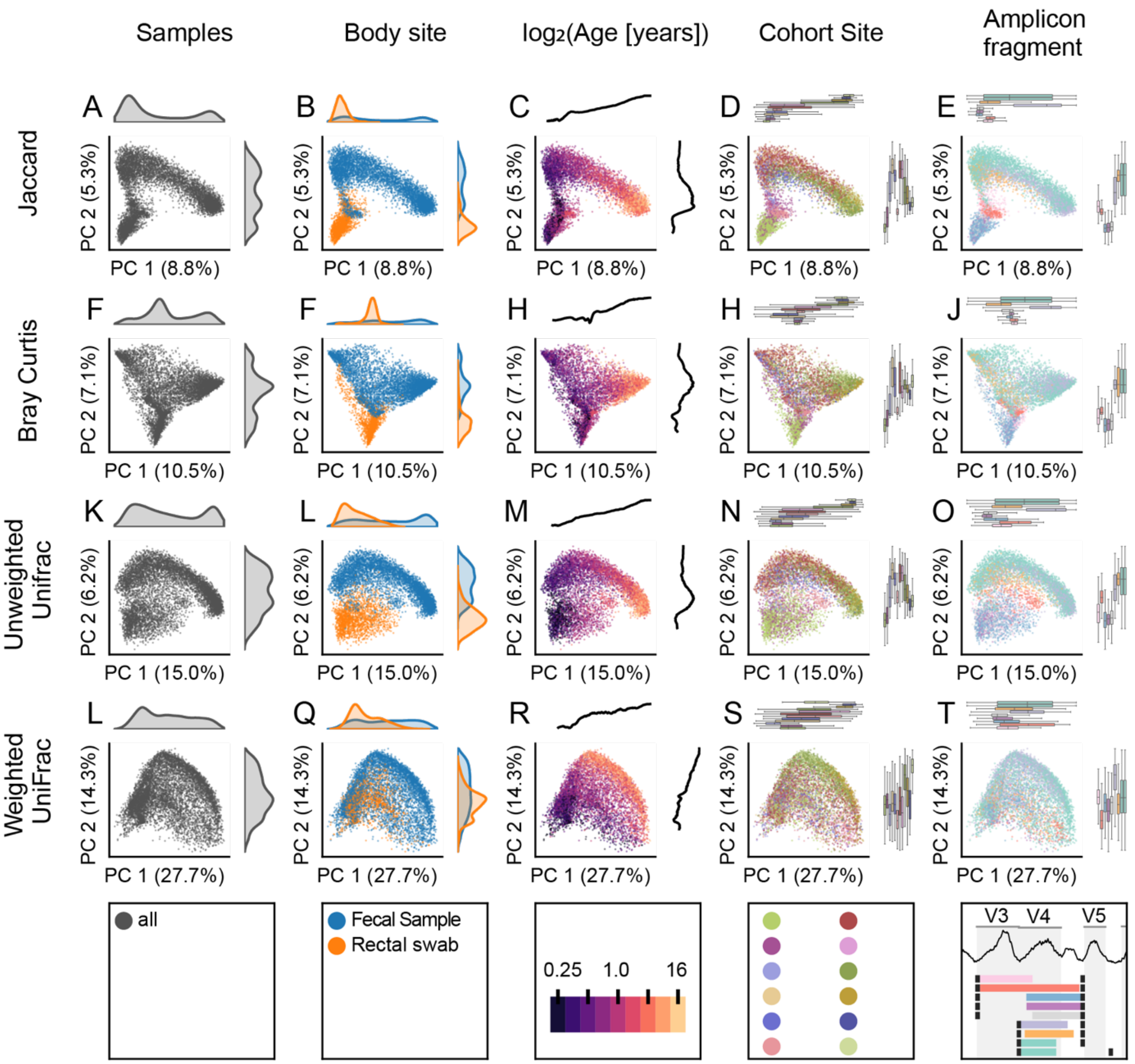


**Figure S12. Relationship between gut microbiome community composition and predictors of microbiome community structure.** PCoA of different measures of between community dissimilarity or distance are shown on each row: (A-E) Jaccard distances, (F-J) Bray Curtis dissimilarity; (K-O) unweighted UniFrac and (L-T) weighted UniFrac distance. (A,F,K,L) Density of samples along the axes; (B,F,L,Q) distribution of samples by specimen type; (C,H,M,R) colored by participant age; (D,H,N,S) colored by cohort; (E, J, O, T) colored by amplicon. Marginal plots show the distribution of samples along the axes; for age (C,H,M,R) the marginal plot shows the average age trace.

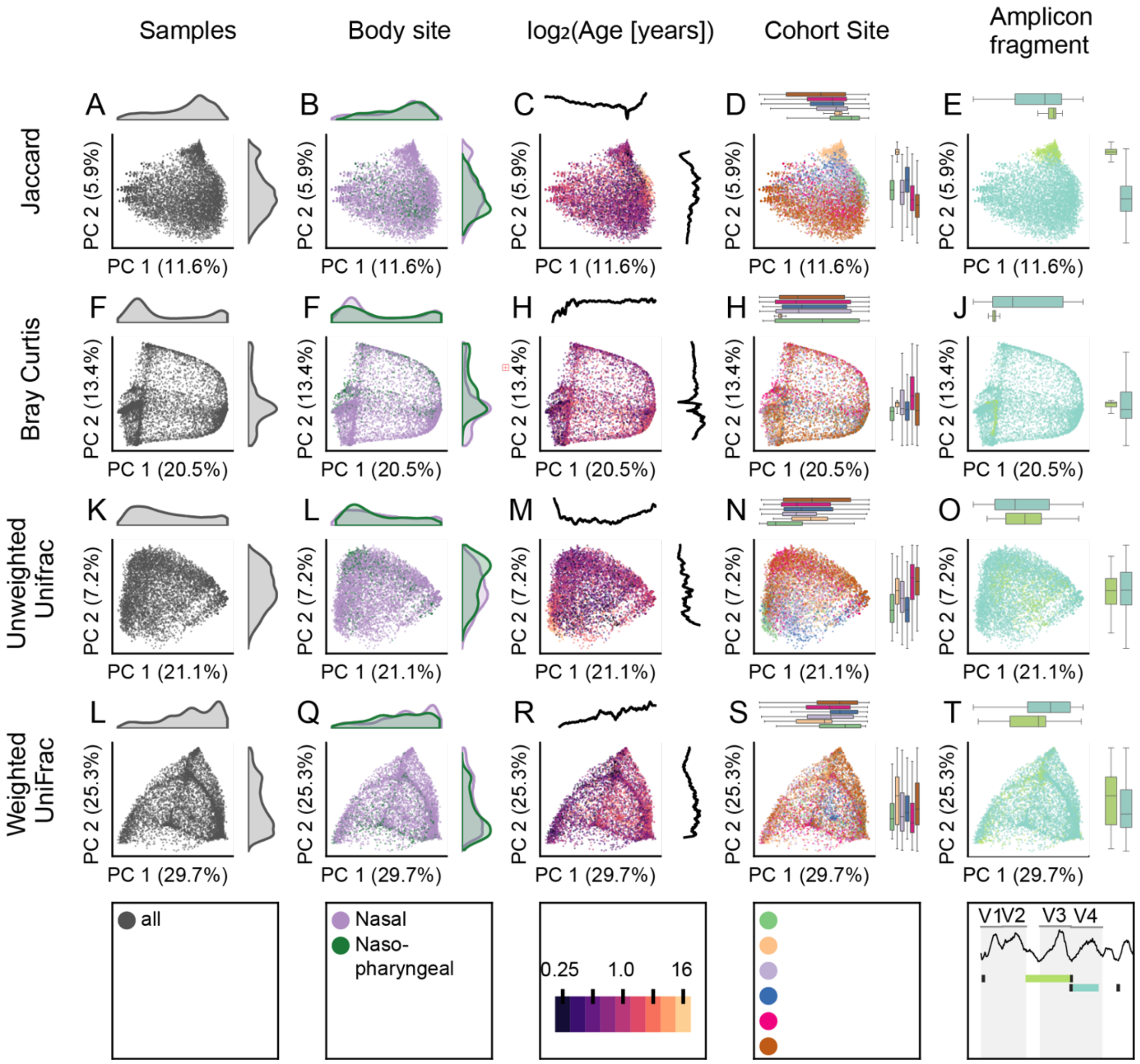


**Figure S13. Upper airway microbiome composition across different metrics and predictors.** Principal coordinate projection of (A-E) Jaccard distance, (F-J) Bray Curtis dissimilarity, (K-O) Unweighted Unirac distance, and (L-T) weighted UniFrac distance colored by (A,F,K,L) the distribution of samples, (B,F,L,Q) nasal (purple) or nasopharyngeal (green) sample type; (C,H,M,R) the log2 age in years at collection; marginal plots show the age (D,H,N,S) ECHO Cohort site, and (E,J,O,T) amplicon fragment.
